# COVID-19 Reproduction Numbers and Long COVID Prevalences in California State Prisons

**DOI:** 10.1101/2024.12.14.24319022

**Authors:** Lee Worden, Rae Wannier, Helena Archer, Seth Blumberg, Ada Kwan, David Sears, Travis C. Porco

## Abstract

Prisons have been hotspots for COVID-19 and likely an important driver of racial disparity in disease burden. From the first COVID-19 case detected through March 25, 2022, 66,684 of 196,652 residents of California’s state prison system were infected, most of them in two large winter waves of outbreaks that reached all 35 of the state prisons. We used individual-level data on disease timing and nightly room assignments in these prisons to reconstruct locations and pathways of transmission statistically, and from that estimated reproduction numbers, locations of unobserved infection events, and the subsequent magnitude and distribution of long COVID prevalence. Where earlier work has recommended smaller cells over large dormitory housing to reduce transmission, recommended use of cells with solid doors over those with bars only, and cautioned against reliance on solid doors (e.g., in cold months when HVAC systems can circulate aerosols), we found evidence of substantial transmission in both dorms and cells regardless of the door and season. Effective reproduction numbers were found to range largely between 0 and 5, in both cells and dorms of all door types. Our estimates of excess case rates suggest that as a result of disparities in incarceration, prison outbreaks contributed to disproportionate disease burden on Black and Indigenous people in California. We estimated that 9,100–11,000 people have developed long COVID as a result of infection in these prison outbreaks, 1,700–2,000 of them with disabling consequences, and that this burden is disproportionately on Black and Indigenous people in comparison to the state as a whole. We urge high-quality medical care for prison residents affected by long COVID, and decarceration to reduce the risk of future outbreaks of both COVID-19 and other diseases.

## 1. Introduction

The SARS-CoV-2 virus spreads readily in congregate settings, from nursing homes (Times, 2022; Krupar and Sadural, 2022; Solis et al., 2020) and crowded workspaces (Times, 2022) to places of worship (James et al., 2020; Katelaris et al.; Henry, 2020) and schools (Manica et al., 2023; Tseng et al., 2023; Times, 2022). Prisons and jails have housed many of the largest outbreaks (Wurcel et al., 2020; Saloner et al., 2020; Kinner et al., 2020; Dutheil et al., 2020; Solis et al., 2020; Times, 2022; Bradshaw, 2021; Hummer, 2020; Denney and Valdez, 2021; Duarte et al., 2022), and have been a focus of particular concern about pandemic control, because of the risks of both transmission within correctional facilities and exportation of infection to outside communities (Reinhart and Chen, 2020, 2021b,a; Jones and Tulloch, 2020; Barnert et al., 2020; Sims et al., 2021; Murphy, 2021; Towers et al., 2022; Wallace et al., 2021; Lofgren et al., 2022; Flagg and Neff, 2020).

This article presents an analysis of SARS-CoV-2 transmission in California’s state prison system from its emergence through March 25, 2022, at the level of individual residents and buildings within each of the 35 institutions (prisons) that were operated by the California Department of Corrections and Rehabilitation (CDCR) during this period. We used the locations of each resident each night, by room, building, and institution, together with dated SARS-CoV-2 test results and symptom reports, to estimate where and when transmission events occurred, and between which individuals, in order to differentiate more and less likely sites and dates of transmission.

We used this reconstruction to investigate a cluster of related questions. We investigated differences in transmission rates between dormitory and cell settings, and between locations with and without solid doors, and seasonal differences in transmission rates in these different types of rooms; we constructed a detailed look at the times and locations of disease transmission and the types of rooms involved; and we projected the times and locations of transmission forward in an estimate of overall rates of long COVID due to infection in CDCR institutions, its distribution over times and locations, and the distribution of both acute and long COVID disease over race/ethnicity and gender. We use these results to discuss implications for ongoing prevention and response to future epidemics, for assessment of the impacts of past management measures, and for care for individuals experiencing ongoing effects of past infection.

In reconstructing transmission rates, we specifically estimated *daily effective reproduction numbers* per day and building in each institution. Reproduction numbers, conventionally referred to by the letter *R*, are a key indicator of the spread of an infectious disease in a population, describing the number of new cases who acquire the virus from a given infected person. Sustained spread of a disease requires a sufficient number of cases’ reproduction numbers to be greater than one, in order to produce an overall steady or increasing number of cases. The magnitude of a reproduction number provides an indication of the effectiveness of possible interventions, such as physical distancing, masking, vaccination, ventilation, and air filtration: for example if reproduction numbers are between one and two, interventions that prevent half of all transmission events should be able to bring the outbreak to a stop. A daily or instantaneous effective reproduction number is a description of the changing state of an outbreak in the form of an estimate of the average reproduction number at one moment in time. It expresses how many cases a given case would cause if the current conditions were to persist unchanged throughout the entire infectious period of that case. Note effective reproduction numbers are distinguished from basic reproduction numbers, conventionally labelled *R*_0_, which refer to the number of cases produced by a case in a population that has not previously been exposed, while an effective reproduction number describes the number of cases caused by a case in the population as it is, at a specific time.

In this analysis of California prisons, effective reproduction numbers can be used as a description of the rate of spread of SARS-CoV-2 on a given day in a given building. Here we present a comparison of estimated effective reproduction numbers by date across different types of housing, given by classifying rooms as dormitories and cells, and their doors as solid and porous.

The differences between locations within a prison with respect to facilitating or preventing SARS-CoV-2 transmission are of particular interest, and in order to analyze difference between locations in this setting it is necessary to take care with timing. Because case detection can occur at a delay from infection, and residents have often been moved from location to location during an outbreak for quarantine and isolation purposes, an individual can be housed in a different location when a positive test occurs from the location where they were infected. For this reason we have ensured that our analysis accounts for the timing of transmission events in the days before detection of any case, to reconstruct the time and location of the actual transmission event preceding each positive test. We used published data on the timing of sensitivity of RT-PCR and antigen tests for SARS-CoV-2, together with the daily movements of residents recorded by CDCR and the timing of their test results and symptom reports, to pinpoint as accurately as possible the true times and places of transmission. This information was used in the construction of effective reproduction numbers for each day in each building, accounting for movements and likely delays between transmission and case detection, so that differences in reproduction numbers and incidence between locations can be assessed.

Because of long-standing disparities in incarceration, correctional institutions have been an important source of racial inequity in disease burden. Black and Indigenous people are severely overrepresented in incarcerated populations (Alexander, 2020; Schlesinger 2008; Wang, 2021; Nowotny et al., 2021; Schlesinger, 2022), and the California system is no exception (Klein et al., 2023; Hayes et al., 2022). The racial composition of the CDCR prisons is reflected in the racial composition of their COVID-19 cases (Chin et al., 2021; Kwan et al., 2022), and the racial distribution of the long-term burden of post-acute disease in the system is therefore a concern as well. For this reason we attended carefully to the racial/ethnic distribution of both the COVID-19 infections in the system and the estimated long COVID cases resulting from those infections. We approached the question in two ways: first, by analyzing associations between *per capita* infection rates and race/ethnicity while controlling also for gender, age, and location; and second, by combining prison infection rates with an estimate of California rates to estimate excess case rates attributable to state prison outbreaks by race/ethnicity. The latter estimate is used to discuss the impact of prison transmission on the burdens of SARS-CoV-2 infection and long COVID in the state.

### Ethics and IRB approval

This project was approved by the IRB at the University of California, San Francisco (IRB no. 21-34030). The IRB waived the need for informed consent given the use of secondary deidentified datasets based on the following criteria: deemed no more than minimal risk to subjects; could not practicably be done without the waiver; will not adversely affect rights and welfare of subjects; and will provide subjects with additional pertinent information after participation.

## 2. Methods

### 2.1. Terminology

The California Department of Corrections and Rehabilitation (CDCR, formerly California Department of Corrections) refers to the prisons it operates as institutions. The buildings of each institution are grouped into facilities, informally known in some cases as yards, with multiple buildings per facility and often multiple facilities per institution. The people incarcerated in these institutions are referred to as residents. The names and standard abbreviations for the 35 institutions active during the period studied^1^ are listed in Table L.1 (Supplement L). Health care in CDCR institutions is managed by a division within CDCR known as “California Correctional Health Care Services (CCHCS).”

### 2.2. Data

Data provided by CDCR recorded the room locations of individual prison residents in each institution’s multiple buildings on each day, and the type of room in which they were housed, from Jan. 1, 2020 through March 25, 2022. For all SARS-CoV-2 cases, it also included dates of symptom onset where available, as well as dates and results of all RT-PCR and antigen tests administered to residents at all CDCR institutions. Only deidentified data were made available for analysis.

While the locations of prison residents at the institution level were identified by institution names, buildings, rooms, and facilities within institutions were identified only by anonymized numbers. We report results by building using these numerical IDs. A classification of room types was provided. We combined several types of cells and dorms into categories `Cell` and `Dorm`. The types `Room`, `Closed Ward`, and `Other` were rare and were combined into the classification `Uncategorized`.

A classification of rooms by solid versus permeable barred doors was compiled by UCSF and Berkeley researchers from information provided by CCHCS. We used it to construct a combined “room type” for each room consisting of one of the above types, `Cell, Dorm`, and `Uncategorized`, together with a door type, `Open, Closed`, and `Uncategorized`.

Individual demographic characteristics recorded included birth year, “phenotypic sex” and race. Race was recorded using a system of nine classifications, of which eight were used in the population analyzed (the Unknown category was not present). We collapsed the variable to six levels by combining the categories `Mexican, Hispanic`, and `Cuban` into a single `Latinx` group, for comparability to California data and to avoid possible deductive disclosure when group sizes are small.

The sex variable was not used as is, but was interpreted together with location to impute gender, as follows. The institutions are segregated into women’s and men’s facilities, and the majority value of the sex variable in each building matched this classification. California’s State Senate Bill 132 (SB 132), effective January 1, 2021, requires CDCR to house prison residents according to their self-identified gender by request (The Moss Group, Inc., 2022), and a number of residents are housed in facilities that do not match their recorded sex. We labeled anyone for whom the sex variable matched the building’s majority at all times as cisgender man or cisgender woman (CM, CW), and all others as transgender, nonbinary, or intersex (TGNBI). Due to the small group size and lack of explicit gender listings, we did not attempt to distinguish between transgender women, transgender men, and others labelled TGNBI. Because SB 132 was not in effect in 2020, because many requests for transfer under SB 132 have reportedly been unfulfilled (Sosa, 2023), and because some may not request housing reassignment, it is likely that these imputed genders are incomplete in some cases, and that the TGNBI category here represents a subset of transgender, gender-nonconforming, nonbinary, and intersex residents. We include this gender variable to gain a view into the effects of gender on disease risk.

### 2.3. Total case counts

Following CDCR’s reporting policy (California Department of Corrections and Rehabilitation, n.d.a), we considered a positive test result after 90 or more days without positive test results to indicate a new case, which may have been a reinfection of a previously infected individual. We compiled the number of positive and negative tests administered by day, new cases detected by day, and seven-day average of cases detected by day in each institution, and in each building.

We used SARS-CoV-2 test results by date to identify distinct outbreaks at each institution, using the California Department of Public Health’s definition as a collection of case detections separated by fourteen or fewer days (California Department of Public Health, n.d.). Cases were considered to be detected on the date of their first positive test. We aggregated the number and sizes of outbreaks at each institution.

### 2.4. Transmission reconstruction

We conducted a computational process of Bayesian estimation to estimate incidence dates and effective reproduction numbers for SARS-CoV-2 transmission for each individual and building of the CDCR institutions on each day. Figure 1 offers an outline of key steps, and see Supplement A for more details.

**Figure 1:**
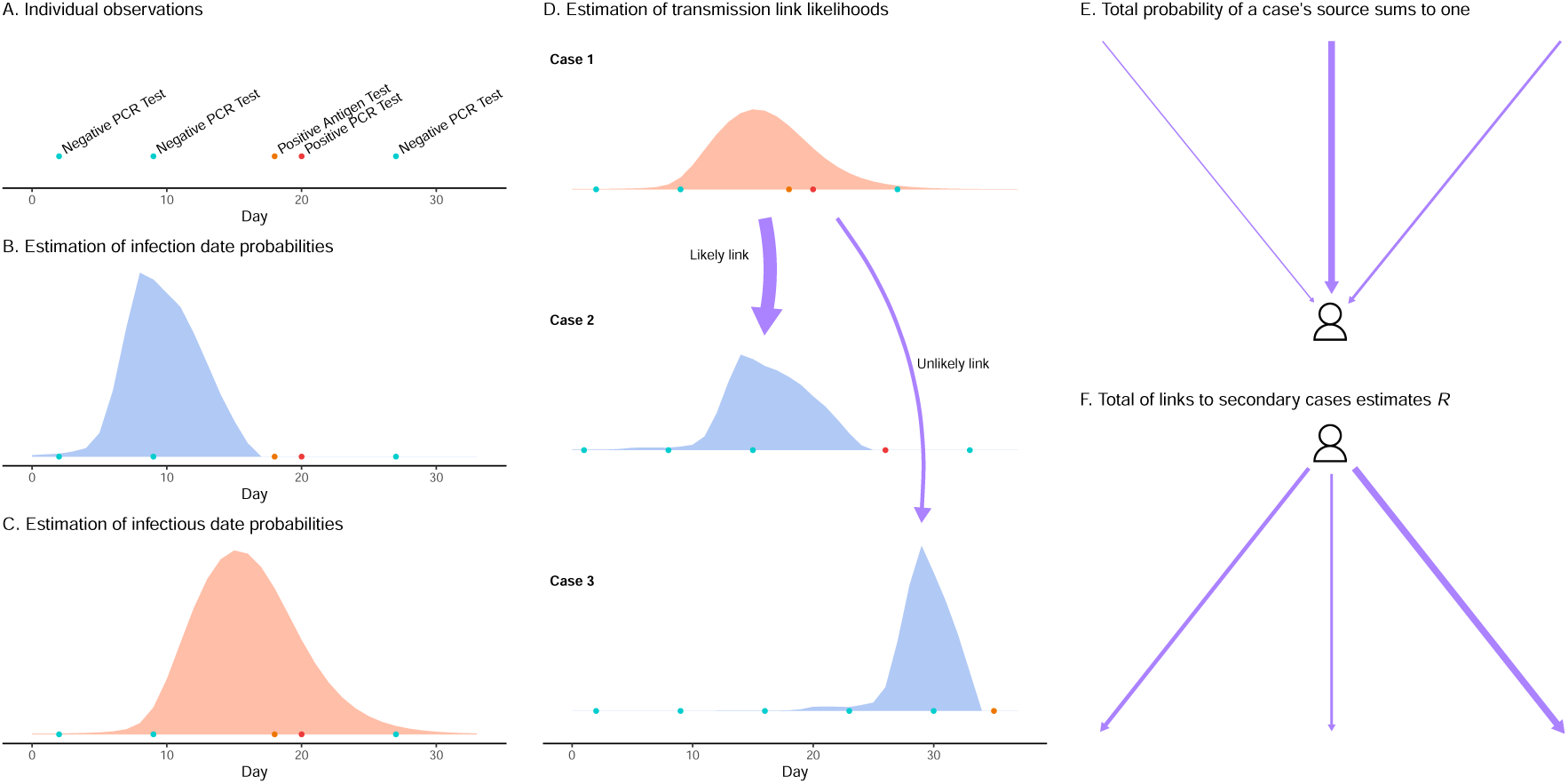
**Key steps in the estimation model**, which builds on the work of Wallinga and Teunis (2004). (A.) Data collected by CDCR provides **dates of positive and negative SARS-CoV-2 tests** administered on individuals (who are identified by anonymized ID numbers), and in some cases (not shown here) reported dates of onset of symptoms. All case histories shown in this figure are fictional. (B.) For each individual, a probability is estimated for each potential **date of infection** based on the probabilities of negative and positive test results and symptom onset at different times during infection. Here, the probability is nonzero on days before the first positive test, and drops off before the earlier negative tests, because the probability of a false negative result declines as the time from infection date to test date increases. (C.) Along with the infection dates, a probability distribution is estimated for **dates the individual was infectious**, by estimating probabilities for the time from infection to transmission and using the above estimate for dates of infection. Specifically, if the individual was to be exposed to susceptible individuals at a constant rate, and one transmission event occurred, this estimates the probability for the day of transmission. (D.) A likelihood is estimated for all **possible transmission links** between all pairs of individuals: the likelihood is higher if the first person’s infectious period and the second person’s infection date are more likely to coincide. (Not shown: likelihood of transmission is also higher on days when individuals were housed in the same room or building; likelihood is assigned to dates of transmission as well as pairs of individuals.) (E.) **Probabilities for all possible transmission links** are estimated from the likelihoods by scaling to reflect that each case has one source. (F.) **Reproduction numbers** are estimated by the expected number of cases infected by each person, based on the above transmission link probabilities. (Not shown: estimation of link probabilities, reproduction numbers, and incidence by date. See appendix for details.)

We used CDCR’s data set to provide dates and results of all RT-PCR and antigen tests administered to residents at all CDCR institutions, dates of onset of symptoms reported for residents, and daily locations of residents at the institution, building, and room levels.

We estimated the likelihood that each resident was infected on a particular day by combining several sources of information. Established estimates of the performance of RT-PCR and antigen tests for the virus were used to combine all positive and negative test results recorded for an individual, together with reported symptom onset dates where available, to construct a probabilistic estimate of date of infection and duration of incubation period for each infected individual. Because symptom onset dates were reported to contain inaccuracies due to record-keeping difficulties experienced during outbreaks, we used a relatively vague prior distribution for symptom onset dates compared to test results (see Supplement A for details). This allowed us to estimate how likely each resident was to have been infected on each day, and how likely they were to transmit the virus to others on each day, based only on their individual test and symptom history.

We then used these estimates together with residents’ daily locations to estimate how likely transmission between individual pairs of residents was to occur on each day. From this we generated an updated estimate of each case’s incidence date and of how many cases were infected by each infected resident each day, and from that effective reproduction numbers (*R*) by building by day, and the number of cases infected in each building each day. This estimation process operated on all cases in an institution together, and was applied to each institution in the CDCR system, generating daily building-level estimates of *R* for all outbreaks in all institutions in the time period studied.

These estimates accounted for the daily movements of residents between buildings, and provided a description of the dynamically changing conditions in each building by day as individuals become infected, recover, and shift locations. Because these estimates took into account where individuals were day by day when they were likely to have been infected, where they were when they were likely to be infectious, and which individuals were in proximity and infectious on days when each other individual at the institution may have become infected, they provided an estimate of which locations within an institution were likely to have been sites of transmission each day. Because reproduction numbers can be understood to reflect local conditions conducive to spread of the disease, these estimates could be used to evaluate the relative safety of different settings. To examine the difference between location of residents at infection and their location at case detection, we compared the number of case detections of individuals housed in each room to the number of incidence events attributed to that room by the estimate, using a linear regression with intercept of zero, with room type as an exposure variable.

After generating the above described estimates for each institution in the CDCR system, we combined them to provide a summary of estimates across the system of prisons. We visualized the distribution of *R* values pertaining to infectious individuals by institution and in each building. We compared *R* values stratified by room and door type, to investigate the hypotheses that cells reduced transmission compared to dorms and that solid doors reduced transmission compared to permeable ones.

In the Results section below we present summaries of the results over the 35 institutions of the CDCR system. In Supplement I, we present the results from each institution in detail, in one panel of figures for each CDCR institution. Supplement J presents one panel for each room type, showing the estimated overall incidence and reproduction number by day in each institution, over all rooms of the given type only. Supplement A documents the estimation methods in detail.

### 2.5. Distribution of cases

Adjusted *per capita* infection risk is relevant to comparing infection rates between groups in the prison population, to infer whether some groups faced higher infection risk than others in the outbreaks that occurred. Previous analyses of some of these outbreaks have shown that *per capita* the risk of infection was distributed relatively evenly across racial/ethnic groups, without the disproportionately high rates among Black, Latino, and Indigenous people that were documented in community transmission in 2020–21 (Chin et al., 2021; Kwan et al., 2022). We performed a similar analysis on the series of outbreaks considered here.

First, we compiled as summary statistics the number of infection events by race/ethnicity and gender. Second, along with the above statistics, we analyzed associations between gender, race/ethnicity, and the interaction of the two and infection risk, to investigate whether different populations were exposed to different risk in the prisons without comparison to the state. Using our reconstruction of incidence dates and locations, we summarized estimated incidence by person and day into number of infection events during each person’s stay in each building where they were housed. Each person’s duration of stay in each building was counted in years and fractions of years. We performed a mixed-effects Poisson regression analysis of the incidence against gender, race/ethnicity, the interaction of gender and race/ ethnicity, age (in age classes under 23, 23–35, 35–50, and over 50), and duration of stay, with location at the building level as a random intercept. We tested multiple null hypotheses: one for each level of the gender and race/ethnicity variables, of no difference from the population-wide rate of infection, and one for each level of the gender-race/ethnicity interaction, of no difference from the infection rate predicted by gender and race/ethnicity independently, with Bonferroni-Holm adjustment for multiple comparisons (see Supplement B for details).

### 2.6. Estimation of excess cases

While the above analysis addresses differences in disease risk given the populations and conditions that existed during these outbreaks, at the same time, such analysis of individual-level risk does not fully describe the burdens and impact of the disease spread in the prisons. The impact of prison outbreaks on the state can be framed as a question of what would be different in the state in their absence. The answer to such a question can only be inferred and estimated. Here we approach it as modelers by way of two counterfactual scenarios.

First, we compiled the raw number of CDCR cases by race/ethnicity in the time period studied, and we note that this statistic provides a very basic counterfactual comparison, namely the number of cases that would not have occurred if every prison resident were imprisoned as they were but COVID-19 transmission into prisons had been prevented.

Second, we additionally constructed an age- and race/ethnicity-adjusted estimate of the number of excess cases in state prison outbreaks compared to community rates of infection, to assess how the occurrence of these prison outbreaks affected the overall number and distribution of cases in the state. This allows a comparison of the impact of these outbreaks to what the disease burden might have been in a counterfactual scenario in which all the residents affected had been in the community and infected at community rates. This second estimate was conducted as follows.

We calculated the cumulative incidence rate by day in each institution from new case detections relative to population size on March 20, 2020, the first day a case was detected in a CDCR prison. We used this reference population size because it was the size before any adjustments to release and intake rates made in response to outbreaks.

Comparison of overall case counts between the CDCR system and the state of California required an estimate of the true number of cases, since case ascertainment rates may have been very different in the two settings. Testing rates in the CDCR system were relatively high: from March 2020 to October 2021 94% of all residents were tested, and residents who were tested were tested an average of 16 times (Kwan et al., 2022), while testing in California was likely much less frequent. Rather than compare counts of confirmed cases directly, we used an estimate of the state’s true number of cases published by the Institute for Health Metrics and Evaluation’s (IHME) estimate of incidence (Institute for Health Metrics and Evaluation, 2022; IHME COVID-19 Forecasting Team, 2021). We adjusted the California estimate by removing incarcerated people from the total infections and population, to estimate community incidence rates in the state excluding carceral institutions. CDCR resident numbers were drawn from the CDCR data described above, while numbers from federal prisons, county jails, and ICE facilities were obtained from UCLA’s COVID Behind Bars project (UCLA Law COVID Behind Bars Data Project, n.d.; Dolovich and Littman, 2020) and adjusted using a broad span of values for the unknown case ascertainment rate (see Supplement D for details). By comparing this estimate to the cases detected within the CDCR system, we obtained a conservative estimate of the excess incidence in the CDCR system. Since the case ascertainment rate in the prisons is likely very high, though not perfect, the estimated excess cases can be used as a low estimate of the true excess cases.

We split the time period studied into three waves: the early outbreaks, the winter of 2020–21, and the subsequent outbreaks, which were predominantly the winter 2021–22 outbreaks. We compared the overall incidence in each wave between the CDCR system and the state, as a fraction of total population. In waves in which the CDCR incidence rate exceeded California’s, we estimated the number of excess cases by race/ethnicity by comparing CDCR incidence to what it would have been at California rates, by race/ ethnicity and age. The estimation of excess cases was conducted as follows.

The count of case detections during the relevant period in California by race/ethnicity was downloaded from California Health and Human Services’s Open Data Portal (California Health and Human Services Open Data Portal, 2023). The ascertainment rate for the state was estimated by the ratio of IHME-estimated total infections to total case detections. True infections and incidence rates in infections per person-year, by race/ethnicity, were then estimated from the ascertainment rate, case detections by race/ ethnicity, and population sizes by race/ethnicity. Age-specific incidence rates by race/ethnicity were estimated from the above using known odds ratios for infection by age (Talaei et al., 2022) and California age distributions stratified by race/ethnicity. California’s “Two or more races” counts were counted as “Other” for these estimates.

A counterfactual estimate of the number of CDCR residents who would have been infected at California rates, by race/ethnicity, was generated from the total person-years spent in CDCR institutions by race/ ethnicity and age using the above incidence rates. From that an estimate of excess cases in the CDCR system by race/ethnicity was derived by subtraction, and the rate of excess CDCR cases relative to California population by division. This sequence of steps is detailed and illustrated in Supplement D.

### 2.7. Long COVID model

While there is much uncertainty about the nature and impact of post-acute effects of SARS-CoV-2 infection, estimates of their prevalence and duration are available (e.g. Davis et al., 2023). We used results from several studies to estimate the epidemiology of long COVID and resulting disability among the known cases in the CDCR resident population. We obtained a baseline rate of long COVID occurrence given infection from a Dutch study (Ballering et al., 2022), age-specific rates from a U.S. study (FAIR Health, 2022), rates by gender and race/ethnicity from the U.S. CDC (CDC, 2023), protection due to vaccination from U.S. and U.K. studies Al-Aly et al. (2022); Ayoubkhani et al. (2022), an estimated rate of recovery from long COVID from a French study (Tran et al., 2022), and a conservative estimate of disability resulting from long COVID from U.K. data (Ayoubkhani et al., 2021; Spiers, 2022) (see Supplement C for details). We constructed a probabilistic model of long COVID prevalence and disability due to long COVID using age, gender, race/ethnicity, vaccination status, and estimated infection dates of resident cases (Supplement C). Location data was used to aggregate prevalence by institution by day.

The effects of race/ethnicity and gender are those observed in community settings; it is not clear whether they may reflect social phenomena such as access to health care, or proximately physical phenomena such as pre-existing comorbidities. Because institutional factors such as access to health care may not occur in the same way in the prison setting, we modeled long COVID prevalence in two ways: first, assuming the effects of race/ethnicity and gender are as reported in community cases (“more stratified” long COVID model, hereafter), and second, assuming those effects do not exist in the prison setting (“less stratified”). The true distribution of long COVID in this setting may lie between these two extremes.

We conservatively took the proportion of long COVID cases that lead to disability at 19% (Ayoubkhani et al., 2021; Spiers, 2022), and used this proportion to produce an estimate of the number of disabling long COVID cases by location, age, race/ ethnicity, and gender. Because this estimate was based on the estimate of long COVID prevalence, it also had more and less stratified versions. Because our estimate of the proportion of long COVID cases that lead to disability was not stratified, we did not estimate the racial/ethnic distribution of disabling long COVID cases separately from that of long COVID overall.

To compare, we used the same model to estimate the prevalence of long COVID and of disability from long COVID in California over the same time period, in total and by race.

### 2.8. Data sharing

Our data sharing agreement with CDCR and CCHCS does not permit publication of the raw source data.

Data requests may be made to the California Correctional Health Care Services and are subject to controlled access, due to requirements to enhance protection of this vulnerable incarcerated population. Source code used in the analysis is available on request from the corresponding author.

## 3. Results

### 3.1. Summary statistics

The first positive SARS-CoV-2 test was administered on March 20, 2020 at the California State Prison in Los Angeles County (LAC). Between then and March 25, 2022, 196,652 individuals were incarcerated in CDCR facilities (Table 1). By the end of that time, 66,684, or 33.9% of them, had tested positive for SARS-CoV-2 while incarcerated in a CDCR facility.

**Table 1:** **Descriptive statistics** of prison residents, person-years, and residents infected (cases) during the study period, stratified by age, race/ethnicity, and gender (TGNBI=transgender, nonbinary, or intersex). “Infected residents” denotes the number of distinct individuals who became infected (note that elsewhere we use the term “Cases” for each infection event of a person, who may be infected multiple times). Ages are estimated from individuals’ birth years as follows: for total and cases, ages are calculated as of March 20, 2020, assuming a birthday of July 1; for person-years ages are calculated as of each day the person is resident in the system, assuming a birthday of July 1.

|  | **Residents**<br>**(N=196,652)** | **Person-Years**<br>**(N=206,904)** | **Infected Residents**<br>**(N=66,684)** |
| --- | --- | --- | --- |
| **Age** |  |  |  |
| <23 | 11,567 (5.9%) | 4,364 (2.1%) | 3,378 (5.1%) |
| 23–35 | 82,046 (42%) | 73,062 (35%) | 24,085 (36%) |
| 35–50 | 65,775 (33%) | 74,672 (36%) | 22,769 (34%) |
| >50 | 37,264 (19%) | 54,805 (26%) | 16,452 (25%) |
| **Race/Ethnicity** |  |  |  |
| Amer. Ind./Alaska Native | 2,306 (1.2%) | 2,485 (1.2%) | 866 (1.3%) |
| Asian/Pac. Isl. | 2,909 (1.5%) | 2,849 (1.4%) | 941 (1.4%) |
| Black | 51,796 (26%) | 59,620 (29%) | 17,096 (26%) |
| Latinx | 88,485 (45%) | 92,583 (45%) | 30,468 (46%) |
| White | 44,189 (22%) | 40,975 (20%) | 14,568 (22%) |
| Other | 6,967 (3.5%) | 8,393 (4.1%) | 2,745 (4.1%) |
| **Gender** |  |  |  |
| Cisgender Man | 185,248 (94%) | 198,572 (96%) | 63,845 (96%) |
| Cisgender Woman | 10,910 (5.5%) | 7,422 (3.6%) | 2,365 (3.5%) |
| TGNBI | 494 (0.25%) | 911 (0.44%) | 474 (0.71%) |

Counting reinfections, 73,386 cases were detected through March 25, 2022, for an average of 1.10 infections per ever-infected resident. By the 15th of October 2020, 15,499 cases were detected, involving 33 of the 35 CDCR institutions. Between that date and the end of March 2021 a much more intense wave of outbreaks swept the system, causing 34,189 cases among residents and involving all 35 institutions. A third wave in the winter of 2021–2022 brought an additional 23,698 cases, again including all 35 institutions. As of March 25, 2022, CDCR reported 253 residents had died as a result of COVID-19 infection (California Department of Corrections and Rehabilitation, n.d.b).

The largest number of cases detected on a single day at an institution was 494 cases, at Pleasant Valley State Prison (PVSP) on Nov. 30, 2020. The seven-day average of cases detected per day exceeded 100 on four occasions, at San Quentin (SQ) in June 2020 (March 20, 2020 population 4,013), Chuckawalla Valley State Prison (CVSP) in May 2020 (2,958), California Men’s Colony (CMC) in January 2021 (3,836), and Wasco State Prison (WSP) in January 2022 (4,564) (Figures E.1, E.2, Supplement E).

There were 174 outbreaks of size 3 or greater (Figure E.2, Supplement E), including every institution. 79 of them exceeded 100 cases, and 27 exceeded 1000 cases. 20 of the 35 institutions had outbreaks extending to 1/3 or more of their March 20, 2020 population, and 32 of the institutions had more than one outbreak exceeding 100 cases.

### 3.2. Transmission estimates

We performed a probabilistic reconstruction of the timing and location of individuals’ infection events and infectious periods, and used it to estimate effective reproduction numbers by individual for each day, which were then aggregated at the building and institution levels.

As mentioned, this fine-grained analysis was motivated by the need to account for movement of individuals between the times of infection and detection, in order to distinguish the effects of different locations and room types on transmission. Because residents were often moved into quarantine and isolation, and sometimes moved for preventative purposes, as outbreaks spread, case detections could potentially occur in different locations from where transmission occurred. Our estimation reconstructs the time of each individuals’ infection as a distribution of possible dates preceding the time of their first positive test, at which times they may have been located elsewhere from when they were tested.

In the period from March 20, 2020, to March 25, 2022, residents’ bed locations were moved from room to room within an institution 803,482 times and across institution boundaries 197,163 times (see Supplement G for details).

We examined the effect of movement and transfer on the apparent location of disease transmission events, we compared the number of cases detected by a positive test while housed in a room to the number of cases infected in the room, as estimated by our reconstruction of transmission dynamics. A linear regression with intercept of zero by room type indicated that infection events tended to exceed detection events in dorm rooms regardless of door type, and the opposite in cells regardless of door type (Supplement G). This may be due to infected residents being moved from dorm settings to smaller cells as a control measure, before detection. As a consequence, if the difference between place of infection and place of detection were not accounted for, reproduction numbers would tend to be overestimated in cells and underestimated in dorms; we have accounted for this effect by using infection dates.

We estimated daily effective reproduction numbers by building in each of the CDCR’s prisons, across the date range May 1, 2020 to March 19, 2021. This procedure also yielded daily probabilistic estimates of the number of cases infected each day (incidence) by building. These results are visualized in Figures I.1–I.35, in the Supplement, presenting the estimated incidence and daily *R* values by day and building in each institution.

We summarized average estimated reproduction numbers by day in the 35 CDCR institutions, providing a look at the overall course of the pandemic in the prisons (Figure 2). Outbreaks are characterized by reproduction numbers greater than one (red, yellow, and green in the figure) as the outbreak spreads, followed by reproduction numbers dropping below one when the outbreak reaches its peak and begins to shrink (blue in the figure). Thus the peak of each outbreak occurs around the time when the color shifts from red to blue at an institution, reading from left to right in the figure. *R* values in the range 1–4 are widespread, with a small number of peaks around 5. The plot shows all days on which estimated total infectiousness profile was at least 0.5 (see Supplement A.1 for details).

**Figure 2:**
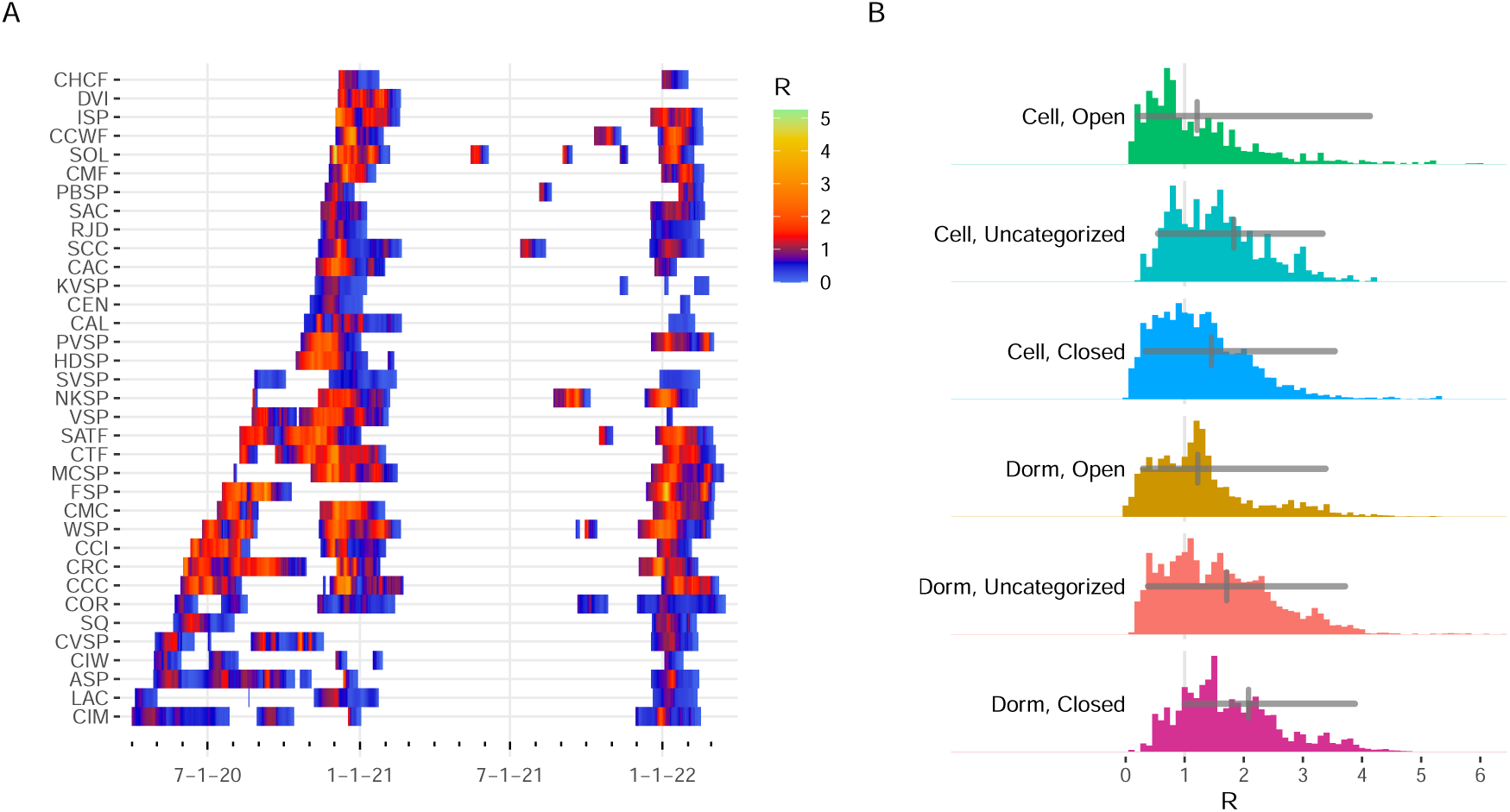
(A) Daily weighted average of **estimated** *R* **values by institution**. Detailed plots of daily *R* by building within each institution are included in the supplement. (B) Distribution of **estimated** *R* **values by room type**. Heavy lines mark the median value and 95% central interval.

Estimates of *R* were then summarized by the room type where each infectious individual was housed. The distribution of estimated *R* values across all infectious individuals in the CDCR system in the time span modeled displayed some difference between room types (Figure 2). Notably, the *R* values seen in celled housing were not systematically lower than those in dorm housing, a notable result given that celled housing had once been assumed safer and less conducive to transmission than dorms. Secondly, the median *R* was actually higher for individuals in cells with solid doors than those in cells with permeable doors, a notable result given the risks associated with cells with bars for doors like those at San Quentin.

To assess the seasonality of transmission in cells and dorms, we aggregated the inferred true incidence events over room types and seasons (Figure 3). All types of cells and dorms hosted significant numbers of transmission events in spring and summer as well as during the systemwide waves of outbreaks in fall and winter. In particular, hundreds of cases were infected in dorm rooms with solid doors in spring and summer months, implying that disuse of cold-weather heating systems does not make the use of these rooms an effective protection against transmission. A chart of seasonal transmission events by institution (Figure J.3C, Supplement J) identifies 8 institutions where over 100 cases were infected in closed cells over a spring and summer: CIM, CIW, CMC, COR, ISP, CCI, and CTF in 2020 and SOL in 2021. The average estimated *R* in closed cells exceeded one during spring-summer outbreaks at CIW, CMC, COR, CCI, CTF, and SOL (Figure J.3B, Supplement J).

**Figure 3:**
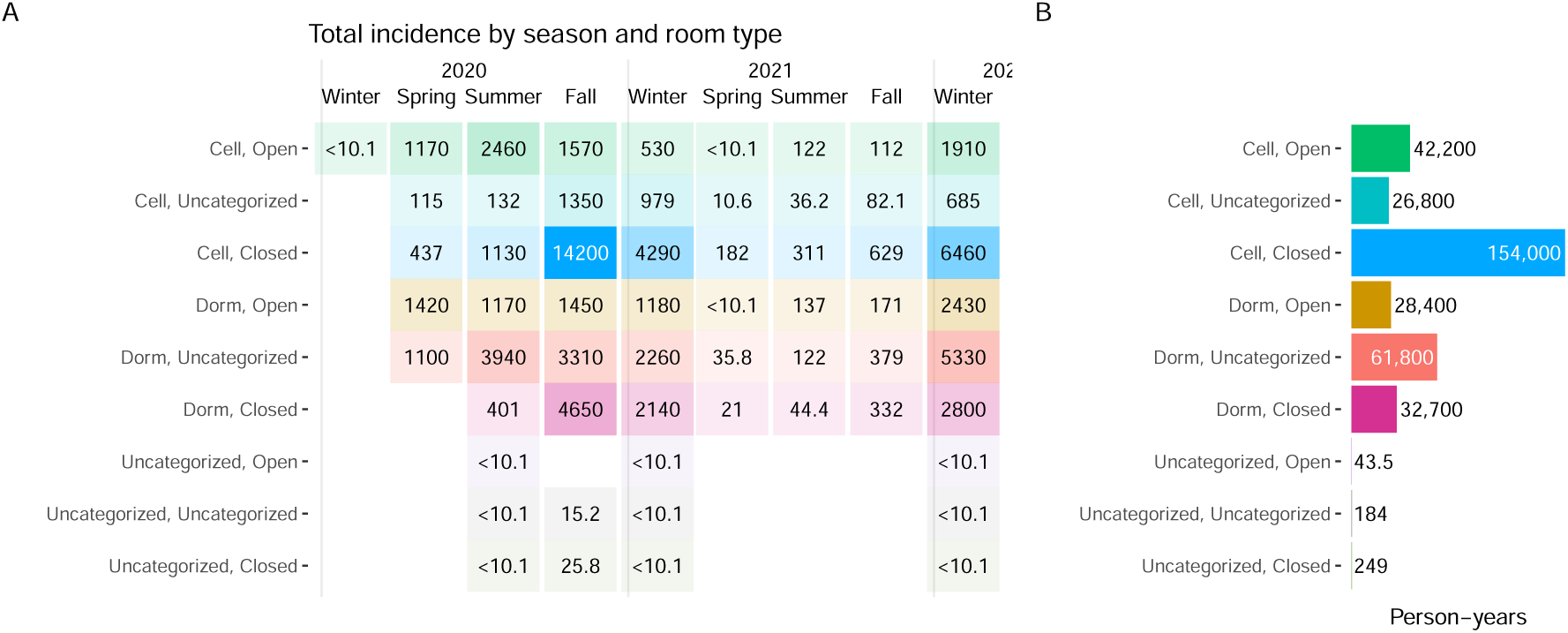
(A) Estimated **total incidence by room type** in each season, (B) **Total person-years per room type**.

### 3.3. Distribution of cases

The raw rate of SARS-CoV-2 infection events per person-year among prison residents was in the range 0.3–0.4 events per person-year for cisgender men, varying by race/ethnicity, while it was in the higher range 0.4–0.5 for cisgender women, and much higher at 0.8–1.0 for TGNBI residents (Figure F.2, Supplement F). There appeared to be some variation between intersectional combinations of gender and race/ethnicity beyond the rates associated with either separately.

Poisson regression analysis for interaction between race/ethnicity and gender, with corrections for clustering by building and for multiple comparisons, found the infection rates in all three gender classifications to be significantly different from the whole-population rate (Figure F.3, Supplement F). After age adjustment and inclusion of a random effect for each building, the rate was found to be highest for cisgender women, higher than average for TGNBI people, and lower than average for cisgender men. The rate for Black people was significantly lower than average, and for categories White and Other was significantly higher than average. Significant differences were found in two intersectional groups: a higher rate in Latinx cisgender men than predicted by their gender and race independently, and lower in American Indian or Alaska Native cisgender men than predicted.

### 3.4. Excess cases

While the waves of cases within the prisons generally co-occurred with surges of COVID cases in California as a whole, the incidence rates were substantially higher inside the prisons than in the state in many institutions during the first wave, and in most during the second wave (Figure 4A,C). While the third wave of prison outbreaks in Winter 2021– 2022 was large, it was exceeded by the massive surge in California cases at that time, when the Omicron variant emerged.

**Figure 4:**
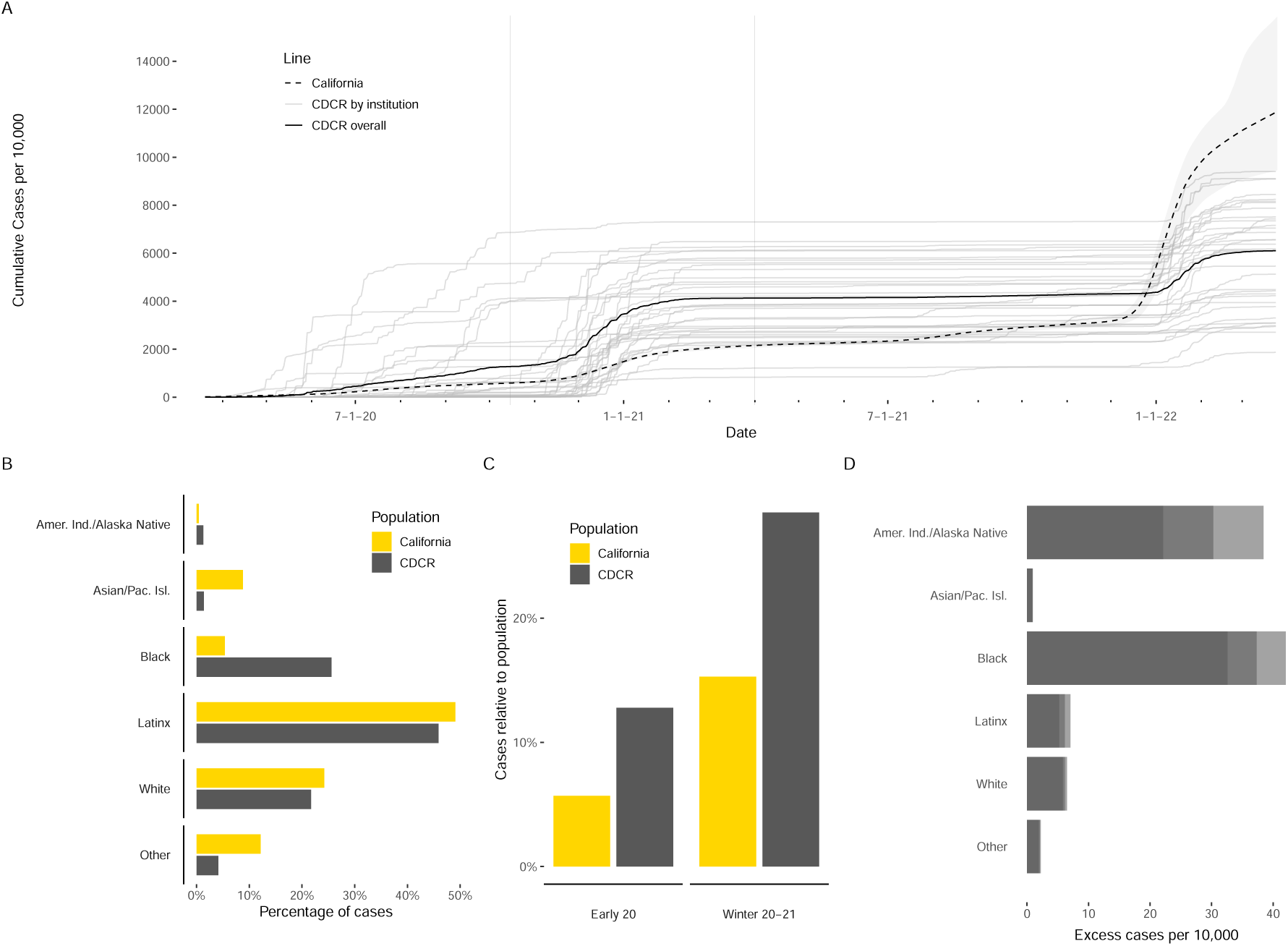
(A) **Cumulative incidence rate of COVID-19 infections** in each CDCR institution (gray), in CDCR overall (black), and in California (black, dashed, with 95% confidence interval), using IHME estimate of true incidence in California (including incarcerated populations). Vertical lines separate three “waves” of transmission. Excess cases in the CDCR system are seen in the first two waves. (B) **Racial/ethnic distribution of COVID-19 infections** in the CDCR system and estimated infections in California over all three waves. (C) **Attack rate** of outbreaks in the CDCR system and estimated attack rate in California, in first two waves. (D) **Estimated excess cases** in CDCR system relative to statewide population, by race/ethnicity, in the first two waves of outbreaks. Gradations of lightness on horizontal bars indicate mean and 95% confidence interval of estimate.

The state of California was 4.1% Black in 2021, and 0.2% was American Indian or Alaskan Native. The CDCR resident population, on the other hand, was 28.4% Black as of March 20, 2020 and 1.2% American Indian or Alaskan Native. Reflecting the makeup of the incarcerated population, a larger fraction of the COVID-19 cases in the CDCR system were Black and Indigenous people than in the state (Figure 4B): 25.6% of CDCR resident COVID-19 cases were Black, while 5.4% of cases in California were, and people classified as American Indian or Alaskan Native made up 1.3% of its cases, while comprising 0.4% of the state’s cases.

To validate this comparison, since differences in testing rates could bias these estimates, we compared testing rates in the CDCR system and in the state by race/ethnicity over the same time period. In the CDCR system, Black, American Indian/Alaska Native, and white residents were tested 13.6, 13.8, and 14.0 times per person-year on average, respectively. Statewide, Black Californians were tested at 1.15 times the rate of white Californians, while the rate for the American Indian classification was 0.83 the white rate. Thus no undercount of Black cases in the state relative to CDCR is indicated, and while Indigenous cases may have been relatively undercounted due to testing coverage in California, the difference is not sufficient to explain the factor of over 3 difference in percentage between the 0.4% of statewide cases and 1.3% of CDCR resident cases who were American Indian or Alaska Native.

We used the IHME estimate of the true count of COVID-19 cases in the state together with testing rates by race/ethnicity and age-specific incidence rates to estimate community rates of infection in the state stratified by race/ethnicity and age, and used that to estimate the number of CDCR residents who would have been infected during the first two waves of transmission if they were not incarcerated. This counterfactual count was 18,000 (95% CI: 15,800– 20,300) cases, while the true number of CDCR cases detected during that time was 49,688. As a result, we estimated that 31,700 (95% CI: 29,400–33,900) COVID-19 cases in excess of the counterfactual number occurred in CDCR outbreaks, at an overall rate of 6.23 (95% CI: 6.21–6.24) excess cases per 10,000 California residents. The excess case rate was far higher for Black prison residents than any others, at 37.3 (95% CI: 32.6–42.0) Black cases per 10,000, followed by 30.3 (95% CI: 22.1–38.4) American Indian and Alaska Native cases per 10,000, with the excess case rate for all other populations falling below 7 per 10,000 (Figure 4D).

### 3.5. Long COVID

Of the 66,684 CDCR residents reported infected with SARS-CoV-2 during the time period studied, our model estimated a total of 10,600 (95% CI: 10,400– 10,800) (more stratified; 9,240 (9,060–9,410) by less stratified model) ever having long COVID symptoms, and 2,010 (95% CI: 1,970–2,050) (resp. 1,760 (1,720–1,790)) ever having disabling long COVID symptoms. The estimated prevalence of long COVID peaked at around 5,600 (resp. 4,900) in early 2021, gradually declined by a fraction, and returned to about its peak level at the end of the period studied in January 2022 (Figure 5A). The estimated prevalence of disability from long COVID followed the same pattern, peaking near 1,100 (resp. 930) and remaining near its peak level (Figure 5B).

**Figure 5:**
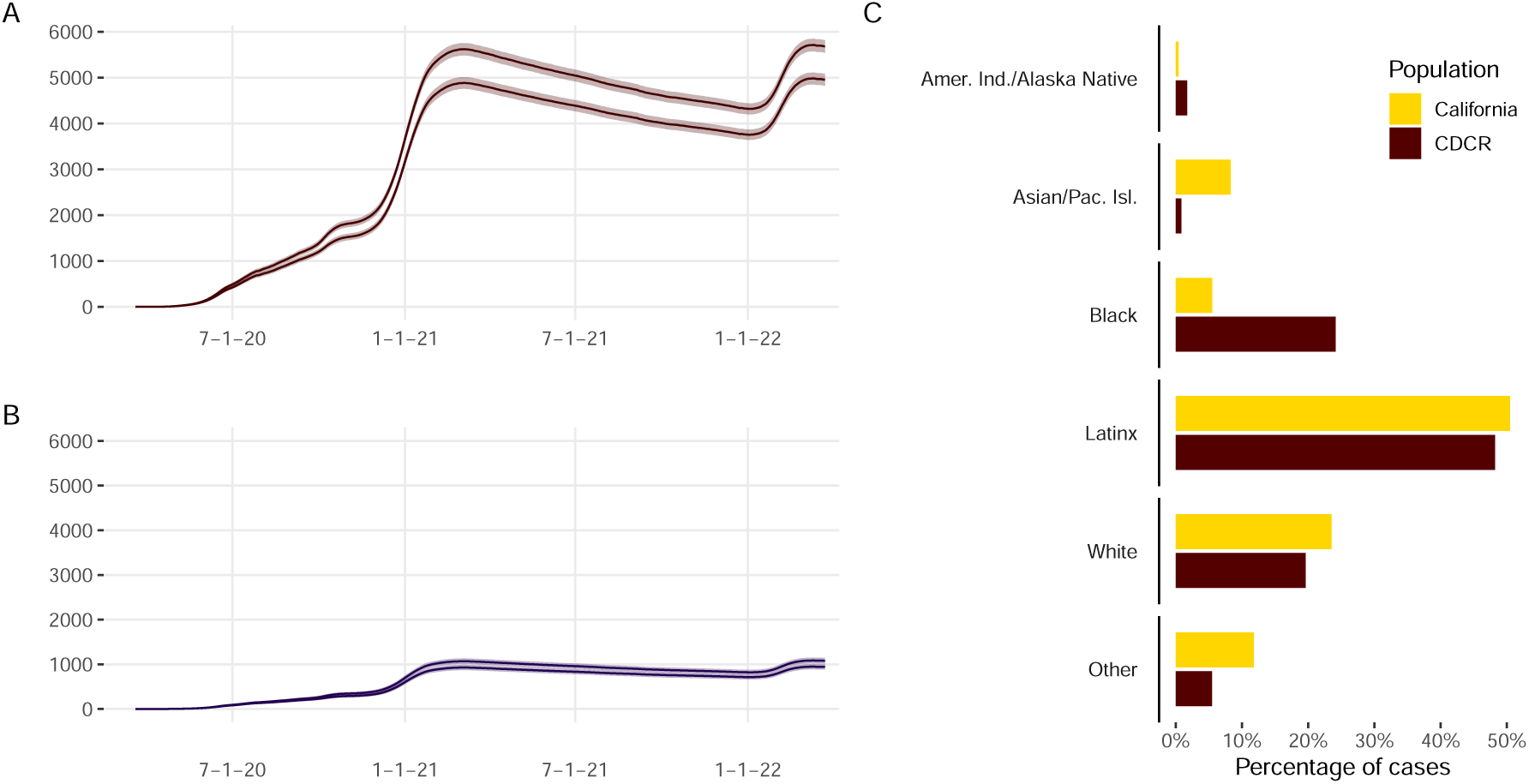
(A) Estimated **long COVID prevalence** in the CDCR system by date, in more (higher curve) and less (lower curve) stratified long COVID models, (B) estimated **prevalence of disability due to long COVID** in the CDCR system by date, in more (higher curve) and less (lower curve) stratified long COVID models, (C) estimated distribution of **ever having long COVID** through March 25, 2022, in the CDCR system and in California.

The estimated racial/ethnic composition of long COVID epidemiology in the prisons reflected the composition of the COVID-19 case burden (Figure 5C). As with acute COVID-19, Black people make up a much higher proportion of the estimated long COVID cases in the prisons than of the California population, at 24.1% (95% CI: 23.3%–25%) of the residents with long COVID in the more stratified model and 25.4% (24.5%–26.2%) in the less stratified one, in comparison to 5.5% (5.45%–5.56%) of estimated long COVID cases in California; 4.1% of California’s population is Black. Similarly, CDCR residents identified as American Indian or Alaskan Native make up 1.71% (95% CI: 1.49%–1.93%) of estimated long COVID cases under the more stratified model and 1.32% (95% CI: 1.13%–1.51%) under the less stratified one, while the American Indian or Alaska Native population makes up 0.432% (0.416%–0.447%) of estimated long COVID cases in California, and 0.2% of California’s population. See Supplement H for more detail.

## 4. Discussion

We have presented statistics and visualizations documenting the spread of COVID-19 in three waves of outbreaks in the California state prison system (CDCR) from the emergence of the pandemic through March 25, 2022, an estimate of the excess cases caused by the CDCR outbreaks, estimates of true incidence and effective reproduction numbers at all CDCR institutions and in the different types of rooms used, and an estimate of the extent and distribution of long COVID and consequent disabling symptoms resulting from these outbreaks.

This study has several limitations. Our inference of *R* and incidence used CDCR’s reported individual testing and symptom report data to identify cases, with the consequence that any infections that were not identified by positive tests or symptom reporting were not included. As a result, daily incidence and overall case numbers may have been underestimated (to the same degree as in any other reporting of CDCR’s case counts, including the CDCR public COVID dashboard). While reproduction number estimates are likely to be relatively robust to undercounting of cases at a consistent rate through time, since the ratio of new cases to existing cases is robust to that difference, if there were changes in the proportion of cases detected through time, or changes in the time from infection to detection, reproduction numbers may have been biased by those differences. There may have been an increase in case detection over time due to limited availability of testing early in the pandemic, which could cause an upward bias in *R* estimates; however, if the change was gradual, the impact on *R* estimation was likely minimal as the typical generation time from infection of one case to infection of secondary cases is only a week or less (e.g. Ferretti et al., 2020). For further discussion of these limitations see Supplement K. Transmission links may have been influenced by patterns not captured by the location and movement data used here, such as staff movements and resident work assignments. The estimation of excess cases, accounting for age and race/ethnicity, does not account for comorbidities, gender differences, or higher resolution, place-based matching to where counterfactually decarcerated individuals are likely to live, ability to shelter in place given employment, vaccination coverage, and other variables. Estimates of the extent and distribution of long COVID and its impacts are necessarily incomplete and more will be revealed as time passes.

Our estimates of average effective reproduction numbers over the infective population at an institution during this period up to January 2022 fell in the range between 0 and 5 (see for example Figure 2), in contrast to some early estimates of higher reproduction numbers in congregate settings (*e.g.* 8.44 in a jail (95% CI: 5.0–13.13) (Puglisi et al., 2020), up to 11.2 in a cruise ship (Mizumoto and Chowell, 2020)). While not reaching such extreme values, our estimates of effective reproduction numbers here did in some cases exceed many estimates of basic reproduction numbers in community transmission of the early dominant strains of SARS-CoV-2 (for example, 2.87 (95% CI, 2.39–3.44) (Billah et al., 2020)). We note that basic reproduction numbers (*R*_0_, the number of new cases generated by a case in the first moments of an outbreak before any individuals have lost susceptibility or control measures have begun) tend to be higher than effective reproduction numbers, and of course the extent and duration of community transmission attest to the danger posed by reproduction numbers in the range found there. We note also that because of smoothing induced by uncertainty in timing of transmission events (see Supplement K for details on this point) as well as averaging over rooms and individuals, our methods may have underestimated the upper extremes of true *R* values. The CDCR dataset used here is a rich one and would likely afford valuable further research into associations between reproduction numbers and time, control efforts, acquired immunity, duration of outbreaks, and other variables.

When transmission was widely thought to be droplet-based and physical distancing was one of the main interventions in use, early evidence suggested that housing incarcerated people in small cells can reduce transmission compared to dormitory housing (Hagan, 2020). In San Quentin, however, nearly all cases were housed in cells, but cells with bars and perforated metal for doors that opened onto a shared airspace that was not well-ventilated, suggesting that the benefits of cells can be undermined by the lack of a door to block airflow. Then, in mid-October 2020, large outbreaks appeared in multiple institutions in cells with solid walls and doors. In a visit to the Substance Abuse Treatment Facility (SATF) in Corcoran, one such institution, scientists observed that heating, ventilation, and air conditioning (HVAC) systems that are used in the fall and winter for heating were using filters that were not adequate to remove infected aerosols from the air when circulating air between cells, were delivering lower air exchange rates than the minimum recommended for infection control, and were inoperative in some cells (Sklar et al., 2023; Sears et al., 2021; Kwan et al., 2022). Because of the seasonal use of these heating systems, it remained unclear whether cells with solid walls and doors might be an effective tool for disease control in spring and summer months.

We found reproduction numbers well above the threshold of one in all types of housing, both celled and dormitory. Reproduction numbers do not appear to be overall reduced in celled housing compared to dorms, a meaningful finding given that cells have been thought to have some protective benefit due to the smaller room size. Further, we found that substantial numbers of residents of cells with solid doors were infected, not only in fall and winter when transmission can be ascribed to circulation of infectious aerosols by heating systems, but also during spring and summer months.

The control methods used to protect the residents of these crowded, unsafe buildings clearly failed. In the absence of evidence to the contrary, and given the nature of congregate settings and high transmissibility of the virus, we assess that any measures short of decarceration of overcrowded prisons are unlikely to be effective at preventing extensive disease spread during a ubiquitous airborne pandemic.

Age- and location-adjusted analysis of the associations of incidence with race/ethnicity and gender found substantial and significant effects of both race/ethnicity and gender as well as their interaction, with infection rates high among cisgender women and TGNBI residents overall, low among Black residents and high among white residents and those of “other” races overall, and higher in Latinx cisgender men and lower in Indigenous cisgender men than predicted by their race/ethnicity and gender taken independently. These results call for further study to understand and treat their causes.

Public health writing on racial disparities often references Ruth Wilson Gilmore’s formulation that racism, as an ongoing social institution, “is the state-sanctioned or extralegal production and exploitation of group-differentiated vulnerability to premature death.” While it has general application, this definition comes from Gilmore’s historical analysis of the growth and overfilling of the California state prison system (Gilmore, 2007). Both incarceration and health disparities are important cases of institutional racism by this definition.

SARS-CoV-2 infection rates in 2020 and winter 2020–21 were higher *per capita* in California’s prison system than in the rest of the state, producing an excess of cases compared to the numbers that would have occurred if all prison residents had been in the communities. Because of the extreme racial imbalance in incarceration, the excess case burden from these prison outbreaks is much higher on Black and Indigenous people than on others.

We estimate that on the order of five to six thousand residents in the CDCR system are likely to be affected by long-term effects of COVID-19 contracted during the period studied, hundreds of them with disabling impacts, and many of them with ongoing effects for years to come. The burden of long COVID and resulting disability is estimated to be in excess of community rates, and to fall disproportionately on Black and Indigenous residents, in the same way as with acute infections. We conclude that in the presence of the prisons’ COVID-19 outbreaks a larger proportion of the statewide burdens of illness, disability, and potential death fell on Black and Indigenous people than if those outbreaks had been smaller or had not occurred.

It follows that the spread of the SARS-CoV-2 virus in the prisons has itself emerged as a new manifestation of institutional racism in California, as the pandemic, together with the legal and management decisions that led to the large CDCR outbreaks of 2020–22, resulted in group-differentiated exposure to death and disability. The lasting impact of these outbreaks embodied by the hundreds of Black and Indigenous Californians living with long COVID symptoms as a result of these outbreaks, many with ongoing disability as a result, is a second, ongoing, new manifestation of institutional racism. A similar dynamic is likely to have occurred in other states and countries.

While the similarly large wave of prison outbreaks in winter 2021–22 did not exceed the very high incidence rates caused by the Omicron surge in the community, this fact does not diminish or negate the impact of the previous waves. The continuing transmission of the disease both in the prisons and in the community is still causing harm, and the harm done by earlier outbreaks is ongoing.

While our estimates of excess cases serve to identify sharp disparities in comparison to community rates of COVID-19 infection, which can be expected to exist in associated long COVID cases as well, we want to note that the comparison to community rates can obscure disparities that exist in the community setting as well as in the carceral one. In this case, Latinx residents make up a very large percentage of the imprisoned population, as well as in the state, and a large proportion of cases are Latinx in both populations. In fact, the rate of infection of Latinx people in California has been exceptionally high (see Supplement D). Given the overrepresentation of Latinx residents in the prisons—45% of the study population, versus 40% of the state population—it is reasonable to think that if Latinx infection rates in the state had been proportionate to the rest of the state, then the excess rate due to prison transmission would likely have been disproportionately high for Latinx as well as Black and Indigenous residents.

Between 1990 and 2009, CDCR was found in multiple court rulings to be providing inadequate medical care, inadequate medical screening of incoming prison residents, and untimely response to medical emergencies, as a result of overcrowding and underprovision of medical services. The U.S. Supreme Court upheld these rulings in 2011, found CDCR to be in violation of the Eighth Amendment to the U.S. Constitution, which prohibits cruel and unusual punishment, and ordered the population to be reduced (560 U.S. 493, 2011; Aviram, 2022). In 2020, as a result of the first San Quentin outbreak, the prison’s management was again found in violation of the Eighth Amendment, in “the worst epidemiological disaster in California correctional history,” by failing to heed recommendations to reduce crowding by decarcerating a portion of the population (270 Cal. Reptr. 3d. 140, 2020; McCoy et al., 2020; Aviram, 2022). While the later ruling awaits appeal, holding incarcerated individuals in crowded buildings with inadequate protections and care while a deadly disease spreads from room to room continues to be cruel and unusual, and will be cruel and wrong even if it becomes usual.

While COVID-19 outbreaks in California state prisons have become fewer since early 2022, they have continued (California Department of Corrections and Rehabilitation, n.d.a). Further outbreaks are likely, both of COVID-19, with possible loss of effectiveness of vaccination and natural immunity (Miller et al., 2023; Lewnard et al., 2023), and of other diseases, both old and new (Park et al., 2021), potentially aggravated by the immune system damage associated with long COVID (Davis et al., 2023; Turner et al., 2023).

We urge comprehensive medical care for all those suffering from ongoing effects of COVID-19 in the prisons, and decarceration to improve prevention of future outbreaks of the disease and of others that may emerge.

## Data Availability

Our data sharing agreement with CDCR and CCHCS does not permit publication of the raw source data.
Data requests may be made to the California Correctional Health Care Services and are subject to controlled access, due to requirements to enhance protection of this vulnerable incarcerated population.
Source code used in the analysis is available on request from the corresponding author.

## Acknowledgements

We are grateful to Brie Williams, Stefano Bertozzi, Kirsten Bibbins-Domingo, Heidi Bauer, Sarah Ackley, Jonathan Dushoff, and Todd Parsons for invaluable feedback and support, and to David Leidner of CDCR for development and maintenance of the data set, including the door types. Special appreciation to Traci Schlesinger and the many others lost in 2020–2022, and to Em “formerly Mike” Sumner. who helped us with sociology questions after we lost Traci. LW and TCP were supported by NIH GM130900. SB and TCP were supported by CDC U01CK000590, as part of the Modeling Infectious Diseases in Healthcare Network. SB was also supported by NIH K12 EY031372. LW, HA, AK, and DS received funding from the California Prison Receivership Office.

## Footnotes

1 Deuel Vocational Institution (DVI) was closed in 2021, and California Correctional Center (CCC) was closed in 2023.

## References

270 Cal. Reptr. 3d. 140, 2020. In re von Staich, 2020.

560 U.S. 493, 2011. Brown v. Plata, 2011. URL https://supreme.justia.com/cases/federal/us/563/493/.

Ziyad Al-Aly, Benjamin Bowe, and Yan Xie. Long COVID after breakthrough SARS-CoV-2 infection. Nature Medicine, 28(7):1461–1467, July 2022. ISSN 1546-170X. doi: 10.1038/s41591-022-01840-0. URL https://www.nature.com/articles/s41591-022-01840-0. Number: 7 Publisher: Nature Publishing Group.

Michelle Alexander. The new Jim Crow: Mass incarceration in the age of colorblindness. The New Press, 2020. ISBN 1-62097-194-1.

Hadar Aviram. The House Always Wins: Doctrine and Animus in California’s COVID-19 Prison Litigation. Case Western Reserve Law Review, 72(3):565– 630, 2022. URL https://scholarlycommons.law.case.edu/caselrev/vol72/iss3/5.

Daniel Ayoubkhani, Sasha King, and Matt Bosworth. Prevalence of Ongoing Symptoms Following Coronavirus (COVID-19) Infection in the UK: 2 December 2021. Technical report, Office of National Statistics (UK), 2021. URL https://tinyurl.com/4wbnxtrn.

Daniel Ayoubkhani, Matthew L Bosworth, Sasha King, Koen B Pouwels, Myer Glickman, Vahé Nafilyan, Francesco Zaccardi, Kamlesh Khunti, Nisreen A Alwan, and A Sarah Walker. Risk of Long COVID in People Infected With Severe Acute Respiratory Syndrome Coronavirus 2 After 2 Doses of a Coronavirus Disease 2019 Vaccine: Community-Based, Matched Cohort Study. Open Forum Infectious Diseases, 9(9):ofac464, September 2022. ISSN 2328-8957. doi: 10.1093/ofid/ofac464. URL https://doi.org/10.1093/ofid/ofac464.

Aranka V. Ballering, Sander K. R. van Zon, Tim C. olde Hartman, and Judith G. M. Rosmalen. Persistence of somatic symptoms after COVID-19 in the Netherlands: an observational cohort study. The Lancet, 400(10350):452–461, August 2022. ISSN 0140-6736, 1474-547X. doi: 10.1016/S0140-6736(22)01214-4. URL https://www.thelancet.com/journals/lancet/article/PIIS0140-6736(22)01214-4/fulltext?ref=pmp-magazine. Publisher: Elsevier.

Elizabeth Barnert, Cyrus Ahalt, and Brie Williams. Prisons: Amplifiers of the COVID-19 Pandemic Hiding in Plain Sight. American Journal of Public Health, 110(7):964– 966, July 2020. ISSN 0090-0036. doi: 10.2105/AJPH. 2020.305713. URL https://ajph.aphapublications.org/doi/full/10.2105/AJPH.2020.305713. Publisher: American Public Health Association.

Md. Arif Billah, Md. Mamun Miah, and Md. Nuruzzaman Khan. Reproductive number of coronavirus: A systematic review and meta-analysis based on global level evidence. PLOS ONE, 15(11):1–17, 11 2020. doi: 10.1371/journal.pone.0242128. URL https://doi.org/10.1371/journal.pone.0242128.

Elizabeth A. Bradshaw. Do Prisoners’ Lives Matter? Examining the Intersection of Punitive Policies, Racial Disparities and COVID-19 as State Organized Race Crime. State Crime Journal, 10:16–44, April 2021. ISSN 2046-6056. doi: 10.13169/statecrime.10.1.0016. URL https://www.scienceopen.com/hosted-document?doi=10.13169/statecrime.10.1.0016. Publisher: Pluto Journals.

State of California. Variants. https://covid19.ca.gov/variants/. Accessed February 15, 2022. Accessed February 15, 2022.

California Department of Corrections and Rehabilitation. Population COVID-19 Tracking. https://www.cdcr.ca.gov/covid19/population-status-tracking/, Accessed April 3, 2023, n.d.a.

California Department of Corrections and Rehabilitation. CDCR Population COVID-19 Tracking, n.d.b. Retrieved July 24, 2024 from https://data.ca.gov/dataset/cdcr-population-covid-19-tracking/resource/5a3f496d-04be-4405-aea0-e83e26076efc.

California Department of Justice, Office of Justice Programs. The california department of justice’s review of immigration detention in california. Technical report, 2019. URL https://oag.ca.gov/sites/all/files/agweb/pdfs/publications/immigration-detention-2019.pdf.

California Department of Public Health. Outbreak Definition and Reporting Guidance, n.d. URL https://www.cdph.ca.gov/Programs/CID/DCDC/Pages/COVID-19/OutbreakDefinitionandReportingGuidance.aspx. https://www.cdph.ca.gov/Programs/CID/DCDC/Pages/COVID-19/OutbreakDefinitionandReportingGuidance.aspx. Accessed Dec. 14, 2021.

California Health and Human Services. COVID-19 Time-Series Metrics by County and State - California Health and Human Services Open Data Portal, 2023. URL https://data.chhs.ca.gov/dataset/covid-19-time-series-metrics-by-county-and-state.

California Health and Human Services Open Data Portal. Statewide covid-19 cases deaths demographics. https://data.chhs.ca.gov/dataset/covid-19-time-series-metrics-by-county-and-state/resource/e2c6a86b-d269-4ce1-b484-570353265183. Accessed 2024-01-08, December 2023. URL https://data.chhs.ca.gov/dataset/covid-19-time-series-metrics-by-county-and-state/resource/e2c6a86b-d269-4ce1-b484-570353265183.

E. Ann Carson and Rich Kluckow. Prisoners in 2022 – statistical tables. Technical report, U.S. Department of Justice, Office of Justice Programs, Bureau of Justice Statistics, 2022. URL https://bjs.ojp.gov/document/p22st.pdf.

Simon Cauchemez, Pierre-Yves Böelle, Christl A. Donnelly, Neil M Ferguson, Guy Thomas, Gabriel M. Leung, Anthony J Hedley, Roy M. Anderson, and Alain-Jacques Valleron. Real-time Estimates in Early Detection of SARS. Emerging Infectious Diseases, 12(1):110– 113, January 2006. ISSN 1080-6040. doi: 10.3201/eid1201.050593. URL https://www.ncbi.nlm.nih.gov/pmc/articles/PMC3293464/.

CDC. Long COVID - Household Pulse Survey - COVID-19, April 2023. URL https://www.cdc.gov/nchs/covid19/pulse/long-covid.htm.

Elizabeth T. Chin, David Leidner, Theresa Ryckman, Yiran E. Liu, Lea Prince, Fernando Alarid-Escudero, Jason R. Andrews, Joshua A. Salomon, Jeremy D. Goldhaber-Fiebert, and David M. Studdert. Covid-19 Vaccine Acceptance in California State Prisons. New England Journal of Medicine, 385(4):374–376, July 2021. ISSN 0028-4793. doi: 10.1056/NEJMc2105282. URL https://doi.org/10.1056/NEJMc2105282. Publisher: Massachusetts Medical Society eprint: https://doi.org/10.1056/NEJMc2105282.

Anne Cori, Neil M. Ferguson, Christophe Fraser, and Simon Cauchemez. A New Framework and Software to Estimate Time-Varying Reproduction Numbers During Epidemics. American Journal of Epidemiology, 178(9):1505– 1512, November 2013. ISSN 0002-9262. doi: 10.1093/aje/kwt133. URL https://www.ncbi.nlm.nih.gov/pmc/ articles/PMC3816335/.

Hannah E. Davis, Lisa McCorkell, Julia Moore Vogel, and Eric J. Topol. Long COVID: major findings, mechanisms and recommendations. Nature Reviews Microbiology, 21(3):133–146, March 2023. ISSN 1740-1534. doi: 10.1038/s41579-022-00846-2. URL https://www.nature.com/articles/s41579-022-00846-2. Number: 3 Publisher: Nature Publishing Group.

Matthew G. T. Denney and Ramon Garibaldo Valdez. Compounding Racialized Vulnerability: COVID-19 in Prisons, Jails, and Migrant Detention Centers. Journal of Health Politics, Policy and Law, 46(5):861–887, October 2021. ISSN 1527-1927. doi: 10.1215/03616878-9156019.

Sharon Dolovich and Aaron Littman. Ucla law covid-19 behind bars data project. UCLA School of Law, 2020.

Catherine Duarte, Drew B. Cameron, Ada T. Kwan, Stefano M. Bertozzi, Brie A. Williams, and Sandra I. McCoy. COVID-19 outbreak in a state prison: a case study on the implementation of key public health recommendations for containment and prevention. BMC public health, 22(1):977, May 2022. ISSN 1471-2458. doi: 10.1186/s12889-022-12997-1.

Fŕedéric Dutheil, Jean-Baptiste Bouillon-Minois, and Mäelys Clinchamps. COVID-19: a prison-breaker? Canadian Journal of Public Health, 111(4):480–481, August 2020. ISSN 1920-7476. doi: 10.17269/s41997-020-00359-6. URL https://doi.org/10.17269/s41997-020-00359-6.

Elóısa Díaz-Franćes and Francisco J. Rubio. On the existence of a normal approximation to the distribution of the ratio of two independent normal random variables. Statistical Papers, 54(2):309–323, May 2013. ISSN 1613-9798. doi: 10.1007/s00362-012-0429-2. URL https://doi.org/10.1007/s00362-012-0429-2.

FAIR Health. Patients diagnosed with post-COVID conditions: an analysis of private healthcare claims using the official ICD-10 diagnostic code. Technical report, 2022. URL https://collections.nlm.nih.gov/catalog/nlm:nlmuid-9918504887106676-pdf.

Luca Ferretti, Alice Ledda, Chris Wymant, Lele Zhao, Virginia Ledda, Lucie Abeler-Dörner, Michelle Kendall, Anel Nurtay, Hao-Yuan Cheng, Ta-Chou Ng, Hsien-Ho Lin, Rob Hinch, Joanna Masel, A. Marm Kilpatrick, and Christophe Fraser. The Timing of COVID-19 Transmission. SSRN Scholarly Paper ID 3716879, Social Science Research Network, Rochester, NY, October 2020. URL https://papers.ssrn.com/abstract=3716879.

Anna Flagg and Joseph Neff. Why Jails Are So Important in the Fight Against Coronavirus, March 2020. URL https://www.themarshallproject.org/2020/03/31/why-jails-are-so-important-in-the-fight-against-coronavirus Section: Coronavirus.

Christophe Fraser. Estimating individual and household reproduction numbers in an emerging epidemic. PloS One, 2(8):e758, August 2007. ISSN 1932-6203. doi: 10.1371/journal.pone.0000758.

Ruth Wilson Gilmore. Golden Gulag: Prisons, Surplus, Crisis, and Opposition in Globalizing California. University of California Press, 2007. ISBN 978-0-520-24201-2.

Katelyn M. Gostic, Lauren McGough, Edward B. Baskerville, Sam Abbott, Keya Joshi, Christine Tedijanto, Rebecca Kahn, Rene Niehus, James A. Hay, Pablo M. De Salazar, Joel Hellewell, Sophie Meakin, James D. Munday, Nikos I. Bosse, Katharine Sherrat, Robin N. Thompson, Laura F. White, Jana S. Huisman, Jéŕemie Scire, Sebastian Bonhoeffer, Tanja Stadler, Jacco Wallinga, Sebastian Funk, Marc Lipsitch, and Sarah Cobey. Practical considerations for measuring the effective reproductive number, Rt. PLOS Computational Biology, 16(12):e1008409, December 2020. ISSN 1553-7358. doi: 10.1371/journal.pcbi.1008409. URL https://journals.plos.org/ploscompbiol/article?id=10.1371/journal.pcbi.1008409. Publisher: Public Library of Science.

Liesl M. Hagan. Mass Testing for SARS-CoV-2 in 16 Prisons and Jails — Six Jurisdictions, United States, April–May 2020. MMWR. Morbidity and Mortality Weekly Report, 69, 2020. ISSN 0149-21951545-861X. doi: 10.15585/mmwr.mm6933a3. URL https://www.cdc.gov/mmwr/volumes/69/wr/mm6933a3.htm.

Joseph Hayes, Justin Goss, Heather Harris, and Alexandria Gumbs. California’s Prison Population. https://www.ppic.org/publication/californias-prison-population/, July 2022. Public Policy Institute of California fact sheet. Accessed June 20, 2023.

Tanya Albert Henry. 5 reasons why religious services pose high risk of COVID-19 spread. https://www.ama-assn.org/delivering-care/public-health/5-reasons-why-religious-services-pose-high-risk-covid-19-spread accessed June 20, 2023, December 2020.

Don Hummer. United States Bureau of Prisons’ Response to the COVID-19 Pandemic. Victims & Offenders, 15 (7-8):1262–1276, October 2020. ISSN 1556-4886. doi: 10.1080/15564886.2020.1829765. URL https://doi.org/10.1080/15564886.2020.1829765. Publisher: Routledge eprint: https://doi.org/10.1080/15564886.2020.1829765.

IHME COVID-19 Forecasting Team. Modeling COVID-19 scenarios for the United States. Nature Medicine, 27(1):94–105, 2021. ISSN 1078-8956. doi: 10.1038/s41591-020-1132-9. URL https://www.ncbi.nlm.nih.gov/pmc/articles/PMC7806509/.

Institute for Health Metrics and Evaluation. COVID-19 projections. Online at https://covid19.healthdata.org/united-states-of-america/california. Accessed 2024-01-21, December 2022.

Allison James, Lesli Eagle, Cassandra Phillips, D. Stephen Hedges, Cathie Bodenhamer, Robin Brown, J. Gary Wheeler, and Hannah Kirking. High COVID-19 Attack Rate Among Attendees at Events at a Church — Arkansas, March 2020. MMWR. Morbidity and why-jails-are-so-important-in-the-fight-against-coronavirMuso.rtality Weekly Report, 69(20):632–635, May 2020. ISSN 0149-2195, 1545-861X. doi: 10.15585/mmwr.mm6920e2. URL http://www.cdc.gov/mmwr/volumes/69/wr/mm6920e2.htm?s_cid=mm6920e2_w.

Lauren Jansen. Investigation of a SARS-CoV-2 B.1.1.529 (Omicron) Variant Cluster — Nebraska, November–December 2021. MMWR. Morbidity and Mortality Weekly Report, 70, 2021. ISSN 0149-21951545-861X. doi: 10.15585/mmwr.mm705152e3. URL https://www.cdc.gov/mmwr/volumes/70/wr/mm705152e3.htm.

Leslie Jones and Olivia Tulloch. COVID-19: Why Are Prisons a Particular Risk, and What Can Be Done to Mitigate this? May 2020. URL https://opendocs.ids.ac.uk/opendocs/handle/20.500.12413/15285. Accepted: 2020-05-07T08:24:25Z Publisher: SSHAP.

Anthea L. Katelaris, Jessica Wells, Penelope Clark, Sophie Norton, Rebecca Rockett, Alicia Arnott, Vitali Sintchenko, Stephen Corbett, and Shopna K. Bag. Epidemiologic Evidence for Airborne Transmission of SARS-CoV-2 during Church Singing, Australia, 2020 - Volume 27, Number 6—June 2021 - Emerging Infectious Diseases journal - CDC. doi: 10.3201/eid2706.210465. URL https://wwwnc.cdc.gov/eid/article/27/6/21-0465_article.

Stuart A. Kinner, Jesse T. Young, Kathryn Snow, Louise Southalan, Daniel Lopez-Acuña, Carina Ferreira-Borges, and Éamonn O’Moore. Prisons and custodial settings are part of a comprehensive response to COVID-19. The Lancet Public Health, 5(4):e188–e189, April 2020. ISSN 2468-2667. doi: 10.1016/S2468-2667(20)30058-X. URL https://www.thelancet.com/journals/lanpub/article/PIIS2468-2667(20)30058-X/fulltext. Publisher: Elsevier.

Brennan Klein, C. Brandon Ogbunugafor, Benjamin J. Schafer, Zarana Bhadricha, Preeti Kori, Jim Sheldon, Nitish Kaza, Arush Sharma, Emily A. Wang, Tina Eliassi- Rad, Samuel V. Scarpino, and Elizabeth Hinton. COVID-19 amplified racial disparities in the US criminal legal system. Nature, 617(7960):344–350, May 2023. ISSN 1476-4687. doi: 10.1038/s41586-023-05980-2. URL https://www.nature.com/articles/s41586-023-05980-2. Number: 7960 Publisher: Nature Publishing Group.

Shiloh Krupar and Amina Sadural. COVID “death pits”: US nursing homes, racial capitalism, and the urgency of antiracist eldercare. Environment and Planning C: Politics and Space, 40(5):1106–1129, August 2022. ISSN 2399-6544. doi: 10.1177/23996544211057677. URL https://doi.org/10.1177/23996544211057677. Publisher: SAGE Publications Ltd STM.

Ada Kwan, David Sears, Stefano M. Bertozzi, and Brie Williams, editors. California State Prisons During the COVID-19 Pandemic: A Report by the CalPROTECT Project. 2022. https://amend.us/wp-content/uploads/2022/05/2022-0501-CalPROTECT-Report.pdf.

Joseph A. Lewnard, Vennis Hong, Jeniffer S. Kim, Sally F. Shaw, Bruno Lewin, Harpreet Takhar, Marc Lipsitch, and Sara Y. Tartof. Increased vaccine sensitivity of an emerging SARS-CoV-2 variant. Nature Communications, 14(1):3854, June 2023. ISSN 2041-1723. doi: 10.1038/s41467-023-39567-2. URL https://www.nature.com/articles/s41467-023-39567-2. Number: 1 Pub-lisher: Nature Publishing Group.

Eric T. Lofgren, Kristian Lum, Aaron Horowitz, Brooke Mabubuonwu, Kellen Meyers, and Nina H. Fefferman. Carceral Amplification of COVID-19: Impacts for Community, Corrections Officer, and Incarcerated Population Risks. Epidemiology, 33(4):480, July 2022. ISSN 1044-3983. doi: 10.1097/EDE.0000000000001476. URL https://journals.lww.com/epidem/Fulltext/2022/07000/Carceral_Amplification_of_COVID_19__Impacts_for.6.aspx.

Los Angeles County Sheriff’s Department. Custody division quarterly report, april–june 2022. Technical report, 2022. URL https://lasd.org/wp-content/uploads/2022/09/Transparency_Custody_Division_Population_2022_Second_Quarter_Report.pdf.

Mattia Manica, Piero Poletti, Silvia Deandrea, Giansanto Mosconi, Cinzia Ancarani, Silvia Lodola, Giorgio Guzzetta, Valeria d’Andrea, Valentina Marziano, Agnese Zardini, Filippo Trentini, Anna Odone, Marcello Tirani, Marco Ajelli, and Stefano Merler. Estimating SARS-CoV-2 transmission in educational settings: A retrospective cohort study. Influenza and Other Respiratory Viruses, 17(1):e13049, 2023. ISSN 1750-2659. doi: 10.1111/irv.13049. URL https://onlinelibrary.wiley.com/doi/abs/10.1111/irv.13049. eprint: https://onlinelibrary.wiley.com/doi/pdf/10.1111/irv.13049.

Conor McAloon, Áine Collins, Kevin Hunt, Ann Barber, Andrew W Byrne, Francis Butler, Miriam Casey, John Griffin, Elizabeth Lane, David McEvoy, Patrick Wall, Martin Green, Luke O’Grady, and Simon J More. Incubation period of COVID-19: a rapid systematic review and metaanalysis of observational research. BMJ Open, 10(8), 2020. ISSN 2044-6055. doi: 10.1136/bmjopen-2020-039652. URL https://bmjopen.bmj.com/content/10/8/e039652. Publisher: British Medical Journal Publishing Group eprint: https://bmjopen.bmj.com/content/10/8/e039652.full.pdf.

S McCoy, SM Bertozzi, D Sears, A Kwan, C Duarte, D Cameron, and B Williams. Urgent memo: COVID-19 outbreak: San Quentin prison. San Francisco, CA, 2020. URL https://amend.us/wp-content/uploads/2020/06/COVID19-Outbreak-SQ-Prison-6.15.2020.pdf.

Jessica Miller, Nicole P. Hachmann, Ai-ris Y. Collier, Ninaad Lasrado, Camille R. Mazurek, Robert C. Patio, Olivia Powers, Nehalee Surve, James Theiler, Bette Korber, and Dan H. Barouch. Substantial Neutralization Escape by SARS-CoV-2 Omicron Variants BQ.1.1 and XBB.1. New England Journal of Medicine, 388 (7):662–664, February 2023. ISSN 0028-4793. doi: 10.1056/NEJMc2214314. URL https://doi.org/10.1056/NEJMc2214314. Publisher: Massachusetts Medical Society eprint: https://doi.org/10.1056/NEJMc2214314.

Kenji Mizumoto and Gerardo Chowell. Transmission potential of the novel coronavirus (COVID-19) onboard the diamond Princess Cruises Ship, 2020. Infectious Disease Modelling, 5:264–270, January 2020. ISSN 2468-0427. doi: 10.1016/j.idm.2020.02.003. URL http://www.sciencedirect.com/science/article/ pii/S2468042720300063.

Philip J. Murphy. Unlocking the Means of COVID-19 Spread From Prisons to Outside Populations. American Journal of Public Health, 111(8):1392–1394, August 2021. ISSN 0090-0036. doi: 10.2105/AJPH.2021.306401. URL https://ajph.aphapublications.org/doi/ abs/10.2105/AJPH.2021.306401. Publisher: American Public Health Association.

Marianne Napoles. Men’s prison in Chino is no longer a reception center, August 2020. URL https://www.championnewspapers.com/news/article_2d0d47a0-e3d6-11ea-bee3-237a46123af4.html.

Kathryn M. Nowotny, Zinzi Bailey, and Lauren Brinkley-Rubinstein. The Contribution of Prisons and Jails to US Racial Disparities During COVID-19. American Journal of Public Health, 111(2):197–199, February 2021. ISSN 0090-0036. doi: 10.2105/AJPH.2020. 306040. URL https://ajph.aphapublications.org/doi/abs/10.2105/AJPH.2020.306040. Publisher: American Public Health Association.

Katie Park, Keri Blakinger, and Claudia Lauer. A Half-Million People Got COVID-19 in Prison. Are Officials Ready for the Next Pandemic?, June 2021. URL https://www.themarshallproject.org/2021/06/30/a-half-million-people-got-covid-19-in-prison-are-officials-read Section: Coronavirus.

Todd L. Parsons and Lee Worden. Assessing the risk of cascading COVID-19 outbreaks from prison-to-prison transfers. Epidemics, 37:100532, December 2021. ISSN 1878-0067. doi: 10.1016/j.epidem.2021.100532.

Prison Policy Initiative. California profile, n.d. URL https://www.prisonpolicy.org/profiles/CA.html.

Wikramaratna PS, Paton RS, Ghafari M, and Loureņco J. Estimating the false-negative test probability of SARS-CoV-2 by RT-PCR. Euro Surveill, 25(50):2000568, 2020.

Lisa B. Puglisi, Giovanni S. P. Malloy, Tyler D. Harvey, Margaret L. Brandeau, and Emily A. Wang. Estimation of COVID-19 Basic Reproduction Ratio in a Large Urban Jail in the United States. Annals of Epidemiology, September 2020. ISSN 1047-2797. doi: 10.1016/j.annepidem.2020.09.002. URL http://www.sciencedirect.com/science/article/pii/S1047279720303471.

Eric Reinhart and Daniel L. Chen. Incarceration And Its Disseminations: COVID-19 Pandemic Lessons From Chicago’s Cook County Jail. Health Affairs, 39(8):1412–1418, August 2020. ISSN 0278-2715. doi: 10.1377/hlthaff.2020.00652. URL https://www.healthaffairs.org/doi/10.1377/hlthaff.2020.00652. Publisher: Health Affairs.

Eric Reinhart and Daniel L. Chen. Association of Jail Decarceration and Anticontagion Policies With COVID-19 Case Growth Rates in US Counties. JAMA network open, 4(9):e2123405, September 2021a. ISSN 2574-3805. doi: 10.1001/jamanetworkopen.2021.23405.

Eric Reinhart and Daniel L. Chen. Carceral-community epidemiology, structural racism, and COVID-19 disparities. Proceedings of the National Academy of Sciences, 118(21):e2026577118, May 2021b. doi: 10.1073/pnas.2026577118. URL https://www.pnas.org/doi/full/10.1073/pnas.2026577118. Publisher: Proceedings of the National Academy of Sciences.

Brendan Saloner, Kalind Parish, Julie A. Ward, Grace Di-Laura, and Sharon Dolovich. COVID-19 Cases and Deaths in Federal and State Prisons. JAMA, 324(6):602–603, August 2020. ISSN 0098-7484. doi: 10.1001/jama.2020.12528. URL https://doi.org/10.1001/jama.2020.12528.

Traci Schlesinger. Equality at the Price of Justice. NWSA Journal, 20(2):27–47, 2008. ISSN 2151-7371. URL https://muse.jhu.edu/pub/1/article/246753. Publisher: Johns Hopkins University Press.

Traci Schlesinger. Mass Incarceration, COVID-19, and Race as Exposure to Early Death. Vincentian Heritage Journal, 36(2), January 2022. ISSN 0277-2205. URL https://via.library.depaul.edu/vhj/vol36/iss2/11.

David Sears, Stefano Bertozzi, Rachel Sklar, Brittany Imwalle, Ada T. Kwan, Sandra I. McCoy, Robert C. Schell, Helena Archer, Chakriya Srey, and Brie Williams. Substance Abuse Treatment Facility (SATF) Site Visit Report. Technical report, March 2021. URL https://amend.us/wp-content/uploads/2021/03/SATF-December-2020-Visit-Report_3.7.21.pdf.

Kaitlyn M. Sims, Jeremy Foltz, and Marin Elisabeth Skidmore. Prisons and COVID-19 Spread in the United States. American Journal of Public Health, 111(8):1534–1541, August 2021. ISSN 0090-0036. doi: 10.2105/AJPH.2021. 306352. URL https://ajph.aphapublications.org/doi/full/10.2105/AJPH.2021.306352. Publisher: American Public Health Association.

Rachel Sklar, Elizabeth Noth, Ada Kwan, David Sears, and Stefano Bertozzi. Ventilation conditions during COVID-19 outbreaks in six California state carceral institutions. PLOS ONE, 18(11):e0293533, November 2023. ISSN 1932–6203. doi: 10.1371/journal.pone.0293533. URL https://journals.plos.org/plosone/article?id=10.1371/journal.pone.0293533. Publisher: Public Library of Science.

Jamie Solis, Carlos Franco-Paredes, Andŕes F. Henao-Martínez, Martin Krsak, and Shanta M. Zimmer. Structural Vulnerability in the U.S. Revealed in Three Waves of COVID-19. The American Journal of Tropical Medicine and Hygiene, 103(1):25–27, July 2020. ISSN 0002-9637, 1476-1645. doi: 10.4269/ajtmh.20-0391. URL https://ajtmh.org/doi/10.4269/ajtmh.20-0391.

Anabel Sosa. More California prisoners are requesting gender-affirming health care, including surgeries. Technical report, CalMatters, June 2023. URL https://calmatters.org/justice/2023/06/gender-affirming-care-california-prisons/.

Nicola Spiers. Recognising and bearing the burden of long COVID-related disability. British Journal of General Practice, 72(715):70–70, February 2022. ISSN 0960-1643, 1478-5242. doi: 10.3399/bjgp22X718361. URL https://bjgp.org/content/72/715/70. Publisher: British Journal of General Practice Section: Life & Times.

Paddy Ssentongo, Anna E. Ssentongo, Navya Voleti, Destin Groff, Ashley Sun, Djibril M. Ba, Jonathan Nunez, Leslie J. Parent, Vernon M. Chinchilli, and Catharine I. Paules. SARS-CoV-2 vaccine effectiveness against infection, symptomatic and severe COVID-19: a systematic review and meta-analysis. BMC Infectious Diseases, 22(1):439, May 2022. ISSN 1471-2334. doi: 10.1186/s12879-022-07418-y. URL https://doi.org/10.1186/s12879-022-07418-y.

Mohammad Talaei, Sian Faustini, Hayley Holt, David A. Jolliffe, Giulia Vivaldi, Matthew Greenig, Natalia Perdek, Sheena Maltby, Carola M. Bigogno, Jane Symons, Gwyneth A. Davies, Ronan A. Lyons, Christopher J. Griffiths, Frank Kee, Aziz Sheikh, Alex G. Richter, Seif O. Shaheen, and Adrian R. Martineau. Determinants of pre-vaccination antibody responses to SARS-CoV-2: a population-based longitudinal study (COVIDENCE UK). BMC Medicine, 20(1):87, February 2022. ISSN 1741-7015. doi: 10.1186/s12916-022-02286-4. URL https://doi.org/10.1186/s12916-022-02286-4.

Miller TE, Beltran WFG, Bard, Gogakos T, Anahtar MN, Astudillo MG, Yang D, Thierauf J, Fisch AS, Mahowald GK, Fitzpatrick MJ, Nardi V, Feldman J, Hauser BM, Caradonna TM, Marble HD, Ritterhouse LL, Turbett SE, Batten J, Georgantas NZ, Alter G, Schmidt AG, Harris JB, Gelfand JA, Poznansky MC, Bernstein BE, Louis DN, Dighe A, Charles RC, Ryan ET, Branda JA, Pierce VM, Murali MR, Iafrate AJ, Rosenberg ES, and Lennerz JK. Clinical sensitivity and interpretation of PCR and serological COVID-19 diagnositcs for patients presenting to the hospital. The FASEB Journal, 34(10):13877–13884, 2020. doi: 10.1096/fj.202001700RR.

The Moss Group, Inc. SB 132: The Transgender Respect, Agency, and Dignity Act Implementation Review Report. Technical report, California Department of Corrections and Rehabilitation, November 2022. URL https://www.cdcr.ca.gov/prea/wp-content/uploads/sites/186/2023/03/Final-SB132-CDCR-Assessment-Report_ADA.pdf.

R. N. Thompson, J. E. Stockwin, R. D. van Gaalen, J. A. Polonsky, Z. N. Kamvar, P. A. Demarsh, E. Dahlqwist, S. Li, E. Miguel, T. Jombart, J. Lessler, S. Cauchemez, and A. Cori. Improved inference of time-varying reproduction numbers during infectious disease outbreaks. Epidemics, 29:100356, December 2019. ISSN 1878-0067. doi: 10.1016/j.epidem.2019.100356.

The New York Times. Early Coronavirus Cases in Nursing Homes, Prisons and Other Locations. The New York Times, January 2022. ISSN 0362-4331. URL https://www.nytimes.com/interactive/2021/us/covid-case-outbreaks.html.

Sherry Towers, Danielle Wallace, Jason Walker, John M. Eason, Jake R. Nelson, and Tony H. Grubesic. A study of SARS-COV-2 outbreaks in US federal prisons: the linkage between staff, incarcerated populations, and community transmission. BMC Public Health, 22(1):482, March 2022. ISSN 1471-2458. doi: 10.1186/s12889-022-12813-w. URL https://doi.org/10.1186/s12889-022-12813-w.

Viet-Thi Tran, Raphäel Porcher, Isabelle Pane, and Philippe Ravaud. Course of post COVID-19 disease symptoms over time in the ComPaRe long COVID prospective e-cohort. Nature Communications, 13(1):1812, April 2022. ISSN 2041-1723. doi: 10.1038/s41467-022-29513-z. URL https://www.nature.com/articles/s41467-022-29513-z. Number: 1 Publisher: Nature Publishing Group.

Yi-Ju Tseng, Karen L. Olson, Danielle Bloch, and Kenneth D. Mandl. Smart Thermometer–Based Participatory Surveillance to Discern the Role of Children in Household Viral Transmission During the COVID-19 Pandemic. JAMA Network Open, 6(6):e2316190, June 2023. ISSN 2574-3805. doi: 10.1001/jamanetworkopen.2023.16190. URL https://doi.org/10.1001/jamanetworkopen.2023.16190.

Simone Turner, M. Asad Khan, David Putrino, Ashley Woodcock, Douglas B. Kell, and Etheresia Pretorius. Long COVID: pathophysiological factors and abnormalities of coagulation. Trends in Endocrinology & Metabolism, April 2023. ISSN 1043-2760. doi: 10.1016/j.tem.2023.03.002. URL https://www.sciencedirect.com/science/article/pii/S1043276023000553.

UCLA Law COVID Behind Bars Data Project. UCLA Law COVID Behind Bars Data, n.d. URL https://github.com/uclalawcovid19behindbars/data.

U.S. Census Bureau. B03002 Hispanic Or Latino Origin by Race, 2021 ACS 1-Year Estimates Detailed Tables, n.d. Retrieved June 2, 2024 from https://data.census.gov/table/ACSDT1Y2021.B03002\?q=B03002\&g=040XX00US06.

U.S. Census Bureau, Population Division. Annual Estimates of the Resident Population by Sex, Race, and Hispanic Origin for California: April 1, 2020 to July 1, 2021. https://www2.census.gov/programs-surveys/popest/tables/2020-2021/state/asrh/sc-est2021-sr11h-06.xlsx. Accessed 2023-05-25, June 2022. URL https://www2.census.gov/programs-surveys/popest/tables/2020-2021/state/asrh/sc-est2021-sr11h-06.xlsx.

Danielle Wallace, John M. Eason, Jason Walker, Sherry Towers, Tony H. Grubesic, and Jake R. Nelson. Is There a Temporal Relationship between COVID-19 Infections among Prison Staff, Incarcerated Persons and the Larger Community in the United States? International Journal of Environmental Research and Public Health, 18(13):6873, January 2021. ISSN 1660-4601. doi: 10.3390/ijerph18136873. URL https://www.mdpi.com/1660-4601/18/13/6873. Number: 13 Publisher: Multidisciplinary Digital Publishing Institute.

Jacco Wallinga and Peter Teunis. Different Epidemic Curves for Severe Acute Respiratory Syndrome Reveal Similar Impacts of Control Measures. American Journal of Epidemiology, 160(6):509–516, September 2004. ISSN 0002-9262. doi: 10.1093/aje/kwh255. URL https://academic.oup.com/aje/article/160/6/509/79472.

Leah Wang. The U.S. criminal justice system disproportionately hurts Native people: the data, visualized, 2021. URL https://www.prisonpolicy.org/blog/2021/10/08/indigenouspeoplesday/.

Alysse G Wurcel, Emily Dauria, Nicholas Zaller, Ank Nijhawan, Curt Beckwith, Kathryn Nowotny, and Lauren Brinkley-Rubinstein. Spotlight on Jails: COVID-19 Mitigation Policies Needed Now. Clinical Infectious Diseases, 71(15):891–892, July 2020. ISSN 1058-4838. doi: 10.1093/cid/ciaa346. URL https://doi.org/10.1093/cid/ciaa346.

