## Supplemental Material for "COVID-19 Reproduction Numbers and Long COVID Prevalences in California State Prisons"

### Supplement A. Mathematical methods used in estimation of daily incidence and reproduction numbers

We estimate effective reproduction numbers and timing of transmission events by a technique based on Wallinga-Teunis estimation (Wallinga and Teunis, 2004; Cauchemez et al., 2006; Fraser, 2007; Cori et al., 2013; Thompson et al., 2019; Gostic et al., 2020). The Wallinga-Teunis technique is a way of estimating a single time-varying reproduction number for an epidemic from a single series of daily case counts. For that, the key piece of information that is needed about the disease transmission process is the serial interval distribution, or more precisely *detection interval*, describing how likely a case is to be caused by a case detected a given number of days before. From that, likelihoods are derived for each possible transmission link, and reproduction numbers are estimated by the expected number of cases caused by a given case. The method described by Wallinga and Teunis (Wallinga and Teunis, 2004; Cauchemez et al., 2006; Fraser, 2007) estimates case reproduction numbers, typically denoted $R$ or $R_c$, the number of cases caused by a given source case over the source case's full disease progression, while a variant technique (Cori et al., 2013; Fraser, 2007) estimates effective reproductive numbers ($R$, or $R_t$), describing transmission rates on a single day, such that the number would be the case reproduction number if conditions were unchanging in time.

We describe here a technique for estimating multiple daily effective reproduction numbers describing transmission in multiple subpopulations, as well as case reproduction numbers and several other quantities, when data is available describing an outbreak in a structured population. Specifically, we describe the use of a data set including multiple positive and negative test results, symptom onset dates, and daily movements in a population structured across multiple locations, to estimate the dates of transmission events and effective reproduction numbers by location by day. This approach may be generalizable to other structured transmission processes.

Briefly, to place our exposition in context, the key steps of the Wallinga-Teunis technique are as follows. Assume a set of cases $x$ with case reporting dates $T_x$, and detection interval distribution $\sigma$, such that $\sigma(\Delta t)$ is the probability that a randomly selected source case and secondary case have reporting dates separated by $\Delta t$ days. Let $T(x)$ be the random variable describing the reporting date of any case $x$. Write the likelihood of a transmission link between any two distinct cases:

$$\begin{aligned} w(x \to y) &= L(\, x \to y \,|\, T(y) - T(x) = T_y - T_x \,) \\ &= \mathbb{P}(\, T(y) - T(x) = T_y - T_x \,|\, x \to y \,) \\ &= \sigma(T_y - T_x). \end{aligned}$$

Assume $N$ total cases. For a given secondary case $y$, the probability that a given $x$ is the source case for $y$ is derived by a Bayesian step, with a prior assumption that all $x$ other than $y$ have equal probability $1/(N-1)$ of being the source case, and that detection intervals are uniformly distributed with probability $c$ per day.

Then

$$
\begin{aligned}
p(x \to y) &= \mathbb{P}(\, x \to y \,|\, T(y) - T(z) = T_y - T_z \text{ for all } z \neq y \,) \\
&= \frac{\mathbb{P}(\, T(y) - T(z) = T_y - T_z \text{ for all } z \neq y \,|\, x \to y \,)\; \mathbb{P}(\, x \to y \,)}{\mathbb{P}(\, T(y) - T(z) = T_y - T_z \text{ for all } z \neq y \,)} \\
&= \frac{\mathbb{P}(\, T(y) - T(z) = T_y - T_z \text{ for all } z \neq y \,|\, x \to y \,)\; \mathbb{P}(\, x \to y \,)}{\sum_{x' \neq y} \mathbb{P}(\, T(y) - T(z) = T_y - T_z \text{ for all } z \neq y \,|\, x' \to y \,)\; \mathbb{P}(\, x' \to y \,)} \\
&= \frac{c^{N-2}\, w(x \to y)/(N-1)}{\sum_{x' \neq y} c^{N-2}\, w(x' \to y)/(N-1)} \\
&= \frac{w(x \to y)}{\sum_{x' \neq y} w(x' \to y)}.
\end{aligned}
$$

Given probabilities $p$ for all potential transmission links, the case reproduction number $R_{\mathrm{c}}(x)$ for each case $x$ is then estimated by the expected number of transmission links from $x$:

$$R_{\mathrm{c}}(x) = \sum_{y} p(x \to y).$$

Our method uses the same step of Bayesian inference to infer probabilities for all possible transmission links, while extending the above method in several ways. We proceed in two main steps. First, dates of positive and negative test results and symptom onsets are used to construct a probability distribution of days on which each individual was likely infected, and on which they were likely infectious. Second, those distributions are used together with daily movements of individuals to construct likelihoods $w(x \to y, t)$ for occurrence of each potential transmission link on each day, and transmission probabilities $p(x \to y, t)$ are inferred by a Bayesian update from the ensemble of $w(x \to y, t)$. From that we derive daily effective reproduction numbers $R$ per individual, day, and location, as well as case reproduction numbers and daily incidence per location.

*Supplement A.1. Estimation of individual incidence and infectiousness profile*

We estimate the timing of individuals' incidence and infectiousness by using a likelihood model parametrized by incidence date $t_0$ and incubation period $t_i$. For each individual $x$ we are given a collection $T^p_+(x)$ and $T^p_-(x)$ of dates of positive and negative RT-PCR tests, $T^a_+(x)$ and $T^a_-(x)$ of dates of positive and negative antigen tests, and $T_s(x)$ of (zero or one) symptom onset dates, which we notate as a vector $T(x) = (T^p_+(x), T^p_-(x), T^a_+(x), T^a_-(x), T_s(x))$.

Following CDCR's reporting policy (California Department of Corrections and Rehabilitation, n.d.a), We consider a positive test result more than 90 days after an initial positive test result to indicate a new infection or reinfection. When an individual has been infected more than once, they are recorded as multiple cases, and the following estimation is conducted once for each case, using all positive test results for that case and all negative test results for the individual. Symptom onset dates are associated with the latest case detected no later than the onset date.

Sensitivity of RT-PCR and antigen testing as a function of time since infection and incubation period have been fit from studies on test sensitivity after symptom onset (PS et al., 2020; TE et al., 2020). We use these as a probability $p^p_+(t_+|t_0, t_i)$ for positive and ($p^p_-(t_-|t_0, t_i) = 1 - p_+(t_-|t_0, t_i)$) negative RT-PCR test results given infection date and symptom onset, and similarly $p^a_+(t_+|t_0, t_i)$ and $p^a_-(t_-|t_0, t_i)$ for antigen tests (Figure A.1A, B).

We model the distribution of reported symptom onset dates as randomly perturbed from the theoretical symptom onset date, given wide observed variance in reported dates relative to positive test results, using

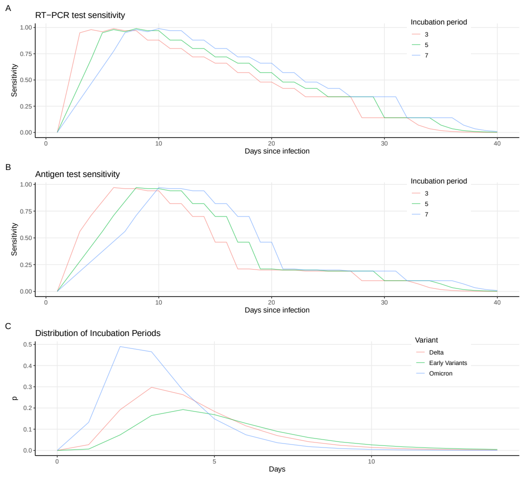


Figure A.1: **Estimated test sensitivities and timing distributions. A.** Probability of positive RT-PCR test given infection, as a function of time since infection, for three example choices of incubation period. **B.** Probability of positive antigen test given infection, as a function of time since infection, for three example choices of incubation period. **C.** Probability distribution of incubation period, in days.

a Cauchy distribution $p_s(t_s|t_0, t_i)$ with center at zero days from the end of the incubation period and scale parameter 6 days.

For a given infection date and incubation period, we can use the above sensitivity curves to provide a likelihood from the known testing and symptom data:

$$\begin{aligned} L_{\mathrm{t}}(t_0, t_i; x) &= L(\, t_0, t_i \,|\, T(x)\,) \\ &= \mathbb{P}(\, T_+^p(x), T_-^p(x), T_+^a(x), T_-^a(x), T_s(x) \,|\, t_0, t_i\,) \\ &= \prod_{t_+ \in T_+^p(x)} p_+^p(t_+|t_0, t_i) \prod_{t_- \in T_-^p(x)} p_-^p(t_-|t_0, t_i) \\ &\qquad \prod_{t_+ \in T_+^a(x)} p_+^a(t_+|t_0, t_i) \prod_{t_- \in T_-^a(x)} p_-^a(t_-|t_0, t_i) \prod_{t_s \in T_s(x)} p_s(t_s|t_0, t_i). \end{aligned}$$

We estimate the unknown parameters $t_0$ and $t_i$ by a Bayesian update from a naive prior. The prior distribution for $t_0$ is uniform with daily probability $\epsilon$. The prior distribution for the incubation period $t_i$ is taken from published estimates: for the original and Alpha variants of SARS-CoV-2, we assume a log-normal distribution $p_i(t_i)$ with mean 5.42 days and standard deviation 2.7 days (McAloon et al., 2020). For the Delta variant, the above distribution is rescaled to have mean 4/5 of the above, and for the Omicron variant it is rescaled to have mean 3/5 of the above (Jansen, 2021). The parametrization for the original variant is used until July 1, 2021, at which time the Delta variant is assumed to be dominant, until December 24, 2021, when it is replaced by Omicron (California) (Figure A.1C).

Then

$$\mathbb{P}(\, t_0, t_i \,|\, T(x)\,) = \frac{\mathbb{P}(\, T(x) \,|\, t_0, t_i\,)\, \epsilon\, p_i(t_i)}{\sum_{t_0'} \sum_{t_i'} \mathbb{P}(\, T(x) \,|\, t_0', t_i'\,)\, \epsilon\, p_i(t_i')}.$$

Ferretti et al. (Ferretti et al., 2020) provide the best known estimates of the time intervals between infection, symptom onset, and transmission events. The infectious period is correlated with the time of symptom onset in individuals who develop symptoms. Infectiousness increases gradually from infection to symptom onset, and the infectious period tends to increase with the incubation period. The time from onset of symptoms to a given transmission event ("TOST") is best fit by a skew-logistic distribution parametrized by the incubation period $t_i$:

$$p_{tost}(t|t_i) = \begin{cases} \frac{e^{-(t-\mu)/\sigma}}{(1+e^{-(t-\mu)/\sigma})^{\alpha+1}} & \text{for } t \geq 0 \\ \frac{e^{-(t\tau/t_i-\mu)/\sigma}}{(1+e^{-(t\tau/t_i-\mu)/\sigma})^{\alpha+1}} & \text{for } t < 0 \end{cases}$$

with $\mu = -4.00$ days, $\sigma = 1.85$ days, $\alpha = 5.85$, $\tau = 5.42$ days, normalized to produce a probability mass function. Note that the TOST can be positive, zero, or negative, depending on whether transmission occurs before, concurrent with, or after the onset of symptoms.

That yields a distribution of incidence dates and infectious periods, with incidence distribution

$$w_i(t_0; x) = \sum_{t_i} \mathbb{P}(\, t_0, t_i \,|\, T(x)\,)$$

and infectiousness distribution

$$w_\lambda(t; x) = \sum_{t_0} \sum_{t_i} p_{tost}(\, t - (t_0 + t_i) \,|\, t_i\,)\; \mathbb{P}(\, t_0, t_i \,|\, T(x)\,).$$

Although these quantities are derived from the posterior values of the above Bayesian estimation, we will sometimes refer to them as prior or raw incidence and infectiousness, because they are used as inputs to the second Bayesian step described below, which yields a posterior estimate of incidence among other quantities.

*Likelihood and probability of transmission links*

Whereas the derivation of transmission probabilities from likelihoods we reviewed above uses a detection interval distribution applied to the intervals between cases' reporting dates, we have multiple dates describing the disease progression of each individual, and so we must adapt the method. We note that the detection interval from case $x$ to $y$ is in fact the sum of multiple intervals: the interval from onset of $x$'s symptoms to transmission from $x$ to $y$, plus the interval from infection of $y$ to onset of symptoms of $y$, plus the interval from symptom onset of $y$ to detection of $y$, minus the interval from symptom onset of $x$ to detection of $x$. It can be decomposed more simply as the interval from the transmission date to detection of $y$, plus the interval from detection date of $x$ to the transmission event. Given distributions $w_i(s)$ and $w_\lambda(s)$ of incidence and daily infectiousness at $s$ days after (or $-s$ days before) the detection of a case, the detection interval distribution is expressed by a convolution of probability distributions:

$$\sigma(\Delta t) = \sum_s w_\lambda(s)\; w_i(s - \Delta t),$$

where $w_\lambda(s)\; w_i(s - \Delta t)$ is the probability that the transmission event occurred $s$ days after the detection of case $x$ and that case $y$ was detected $\Delta t$ days after case $x$, given transmission from $x$ to $y$.

We use this formulation without case detection dates, to construct likelihoods of transmission events from their infectiousness and incidence, respectively:

$$\begin{aligned} w(x \to y, t) &= L(\, x \to y \text{ on day } t \,|\, T(z) = T_z \text{ for all } z\,) \\ &= w_\lambda(x, t)\; w_i(y, t). \end{aligned}$$

The published Wallinga-Teunis technique uses a uniform prior probability of transmission for all pairs, updated using timing information to a posterior distribution. Here we have location information to distinguish some transmission events as more likely than others, and we include this as a prior assumption. We assume a uniform probability $\alpha_{\text{wb}}$ of transmission when a pair is located in the same building. When they are not the same, transmission is assumed to have a smaller probability $\alpha_{\text{bb}} < \alpha_{\text{wb}}$. When the pair is located in the same room, the transmission probability is a higher value $\alpha_{\text{wr}} > \alpha_{\text{wb}}$.

Let $B(x,t)$ stand for the building housing $x$ on day $t$, and $R(x,t)$ the room housing $x$ on day $t$, and let $F$ stand for the set of buildings that are within the institution (prison) being modeled. Then the prior probability of transmission is assumed to be

$$\mathbb{P}(x \to y, t) = \alpha(x,y,t)\ d$$

where

$$\alpha(x,y,t) = \begin{cases} \alpha_{\text{wr}} & \text{if } R(x,t) = R(y,t), B(x,t) \in F, B(y,t) \in F, \\ \alpha_{\text{wb}} & \text{if } R(x,t) \neq R(y,t), B(x,t) = B(y,t), B(x,t) \in F, B(y,t) \in F, \\ \alpha_{\text{bb}} & \text{if } B(x,t) \neq B(y,t), B(x,t) \in F, B(y,t) \in F, \\ 0 & \text{otherwise.} \end{cases}$$

We model one CDCR institution at a time, while individual prison residents are sometimes moved from one institution to another. We assume transmission between specific residents to have zero likelihood when either is not at the institution being modeled. Instead, to avoid pathological results, we assume a constant force of infection applied to individuals on days they are not at the institution. Similarly, we assume a constant small force of infection on residents within the institution, to allow for the possibility of transmission to residents from staff, visitors, or other sources.

We model this by assuming a constant daily likelihood that a case is an index case, that is, one whose transmission source is outside the population being modeled:

$$\begin{aligned} w_{\text{index}}(y,t) &= L(\, y \text{ an index case on day } t \,|\, T(y) = T_y\,) \\ &= \lambda_{\text{index}}\ w_i(y,t). \end{aligned}$$

This is coupled with a prior probability

$$\delta(y,t) = \begin{cases} \delta_{\text{inside}} & \text{if } B(y,t) \in F, \\ \delta_{\text{outside}} & \text{otherwise.} \end{cases}$$

We can then use the above likelihoods and priors to infer transmission probabilities per pair per day, by Bayesian inference with an uninformed prior assumption of uniform probability $c$ for values of the $T$ vector, and as discussed above for transmission events:

$$
\begin{aligned}
p(x \to y, t) &= \mathbb{P}(\, x \to y \text{ on day } t \,|\, T(z) = T_z \text{ for all } z\,) \\
&= \frac{\mathbb{P}(\, T(z) = T_z \text{ for all } z \,|\, x \to y \text{ on day } t\,)\; \mathbb{P}(\, x \to y \text{ on day } t\,)}{\mathbb{P}(\, T(z) = T_z \text{ for all } z\,)} \\
&= \mathbb{P}(\, T(z) = T_z \text{ for all } z \,|\, x \to y \text{ on day } t\,)\; \mathbb{P}(\, x \to y \text{ on day } t\,) \;/ \\
&\qquad \Big[\sum_{t'} \Big[ \sum_{x' \neq y} \mathbb{P}(\, T(z) = T_z \text{ for all } z \,|\, x' \to y \text{ on day } t'\,)\, \mathbb{P}(\, x' \to y \text{ on day } t'\,) + \\
&\qquad \mathbb{P}(\, T(z) = T_z \text{ for all } z \,|\, y \text{ an index case on day } t'\,)\, \mathbb{P}(\, y \text{ an index case on day } t'\,) \Big]\Big];
\end{aligned}
$$

or

$$
\begin{aligned}
p(x \to y, t) &= \frac{c^{N-2}\, w(x \to y, t)\, \alpha(x, y, t)\, d}{\sum_{t'} \Big[ \sum_{x' \neq y} c^{N-2}\, w(x' \to y, t')\, \alpha(x, y, t)\, d + c^{N-1}\, w_{\text{index}}(y, t')\, \delta(y, t) \Big]} \\
&= \frac{w(x \to y, t)\, \alpha(x, y, t)}{\sum_{t'} \Big[ \sum_{x' \neq y} w(x' \to y, t')\, \alpha(x, y, t) + (c/d)\, w_{\text{index}}(y, t')\, \delta(y, t) \Big]}.
\end{aligned}
$$

The constant factor $c/d$ can be absorbed, along with $\lambda_{\text{index}}$, into the values of the constants $\delta_{\text{inside}}$ and $\delta_{\text{outside}}$.

These transmission link probabilities do not in general sum to one, because there is some probability of being an index case:

$$
p_{\text{index}}(y, t) = \frac{(c/d)\, w_{\text{index}}(y, t)\, \delta(y, t)}{\sum_{t'} \Big[ \sum_{x' \neq y} w(x' \to y, t')\, \alpha(x, y, t) + (c/d)\, w_{\text{index}}(y, t')\, \delta(y, t) \Big]}.
$$

*Estimation of effective reproduction number*

Since this procedure estimates the timing as well as source of each case's infection, it can be used to estimate both case and effective reproduction numbers. An individual's case reproduction number is estimated as in the standard Wallinga-Teunis method. The total probability of transmission from $x$ to $y$ is $p(x \to y) = \sum_t p(x \to y, t)$, and the case reproduction number is

$$
R_{\text{c}}(x) = \sum_y p(x \to y).
$$

If needed, this estimate can be adjusted as in (Cauchemez et al., 2006) when some of case $x$'s infectious period was spent away from the institution, or had not yet been observed when the data was recorded.

An effective reproduction number $R$ is a description of conditions at a moment in time, equal to the number of cases that would be produced by a case over the case's entire history if those conditions were held constant. The relation between case and instantaneous reproduction numbers is the reconstruction of the case $R_{\text{c}}(x)$ from the instantaneous $R(x, t)$ over time:

$$
R_{\text{c}}(x) = \sum_t R(x, t)\, w_\lambda(x, t).
$$

These conditions can be satisfied by defining an individual's effective reproduction number on day $t$ as a ratio of incidence to infectiousness:

$$R(x,t) = \sum_y p(x \to y, t)/w_\lambda(x,t).$$

Here we present a mathematical derivation of this estimate, and a means of estimating its variance and confidence intervals.

For notation, assume a population $\mathbf{X}$ of individuals. Every case $y$ has an unknown day of infection $t_0(y)$. Every non-index case has an unknown source case $s(y)$ and we can put $s(y) = \emptyset$ for an index case. A transmission network is a choice of two values per case for these unknowns:

$$\mathbf{T} = (\,(s_y, t_y) \text{ for } y \in \mathbf{X}\,).$$

The Wallinga-Teunis estimate of transmission links assumes that the source case and infection date of one case are independent of others', so that the probability structure is the product of their probabilities:

$$\begin{aligned}\mathbb{P}(\,\mathbf{T} = (\,(s_y, t_y) \text{ for } y \in \mathbf{X}\,) \,|\, \text{data}\,) &= \prod_{y \in \mathbf{X}} \mathbb{P}(\,(s(y), t_0(y)) = (s_y, t_y) \,|\, \text{data}\,) \\ &= \prod_{y \in \mathbf{X}} p(s_y \to y, t_y).\end{aligned}$$

Assume each individual $x$ has an unobserved daily effective reproduction number $R(x,t)$, and generates a Poisson number of secondary cases with mean $R(x,t)\, w_\lambda(x,t)$ on day $t$.

We want a probability structure for the variables $R(x,t)$ based on data, which we will derive using the Wallinga-Teunis estimate for transmission networks:

$$\begin{aligned}&\mathbb{P}(\,R(x,t) = R_{x,t} \text{ for all } x,t \,|\, \text{data}\,) \\ &\quad = \sum_{\text{all networks } \mathbf{T}} \mathbb{P}(\,R(x,t) = R_{x,t} \text{ for all } x,t \,|\, \mathbf{T} = (\,(s_y,t_y)\,)\,)\; \mathbb{P}(\,\mathbf{T} = (\,(s_y,t_y)\,) \,|\, \text{data}\,)\end{aligned}$$

The second factor of that expression is the Wallinga-Teunis estimate. The first factor needs to be defined using Bayes's theorem. For readability let $dR_{x,t}$ be shorthand for the infinitesimal interval $[R_{x,t}, R_{x,t}+dR_{x,t})$:

$$\begin{aligned}&\mathbb{P}(\,R(x,t) \in dR_{x,t} \text{ for all } x,t \,|\, \mathbf{T} = (\,(s_y,t_y)\,)\,) \\ &\quad = \frac{\mathbb{P}(\,\mathbf{T} = (\,(s_y,t_y)\,) \,|\, R(x,t) \in dR_{x,t} \text{ for all } x,t\,)\; \mathbb{P}(\,R(x,t) \in dR_{x,t} \text{ for all } x,t\,)}{\mathbb{P}(\,\mathbf{T} = (\,(s_y,t_y)\,)\,)} \\ &\quad = \frac{\mathbb{P}(\,\mathbf{T} = (\,(s_y,t_y)\,) \,|\, R(x,t) \in dR_{x,t} \text{ for all } x,t\,)\; \mathbb{P}(\,R(x,t) \in dR_{x,t} \text{ for all } x,t\,)}{\int \mathbb{P}(\,\mathbf{T} = (\,(s_y,t_y)\,) \,|\, R(x,t) \in dR_{x,t} \text{ for all } x,t\,)\; \mathbb{P}(\,R(x,t) \in dR_{x,t} \text{ for all } x,t\,)}\end{aligned}$$

The likelihood of a network given a collection of $R$ values depends on each $R$ only through the number of secondary cases attributed to it: let $r_{x,t}(\mathbf{T}) = \#\{y | (s_y, t_y) = (x,t)\}$ where $\mathbf{T} = (\,(s_y, t_y)\,)$, and

$$\begin{aligned}
&\mathbb{P}(\,\mathbf{T} = (\,(s_y,t_y)\,)\,|\,R(x,t) = R_{x,t} \text{ for all } x,t\,) \\
&\quad = \prod_{x,t} \mathbb{P}(\,\#\{(s(y),t_0(y)) = (x,t)\} = r_{x,t}(\mathbf{T})\,|\,R(x,t) = R_{x,t}\,) \\
&\quad = \prod_{x,t} \frac{(R_{x,t}\,w_\lambda(x,t))^{r_{x,t}(\mathbf{T})}\,e^{-R_{x,t}\,w_\lambda(x,t)}}{r_{x,t}(\mathbf{T})!}.
\end{aligned}$$

We take an uninformative prior for $R$ defined by $R(x,t)\,w_\lambda(x,t) \sim \text{Gamma}(\varepsilon,\varepsilon)$ with vanishingly small $\varepsilon$, independent for each $x,t$. Independence gives us

$$\begin{aligned}
&\mathbb{P}(\,R(x,t) \in dR_{x,t} \text{ for all } x,t\,|\,\mathbf{T} = (\,(s_y,t_y)\,)\,) \\
&\quad = \frac{\prod_{x,t} \mathbb{P}(\,\#\{(s(y),t_0(y)) = (x,t)\} = r_{x,t}(\mathbf{T})\,|\,R(x,t) = R_{x,t}\,)\;\mathbb{P}(\,R(x,t) \in dR_{x,t}\,)}{\int \prod_{x,t} \mathbb{P}(\,\#\{(s(y),t_0(y)) = (x,t)\} = r_{x,t}(\mathbf{T})\,|\,R(x,t) = R_{x,t}\,)\;\mathbb{P}(\,R(x,t) \in dR_{x,t}\,)} \\
&\quad = \frac{\prod_{x,t} \mathbb{P}(\,\#\{(s(y),t_0(y)) = (x,t)\} = r_{x,t}(\mathbf{T})\,|\,R(x,t) = R_{x,t}\,)\;\mathbb{P}(\,R(x,t) \in dR_{x,t}\,)}{\prod_{x,t} \int \mathbb{P}(\,\#\{(s(y),t_0(y)) = (x,t)\} = r_{x,t}(\mathbf{T})\,|\,R(x,t) = R_{x,t}\,)\;\mathbb{P}(\,R(x,t) \in dR_{x,t}\,)} \\
&\quad = \prod_{x,t} \mathbb{P}(\,R(x,t) \in dR_{x,t}\,|\,\mathbf{T} = (\,(s_y,t_y)\,)\,)
\end{aligned}$$

where each $R(x,t)$ can be inferred independently from $\mathbf{T}$ using only the relevant number of secondary cases.

To infer a posterior distribution from the number of secondary cases, we note that the gamma distribution is a conjugate prior for the Poisson distribution of events, and the posterior is known to be

$$R(x,t)\,w_\lambda(x,t) \sim \text{Gamma}(\,r_{x,t}(\mathbf{T}), 1\,)$$

(we take $w_\lambda(x,t)$ as a known constant for these steps), which makes the mean of $R(x,t)$ equal to $r_{x,t}(\mathbf{T})/w_\lambda(x,t)$ and its variance $r_{x,t}(\mathbf{T})/w_\lambda(x,t)^2$.

Returning to the problem of estimation from data, let $G(\,R\,|\,r_{x,t}(\mathbf{T})\,)$ be the density function of the above gamma distribution, then

$$\begin{aligned}
&\mathbb{P}(\,R(x,t) \in dR_{x,t}\,|\,\text{data}\,) \\
&\quad = \sum_{\text{all networks } \mathbf{T}} \mathbb{P}(\,R(x,t) \in dR_{x,t}\,|\,\mathbf{T}\,)\;\mathbb{P}(\,\mathbf{T} = (\,(s_y,t_y)\,)\,|\,\text{data}\,) \\
&\quad = \sum_{\text{all networks } \mathbf{T}} G(\,R_{x,t}\,|\,r_{x,t}(\mathbf{T})\,)\,dR_{x,t}\;\mathbb{P}(\,\mathbf{T} = (\,(s_y,t_y)\,)\,|\,\text{data}\,) \\
&\quad = \sum_r G(\,R_{x,t}\,|\,r\,)\,dR_{x,t}\;\mathbb{P}(\,r_{x,t}(\mathbf{T}) = r\,|\,\text{data}\,).
\end{aligned}$$

The random variable $r_{x,t}(\mathbf{T})$ is a sum of Bernoulli random variables,

$$r_{x,t}(\mathbf{T}) = \sum_y 1(x \to y, t),$$

whose probabilities $p(x \to y, t)$ are known. The expected value of $R(x,t)$ is then

$$\begin{aligned}
\mathbb{E}(\, R(x,t) \,|\, \text{data}\,) &= \sum_r \mathbb{E}(\, R_{x,t} \,|\, r\,)\; \mathbb{P}(\, r_{x,t}(\mathbf{T}) = r \,|\, \text{data}\,) \\
&= \sum_r (r/w_\lambda(x,t))\; \mathbb{P}(\, r_{x,t}(\mathbf{T}) = r \,|\, \text{data}\,) \\
&= \mathbb{E}(\, r_{x,t}(\mathbf{T}) \,|\, \text{data}\,)/w_\lambda(x,t) \\
&= \frac{\sum_y p(x \to y, t)}{w_\lambda(x,t)}.
\end{aligned}$$

Assuming the effective number of terms of the sum is reasonably large, a normal approximation can be used to estimate confidence intervals for $R(x,t)$ numerically, using its mean and variance:

$$\begin{aligned}
\text{Var}(\, R(x,t) \,|\, \text{data}\,) &= \sum_r \text{Var}(\, R_{x,t} \,|\, r\,)\; \mathbb{P}(\, r_{x,t}(\mathbf{T}) = r \,|\, \text{data}\,) \\
&= \sum_r (r/w_\lambda(x,t)^2)\; \mathbb{P}(\, r_{x,t}(\mathbf{T}) = r \,|\, \text{data}\,) \\
&= \mathbb{E}(\, r_{x,t}(\mathbf{T}) \,|\, \text{data}\,)/w_\lambda(x,t)^2 \\
&= \frac{\sum_y p(x \to y, t)}{w_\lambda(x,t)^2}.
\end{aligned}$$

Because of the role of the infectiousness profile $w_\lambda()$ in the relation between $R$ and case reproduction numbers, we consider $w_\lambda()$ to be a natural weighting for the daily reproduction numbers, and use it as a weighting for the $R$ values when constructing statistics and data visualization. When considering $R$ values at the level of buildings, we use the sum of infectiousness over the building as a weighting term.

*Effective reproduction number at aggregate level*

Is the effective reproduction number in a building best estimated by the average of individuals' $R$? What is its distribution? Here we estimate it directly from the transmission network distribution. The analogous steps can be used to estimate it at the room or institution level.

Using the notation from the previous section, we posit a single unobserved parameter $R(A,t)$ associated with location $A$ on day $t$, such that transmissions from individuals in location $A$ are a Poisson process with rate $R(A,t)\, w_\lambda(A,t)$, with $w_\lambda(A,t) = \sum_{\{\, x \,|\, B(x,t)=A\,\}} w_\lambda(x,t)$, and estimate the value of $R(A,t)$ directly.

Given a transmission network $\mathbf{T}$, the number of transmission events from $A$ at $t$ is

$$\begin{aligned}
r_{A,t}(\mathbf{T}) &= \sum_{\{x|B(x,t)=A\}} \sum_y 1(x \to y, t) \\
&= \sum_{\{x|B(x,t)=A\}} r_{x,t}(\mathbf{T}).
\end{aligned}$$

An uninformative gamma prior for $R(A,t)$ gives a posterior estimate of

$$R(A,t)\, w_\lambda(A,t) \sim \text{Gamma}(r_{A,t}(\mathbf{T}), 1),$$

with mean $\mathbb{E}(R(A,t)|\mathbf{T}) = r_{A,t}(\mathbf{T})/w_\lambda(A,t)$ and variance $\text{Var}(R(A,t)|\mathbf{T}) = r_{A,t}(\mathbf{T})/w_\lambda(A,t)^2$.

Across realizations of the transmission network, we obtain

$$\begin{aligned}
&\mathbb{P}(\, R(A,t) \in dR_{A,t} \,|\, \text{data}\,) \\
&\qquad = \sum_{\mathbf{T}} \mathbb{P}(\, R(A,t) \in dR_{A,t} \,|\, \mathbf{T}\,)\, P(\, \mathbf{T} = (\,(s_y,t_y)\,) \,|\, \text{data}\,) \\
&\qquad = \sum_{\mathbf{T}} G(\, R_{A,t} \,|\, r_{A,t}(\mathbf{T})\,)\, dR_{A,t}\, P(\, \mathbf{T} = (\,(s_y,t_y)\,) \,|\, \text{data}\,) \\
&\qquad = G(\, R_{A,t} \,|\, r\,)\, dR_{A,t}\, P(\, r_{A,t}(\mathbf{T}) = r \,|\, \text{data}\,).
\end{aligned}$$

As with $R(x,t)$, the estimate of $R(A,t)$ from data is a mixture of gamma variables with mean

$$\begin{aligned}
\mathbb{E}(\, R(A,t) \,|\, \text{data}\,) &= \sum_{\{x|B(x,t)=A\}} \sum_y p(x \to y,t)/w_\lambda(A,t) \\
&= \sum_{\{x|B(x,t)=A\}} \frac{w_\lambda(x,t)}{w_\lambda(A,t)} \mathbb{E}(R(x,t)|\text{data})
\end{aligned}$$

and variance

$$\begin{aligned}
\text{Var}(\, R(A,t) \,|\, \text{data}\,) &= \sum_{\{x|B(x,t)=A\}} \sum_y p(x \to y,t)/w_\lambda(A,t)^2 \\
&= \frac{\sum_{\{x|B(x,t)=A\}} w_\lambda(x,t)^2\, \text{Var}(R(x,t)|\text{data})}{w_\lambda(A,t)^2}.
\end{aligned}$$

This means the mean estimate is a weighted average of the individual means by infectiousness, not a straight average, and the variance is smaller than the individual estimates' variances, because $w_\lambda(x,t) < 1$.

*Estimation of incidence*

The probability that an individual was infected on a given day is

$$I(y,t) = \sum_x p(x \to y,t) + p_{\text{index}}(y,t).$$

This can be termed the expected incidence of case $y$ on day $t$. Note that this is the posterior incidence mentioned earlier, in distinction to the prior incidence $w_i(y,t)$ that is estimated from test and symptom reports without taking into account likelihood of transmission events. The true incidence in a location, in terms of the number of people infected in a location on a day (in contrast to the number of cases detected per location per day), is estimated by

$$I(A,t) = \sum_{\{x|B(x,t)=A\}} I(x,t).$$

### Supplement B. Statistical testing for infection risk

We used Poisson regression to examine associations between race/ethnicity, gender, and rate of infection, controlling for age and building location. The following R code was used:

```
glmer(inc ~ age.class + Gender + Race + Gender:Race + offset(log(years)) + (1|loc), data=.,
family=poisson(link='log'))
```

This model can be expressed as

$$\begin{aligned}\log \mathbb{E}[Y \mid a., g., r.] &= \beta_0 + \beta_1 a_2 + \beta_2 a_3 + \beta_3 a_4 + \beta_4 g_{\mathrm{CW}} + \beta_5 g_{\mathrm{TGNBI}} + \beta_6 r_{\mathrm{A}} + \cdots + \beta_{10} r_{\mathrm{W}} \\ &\quad + \beta_{11} g_{\mathrm{CW}} r_{\mathrm{A}} + \cdots + \beta_{20} g_{\mathrm{TGNBI}} r_{\mathrm{W}} + e \\ &= \beta_0 + \sum_k \beta_k X_k + e\end{aligned}$$

where $a.$, $g.$ and $r.$ are indicator functions for the levels of age, gender, and race, omitting levels 1, CM, and B respectively.

Then $\beta_0$, the intercept, is a prediction of the log rate of events per year for a Black cisgender man in age class 1. The difference in prediction from age class 1 to age class 2 is $\beta_1$, assumed to be independent of gender and race. For a white cisgender man in age class 1, the prediction is

$$\log \mathbb{E}[Y] = \beta_0 + \beta_{10},$$

while if the same person were a cisgender woman it would be

$$\log \mathbb{E}[Y] = \beta_0 + \beta_{10} + \beta_4 + \beta_{19},$$

since both $g_{\mathrm{CW}}$ and $g_{\mathrm{CW}} r_{\mathrm{W}}$ are affected. The effect of being CW rather than CM depends on race: for white people it is $\beta_4 + \beta_{19}$, while for black people it is $\beta_4$.

Our hypotheses to test are

- The prediction for gender CW is equal to the whole-population rate, and the same for TGNBI and CM.
- The rate for race/ethnicity B is equal to the whole-population rate, and the same for each other level of the race/ethnicity variable.
- The rate for e.g., CW B, and all other gender-race tuples, is no different from the prediction given the gender and race/ethnicity, but without the interaction term.

These hypotheses will be expressed by a contrast matrix, each row of which is a contrast vector expressing one of these hypotheses as a combination of variables to be compared to zero in a two-sided test.

We begin by writing the log-rate for the whole population. This is $\log \langle Y \rangle$, the log-rate taken over the entire data set. Given that $Y = \beta_0 + \sum_k \beta_k X_k + e$, that should be equal to

$$\log \mathbb{E}[Y] = \beta_0 + \sum_k \beta_k \langle X_k \rangle = \beta_0 + \frac{\sum_k \beta_k \sum_i (X_k)_i}{\sum_i 1}$$

over all the observations $i$ in the population.[2]

The effect of being CW rather than CM, for a given observation $i$, is $\beta_4 + \sum_k \beta_{9+2k} (r_k)_i$. What is the effect of being CW compared to the population as a whole? The log-rate over all CW observations is

$$\log \mathbb{E}[Y \mid g_{CW} = 1] = \beta_0 + \frac{\sum_i (g_{CW})_i \sum_k \beta_k (X_k)_i}{\sum_i (g_{CW})_i},$$

[2] An observation in this regression is not a person, but a collection of person-days with a common age class, gender, race/ethnicity, and location. Here when I sum over the index $i$ it should be understood as shorthand for summing over each person-day in the set of observations.

and the difference between that and the whole-population log-rate is

$$\begin{aligned}\log \mathbb{E}[Y \mid g_{CW}=1] - \log \mathbb{E}[Y] &= \frac{\sum_k \beta_k \sum_i (g_{CW})_i (X_k)_i}{\sum_i (g_{CW})_i} - \frac{\sum_k \beta_k \sum_i (X_k)_i}{\sum_i 1} \\ &= \sum_k \beta_k \sum_i \left( \frac{(g_{CW})_i}{n_{CW}} - \frac{1}{n} \right) (X_k)_i \\ &= \sum_k \beta_k c_k\end{aligned}$$

where $c$ is a contrast vector, if

$$\begin{aligned} c_k &= \sum_i \left( \frac{(g_{CW})_i}{n_{CW}} - \frac{1}{n} \right) (X_k)_i = \frac{1}{n_{CW}} \langle g_{CW}, X_k \rangle - \langle X_k \rangle \\ c = c_{CW} &= \left( \frac{1}{n_{CW}} g_{CW} - \frac{1}{n} \mathbf{1} \right)^{\mathsf{T}} X, \end{aligned}$$

$X$ being the design matrix $\big((X_k)_i\big)$.

The contrast for $g_{TGNBI}$ is defined the same way. For $g_{CM}$ it is the same as well, except that it is implicit in the design matrix rather than explicit, because $CM$ is the reference level that was omitted:

$$\begin{aligned} c_{CM} &= \left( \frac{1}{n_{CM}} g_{CM} - \frac{1}{n} \mathbf{1} \right)^{\mathsf{T}} X \\ &= \left( \frac{1}{n - n_{CW} - n_{TGNBI}} (1 - g_{CW} - g_{TGNBI}) - \frac{1}{n} \mathbf{1} \right)^{\mathsf{T}} X \\ &= \left( \frac{n\,(1 - g_{CW} - g_{TGNBI}) - (n - n_{CW} - n_{TGNBI})\,\mathbf{1}}{n(n - n_{CW} - n_{TGNBI})} \right)^{\mathsf{T}} X \\ &= \left( \frac{n_{CW} - n\,g_{CW} + n_{TGNBI} - n\,g_{TGNBI}}{n(n - n_{CW} - n_{TGNBI})} \right)^{\mathsf{T}} X \\ &= \frac{1}{n - n_{CW} - n_{TGNBI}} \left( \frac{n_{CW}}{n} - g_{CW} + \frac{n_{TGNBI}}{n} - g_{TGNBI} \right)^{\mathsf{T}} X \\ &= \frac{1}{n - n_{CW} - n_{TGNBI}} \left( n_{CW} \left( \frac{1}{n} - \frac{1}{n_{CW}} g_{CW} \right) + n_{TGNBI} \left( \frac{1}{n} - \frac{1}{n_{TGNBI}} g_{TGNBI} \right) \right)^{\mathsf{T}} X \\ &= \frac{1}{n - n_{CW} - n_{TGNBI}} \big( - n_{CW} c_{CW} - n_{TGNBI} c_{TGNBI} \big) \\ &= -\frac{n_{CW}}{n_{CM}} c_{CW} - \frac{n_{TGNBI}}{n_{CM}} c_{TGNBI}. \end{aligned}$$

The race/ethnicity variable works just the same way, including a substitution for the reference level.

Finally, to compare intersecting race/ethnicity-gender strata with the distribution implied by a lack of interaction, we must define that distribution. We name the effects defined above $\Delta Y_{CW}$, etc., so that e.g., $\log \mathbb{E}[Y \mid g_{CW}=1] = \log \mathbb{E}[Y] + \Delta Y_{CW}$. We that assume in the absence of interaction effects are additive, so that the expected value predicted by this additive model is e.g.

$$\log \mathbb{E}_N[Y \mid g_{CW}=1, r_W=1] = \log \mathbb{E}[Y] + \Delta Y_{CW} + \Delta Y_W.$$

We then compare that prediction to the true expected value with interactions:

$$\begin{aligned}\Delta Y_{CW,W} &= \log \mathbb{E}[Y \mid g_{CW} = 1, r_W = 1] - \log \mathbb{E}_N[Y \mid g_{CW} = 1, r_W = 1] \\ &= \frac{\sum_k \beta_k \sum_i (g_{CW})_i (r_W)_i (X_k)_i}{\sum_i (g_{CW})_i (r_W)_i} - (\log \mathbb{E}[Y] + \Delta Y_{CW} + \Delta Y_W) \\ c_{CW,W} &= \left( \frac{g_{CW} \odot r_W}{n_{CW,W}} - \frac{1}{n} \right)^{\mathsf{T}} X - c_{CW} - c_W\end{aligned}$$

where $\odot$ stands for the elementwise (Hadamard) product of vectors.

### Supplement C. Long COVID model details

We modeled the prevalence of long COVID in this population as a time-dependent probabilistic process at the individual level, influenced by age and vaccination. As the probability of experiencing long COVID symptoms declines with time since infection, with detailed dynamics unknown and long-term outcomes currently unknowable, we assume geometrically decaying probability of experiencing symptoms at $t$ days after onset of (acute) symptoms:

$$P(\text{ long COVID on day } t \text{ given infection }) = k_{\text{lc}}\, e_{\text{a}}(a)\, e_{\text{v}}(v)\, e_{\text{re}}(re)\, e_{\text{g}}(g)\, r^{t-21}, \quad t \geq 21$$

where $e_{\text{a}}(a)$ is the effect of age $(a)$, $e_{\text{v}}(v)$ is the effect of vaccination $(v)$, $e_{\text{re}}(re)$ is the effect associated with race and ethnicity $(re)$, $e_{\text{g}}(g)$ is the effect associated with gender $(g)$, and $k_{\text{lc}}$ and $r$ are constants.

We estimate these effects from available data. Age-specific prevalences of long COVID among COVID-19 patients are available from a U.S. study (FAIR Health, 2022):

$$e_{\text{a}}(a) = \begin{cases} 3.65, & a \leq 12 \\ 6.78, & 12 < a \leq 22 \\ 17.0, & 22 < a \leq 35 \\ 34.6, & 35 < a \leq 50 \\ 32.0, & 50 < a \leq 64 \\ 6.03, & 64 < a \end{cases}$$

Long COVID appears to occur less commonly in men than others, given infection, with relative risk of 33.0/21.0 for women compared to men, and 47.8/21.0 for TGNBI individuals (CDC, 2023):

$$e_{\text{g}}(g) = \begin{cases} 21.0, & g = \texttt{CM} \\ 33.0, & g = \texttt{CF} \\ 47.8, & g = \texttt{TGNBI}. \end{cases}$$

Race and ethnicity also affect long COVID risk given infection, at rates (relative to non-Hispanic whites) of 31.5/26.7 for Hispanic or Latino, 18.5/26.7 for Asian, and 28.2/26.7 for Black respondents, and 38.1/26.7

for those of other or multiple races (CDC, 2023):

$$e_{\rm re}(re) = \begin{cases} 26.7, & re = \texttt{White} \\ 31.5, & re = \texttt{Latinx} \\ 18.5, & re = \texttt{Asian} \\ 28.2, & re = \texttt{Black} \\ 38.1, & \text{otherwise.} \end{cases}$$

Ballering *et al.* found the whole-population rate is 12.7% of patients with COVID-19 (Ballering et al., 2022), in a Dutch population of COVID-positive study participants with median age 52.4 years and standard deviation 11.7 years. That rate is measured in an almost entirely white and pre-vaccination population. The reported gender distribution is 58% women, and none TGNBI. We use it to provide a baseline for the probability in unvaccinated individuals by fixing known values to the study population averages $\langle e_{\rm a}\rangle_{\rm B}$, $\langle e_{\rm v}\rangle_{\rm B}$, $\langle e_{\rm re}\rangle_{\rm B}$, and $\langle e_{\rm g}\rangle_{\rm B}$, by assuming a Gaussian age distribution in the Ballering study, truncated at 0 and 100 years, no vaccination effect ($\langle e_{\rm v}\rangle_{\rm B} = 1$), all white, and gender distribution as reported, and solving for $k_{\rm lc}$:

$$0.127 = k_{\rm lc}\,\langle e_{\rm a}\rangle_{\rm B}\,\langle e_{\rm re}\rangle_{\rm B}\,\langle e_{\rm g}\rangle_{\rm B}.$$

Vaccination reduces the overall probability of long COVID given infection to 9%–11% (Al-Aly et al., 2022; Ayoubkhani et al., 2022). We conservatively use the low estimate of 9%, taking $e_{\rm v}(v) = 9/12.7$ for individuals with one or more vaccination dose and 1 for those without. The daily continuation rate $r$ of the syndrome is estimated from a French cohort study (Tran et al., 2022) in which 85% of long COVID cases that last to two months continue to one year.

The onset date for each case is assumed distributed across the infectious period according to the infectiousness profile that is estimated by our detailed transmission model ($w_\lambda(t;x)$; Supplement A.1). The resident data set provides birth years without months and days, so we estimate ages at the date of symptom onset based on a birthday of July 1.

64% of long COVID sufferers report that their symptoms have interfered with their day-to-day activities, and 19% report their ability to do daily living activities "limited a lot" (Ayoubkhani et al., 2021; Spiers, 2022). In addition to the above estimate of long COVID prevalence, we estimated the numbers of prison residents disabled by long COVID as time passes by estimating conservatively that 19% of long COVID cases become disabling.

The less stratified model for long COVID and associated disability is as above, but with $e_{\rm re}$ and $e_{\rm g}$ fixed at the white and cisgender male rates for all individuals.

We used the above models to estimate the number of prison residents who had long COVID symptoms, and who were disabled by them, over time and by location in the CDCR system, using race/ethnicity, gender, age, and vaccination status from the CDCR data set, and tracking individuals across multiple infections and transfers between locations. We aggregated the estimates into total numbers affected, and numbers by race/ethnicity and gender.

For California cases we used California Department of Health and Human Services statistics on COVID-19 cases by race and ethnicity (California Health and Human Services, 2023) and vaccination by race and ethnicity (California Health and Human Services, 2023) with U.S. Census Department estimates of California population by race and ethnicity (U.S. Census Bureau, Population Division, 2022). We estimated the proportion of cases who were vaccinated by estimating the efficacy of vaccination (one or more doses) at 75% (Ssentongo et al., 2022, Fig. 2), so that where $p_{\rm v}$ is the proportion of the population vaccinated,

$$\frac{0.25\,p_{\rm v}}{0.25\,p_{\rm v} + (1 - p_{\rm v})}$$

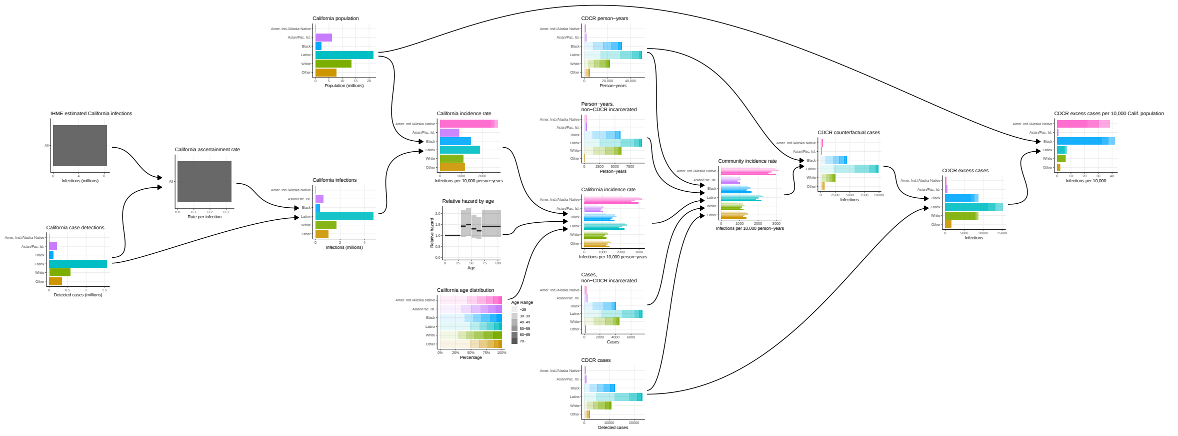


Figure D.1: **Flow diagram of steps in estimation of excess cases.** Gradations of transparency in horizontal bars indicate central estimate and 95% confidence intervals (confidence intervals not shown for estimates stratified by both age and race/ethnicity).

is the proportion of cases who were vaccinated.

### Supplement D. Estimation of excess cases

Figure D.1 depicts the steps of the estimation of excess cases in CDCR outbreaks from CDCR and California data.

We estimated the excess cases associated with CDCR outbreaks in the period March 1, 2020–April 1, 2021, bya series of steps (Figure D.1). Variance of estimates was propagated, except where otherwise noted, using an assumption of independent normally distributed variables with small coefficient of variation, under which

$$\begin{aligned}\mathbb{E}[AB] &= \mathbb{E}[A]\,\mathbb{E}[B]\\ \mathrm{Var}(AB) &= \mathrm{Var}(A)\ \mathrm{Var}(B) + \mathrm{Var}(A)\,\mathbb{E}[B]^2 + \mathbb{E}[A]^2\ \mathrm{Var}(B)\end{aligned}$$

and

$$\begin{aligned}\mathbb{E}[A/B] &= \frac{\mathbb{E}[A]}{\mathbb{E}[B]}\\ \mathrm{Var}(A/B) &= \frac{\mathbb{E}[A]^2}{\mathbb{E}[B]^2}\left(\frac{\mathrm{Var}(A)}{\mathbb{E}[A]^2} + \frac{\mathrm{Var}(B)}{\mathbb{E}[B]^2}\right)\end{aligned}$$

(Díaz-Francés and Rubio, 2013).

The Institute for Health Metrics and Evaluation has provided a model-based estimate of true infections by date in California, accounting for incomplete ascertainment in the reported case counts (Institute for Health Metrics and Evaluation, 2022; IHME COVID-19 Forecasting Team, 2021). We used IHME's model data to obtain an estimate of total infections during the first two waves of outbreaks identified.

Combining that with California's total reported cases during the same time period (California Health and Human Services Open Data Portal, 2023) produced an estimate of overall ascertainment rate. The first two moments of this estimate were estimated as follows. Given that $D$ of $I$ cases were detected, taking a vague

Gamma-distributed prior for the ascertainment rate $a$, and the number of detections Poisson distributed at rate $aI$, we obtain a posterior estimate of $aI$ Gamma distributed with mean $D$ and variance $D$, so that $a$ has mean $D/I$ and variance $D/I^2$. With $I$ being a random variable with mean $\bar{I}$ and variance $\bar{\bar{I}}$ (and likewise $\bar{D}$ and $\bar{\bar{D}}$ for $D$), using the above approximations we have

$$\begin{aligned}
\mathbb{E}[a] &= \mathbb{E}[a \mid D, I] \\
&= \mathbb{E}[D/I] \\
&= \frac{\bar{D}}{\bar{I}} \\
\operatorname{Var}(a) &= \mathbb{E}[\operatorname{Var}(a \mid D, I)] + \operatorname{Var}(\mathbb{E}[a \mid D, I]) \\
&= \mathbb{E}\left[\frac{D}{I^2}\right] + \operatorname{Var}\left(\frac{D}{I}\right) \\
&= \frac{\bar{D}}{\bar{I}^2} + \frac{\bar{D}^2}{\bar{I}^2}\left(\frac{\bar{\bar{D}}}{\bar{D}^2} + \frac{\bar{\bar{I}}}{\bar{I}^2}\right) \\
&= \frac{\bar{D}}{\bar{I}^2} + \frac{\bar{D}^2\,\bar{\bar{I}}}{\bar{I}^4},
\end{aligned}$$

since $D$ is known exactly.

Given that ascertainment rate and the number of confirmed cases by race/ethnicity, we derived an estimated true incidence by race/ethnicity. Combining true incidence with California population by race/ethnicity (U.S. Census Bureau, n.d.), we obtained an estimate of incidence rate in cases per person-year by race/ethnicity.

The purpose of estimating incidence rates per person-year in California was to estimate the counterfactual cases that would have occurred if CDCR residents had been infected at the rates occurring in the community. That counterfactual estimate required adjustment for the different age distributions of the resident and state populations. We obtained an estimate of the effect of age on infection from the UK-based COVIDENCE study (Talaei et al., 2022). Adjusted odds ratios for seroconversion were provided for each age class, up to 29, 30–39, 40–49, 50–59, 60–69, and 70 and over. These were used to estimate hazard ratios for infection by assuming rare events.

Using these relative hazards, California's age distribution by race/ethnicity, and the estimated incidence rate stratified by race/ethnicity, we estimated statewide incidence rates by age and race/ethnicity, as follows. Let $P_{a,r}$ be the population by age range and race/ethnicity, $h_r$ be the incidence rate by race/ethnicity, and let $HR_a$ be the relative hazard by age. Then we assume that the incidence rate $h_{a,r}$ by age and race/ethnicity satisfies two conditions:

$$h_r = \frac{\sum_a P_{a,r}\, h_{a,r}}{\sum_a P_{a,r}} \qquad \text{for each } h_r,$$

and

$$\frac{h_{a,r}}{h_{a_0,r}} = HR_a \qquad \text{for each } h_{a,r}.$$

This is satisfied by

$$h_{a,r} = h_r\, \frac{HR_a \sum_b P_{b,r}}{\sum_b HR_b P_{b,r}}.$$

We then adjusted that statewide incidence rate by removing CDCR and other incarcerated cases to obtain an estimate of incidence in the state outside the CDCR system. Specifically, from the incidence rate by age and race/ethnicity a corresponding estimate of absolute incidence was derived using the state population by age and race/ethnicity; stratified absolute incidence and total person-years were adjusted downward by the CDCR numbers; and stratified community incidence was estimated by adjusted infections per adjusted person-year.

Non-CDCR incarcerated statistics were estimated as follows. For California county jails, the race/ethnicity distribution was imputed from 2021 percentages (Prison Policy Initiative, n.d.), and age distribution from first quarter 2022 in LA County Jail (Los Angeles County Sheriff's Department, 2022). The state's federal prison population race/ethnicity and age distribution was imputed from 2022 national figures for U.S. Bureau of Prisons (BOP) sentenced residents (Carson and Kluckow, 2022). The race/ethnicity and age distribution of ICE detainees was imputed from 2019 figures from California facilities (California Department of Justice, Office of Justice Programs, 2019). In light of uncertainty due to imputation and possible missingness of data, in each of these three classes of facilities, the fractions in each age and race/ethnicity stratum were given confidence intervals uniform in width and wide enough to include zero in the stratum with the largest central estimate. The statewide case ascertainment rate implied by the IHME estimate is approimately 1 in 3; total incidence in these non-CDCR facilities was modeled by scaling case detections by a pair of bracketing estimates of ascertainment rate of 100% or 1 in 9 with equal probability. Given unstratified incidences, age- and race/ethnicity-specific incidence rates in the above populations were imputed from the population distributions, with equivalently wide confidence intervals. We note that the non-CDCR incarcerated population, at under 20,000 cases, accounts for less than 1% of the case detections in California during the two waves studied here.

Combining community incidence rates in infections per person-year by age and race/ethnicity with the count of person-years spent by all residents in CDCR institutions during the first two waves of outbreaks stratified by age and race/ethnicity, we obtained a counterfactual estimate of infections among CDCR residents at California community rates during that time.

We estimated excess cases from CDCR outbreaks by race/ethnicity, by subtracting the estimated counterfactual cases at community rates from the recorded cases at CDCR rates. We then converted that result to excess cases per 10,000 California population by race/ethnicity.

#### Supplement E. Outbreaks

Figure E.1 plots the daily rate of cases detected at each CDCR institution, the seven-day average of case detections in (A) and the daily positive and negative tests in (B). Figure E.2 shows the number of cases in each outbreak at each institution, alongside the institutions' population size as of March 20, 2020.

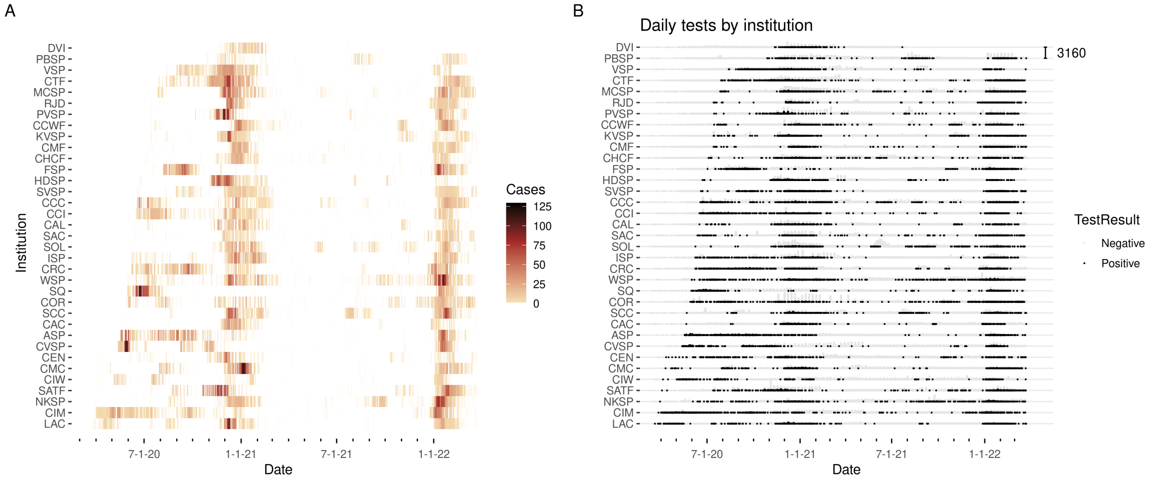


Figure E.1: **(A) Cases detected by testing, by day** (7-day average) by institution; **(B) Positive and negative tests per day** by institution. Institutions are ordered from bottom to top by the first date of detection of a case. Scale bar in upper right of (B) gives the highest number of tests administered in a single day in one institution.

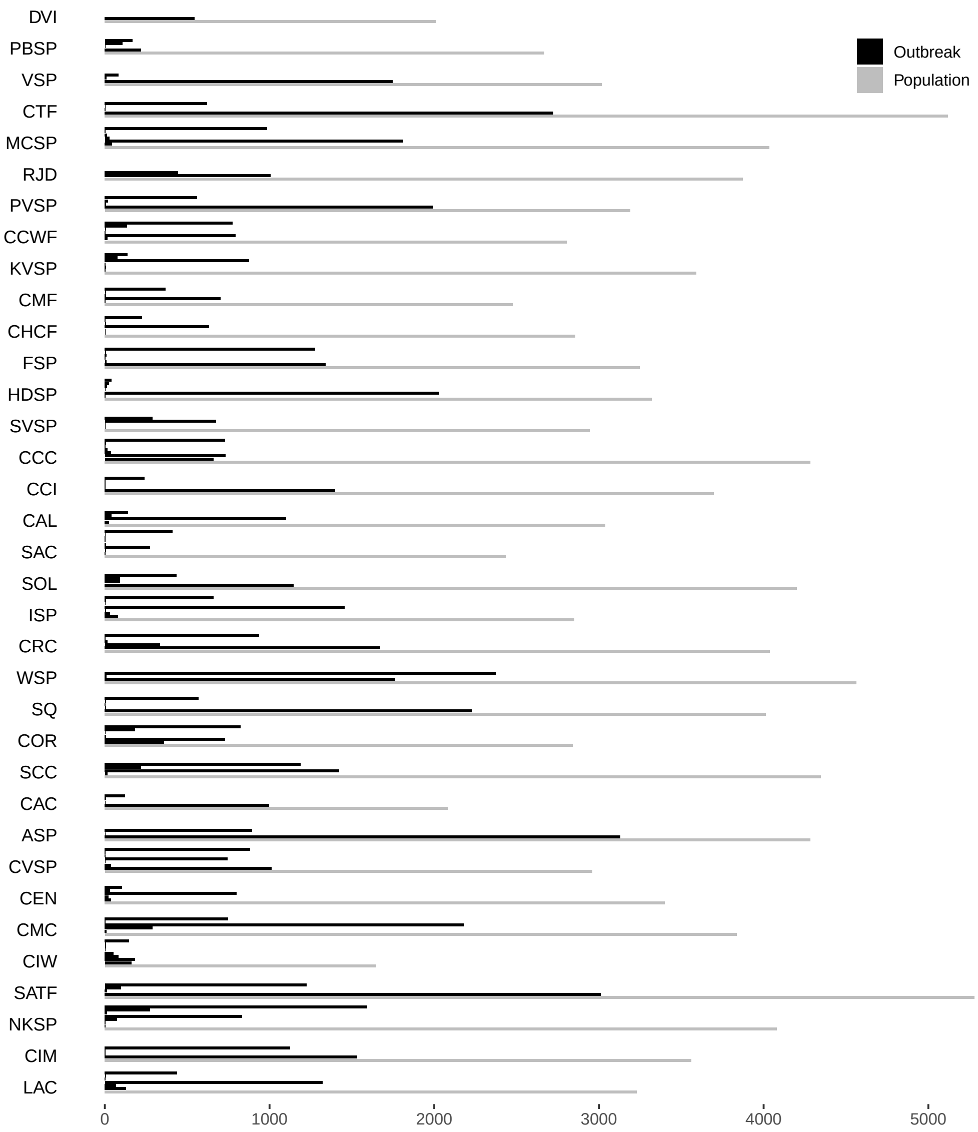


Figure E.2: Outbreak sizes by institution. Outbreaks of size 3 or greater are shown (black), with institution population size as of March 20, 2020 for comparison (gray). Adapted and updated from figure 1 of (Parsons and Worden, 2021).

### Supplement F. Comparisons by race/ethnicity and gender

Confirmed SARS-CoV-2 infections by race/ethnicity and gender are presented in Figure F.1, and corresponding incidence rates in cases per person-year in Figure F.2.

The adjusted estimates of effect of race/ethnicity, gender, and the intersection of the two are presented in Figure F.3. Estimated effects of non-intersected gender and race/ethnicity are relative to the population as a whole, and of intersections are relative to the effect predicted by gender and race/ethnicity separately. $P$-values are Bonferroni-Holm adjusted for multiple comparisons.

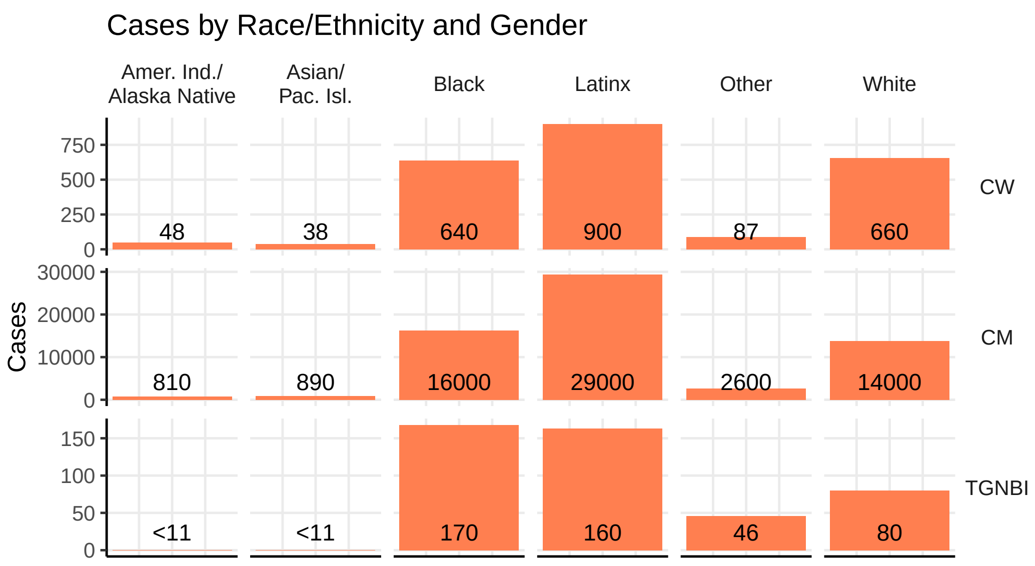


Figure F.1: **COVID-19 cases by race/ethnicity and gender** (C=cisgender, M=men, W=women, TGNBI=transgender, nonbinary, or intersex).

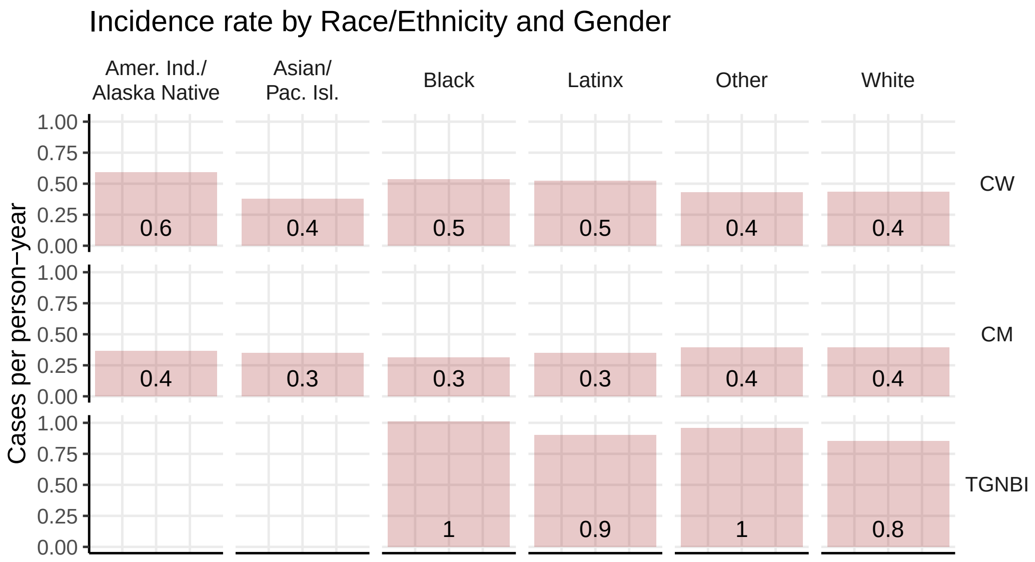


Figure F.2: **Estimated COVID-19 incidence rate by race/ethnicity and gender**, in infection events per person-year (C=cisgender, M=men, W=women, TGNBI=transgender, nonbinary, or intersex). Entries corresponding to small cell sizes in Figure F.1 are omitted, to avoid possible deductive disclosure.

Infection risk estimates

| | Estimate | Std. Error | *P* |
|---|---|---|---|
| CM | –0.0237 | 0.00543 | **<0.001** |
| CW | 0.597 | 0.145 | **<0.001** |
| TGNBI | 0.292 | 0.0412 | **<0.001** |
| Black | –0.0475 | 0.00634 | **<0.001** |
| Amer. Ind./Alaska Native | 0.0515 | 0.033 | 0.913 |
| Asian/Pac. Isl. | –0.0613 | 0.0316 | 0.661 |
| Latinx | 0.0019 | 0.00444 | 1.000 |
| Other | 0.0656 | 0.0172 | **0.003** |
| White | 0.0526 | 0.00787 | **<0.001** |
| CM Black | –0.000173 | 0.00106 | 1.000 |
| CM Amer. Ind./Alaska Native | –0.0225 | 0.00704 | **0.032** |
| CM Asian/Pac. Isl. | –0.0036 | 0.00793 | 1.000 |
| CM Latinx | 0.00474 | 0.00126 | **0.004** |
| CM Other | –0.00137 | 0.00346 | 1.000 |
| CM White | –0.00769 | 0.0031 | 0.248 |
| CW Black | 0.0326 | 0.0271 | 0.992 |
| CW Amer. Ind./Alaska Native | 0.176 | 0.111 | 0.905 |
| CW Asian/Pac. Isl. | –0.147 | 0.127 | 0.995 |
| CW Latinx | 0.0127 | 0.0217 | 1.000 |
| CW Other | –0.0218 | 0.0833 | 1.000 |
| CW White | –0.0623 | 0.0264 | 0.321 |
| TGNBI Black | 0.12 | 0.0448 | 0.151 |
| TGNBI Amer. Ind./Alaska Native | 0.0797 | 0.265 | 1.000 |
| TGNBI Asian/Pac. Isl. | –0.0761 | 0.245 | 1.000 |
| TGNBI Latinx | –0.0642 | 0.0494 | 0.983 |
| TGNBI Other | –0.0681 | 0.102 | 1.000 |
| TGNBI White | –0.0819 | 0.0793 | 0.999 |

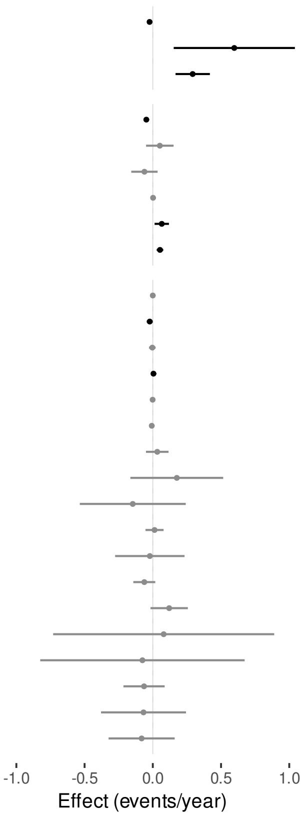


Figure F.3: Poisson regression estimates of effects of gender (C=cisgender, M=men, W=women, TGNBI=transgender, nonbinary, or intersex), race/ethnicity, and intersections of gender and race/ethnicity on COVID-19 infection risk. Estimated effects of non-intersected gender and race/ethnicity are relative to the population as a whole, and of intersections are relative to the effect predicted by gender and race/ethnicity separately. *P*-values are Bonferroni-Holm adjusted for multiple comparisons.

### Supplement G. Movements and Transfers

We compiled tables of number of transfers to and from each institution by day, and number of movements between rooms within each institution by day. We defined a transfer to include any transition of a resident from not being housed at an institution to being housed there the next night or vice versa; thus admissions from outside the prison system, releases, and deaths were included, as well as movements between institutions, and we distinguished transfers from, to, and between institutions in summary statistics. We defined a movement as any transition in nightly housing from a room to a different room at the same institution.

In the period from March 20, 2020, to March 25, 2022, there were 47,837 transfers of residents into the CDCR system, 72,777 out of it, and 76,549 between institutions (Figure G.1, G.2). Transfers were reduced substantially several times, including at the times of winter waves of outbreaks, and resumed afterward. We note that Central California Women's Facility (CCWF), North Kern State Prison (NKSP), and Wasco State Prison (WSP) were used by CDCR as reception centers during this time period, where new arrivals to the system were held initially before assignment to another location (Napoles, 2020), and that only CCWF, California Institution for Women (CIW), and Folsom State Prison (FSP) house women prisoners (FSP includes facilities for both men and women).

Residents were moved from one room to another within the same institution 803,482 times over the same time period. Movements were stopped and restarted in some institutions, but never paused system-wide. Movements were more frequent during the periods of COVID-19 outbreaks in the winter of 2020–21 and the winter of 2021–22 (Figure G.3).

To examine the effect of movement and transfer on the apparent location of disease transmission events, we plotted the estimated number of cases infected while housed in each room against the number of cases detected by positive tests while housed there (Figure G.4). Linear regression with intercept of zero by room type is shown by straight lines with ribbons indicating 95% confidence intervals.

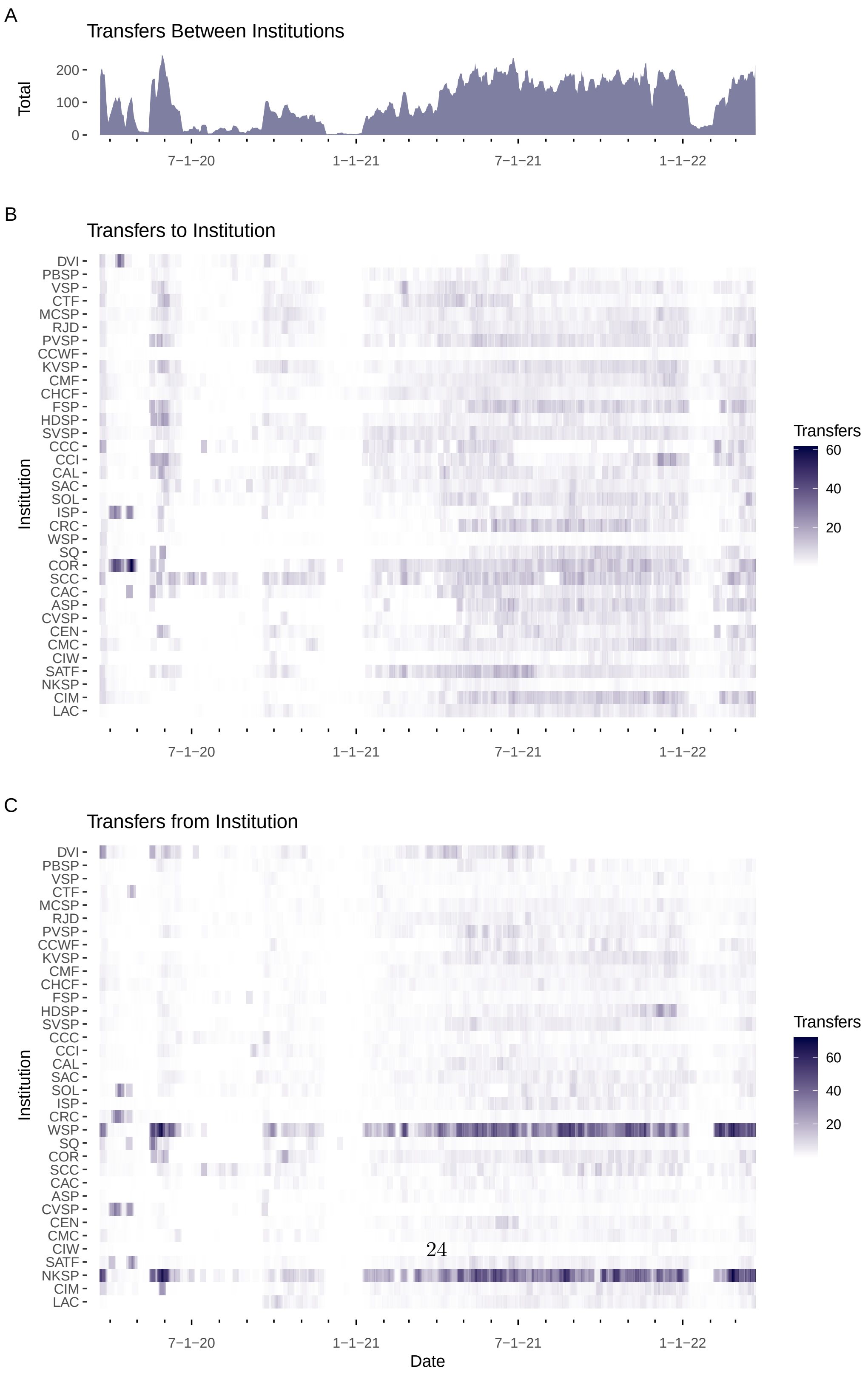




Figure G.1: **Daily transfers** (7-day average) of residents CDCR institutions: (A) over all institutions; (B) to each institution; (C) from each institution, including movements into and out of the CDCR system.

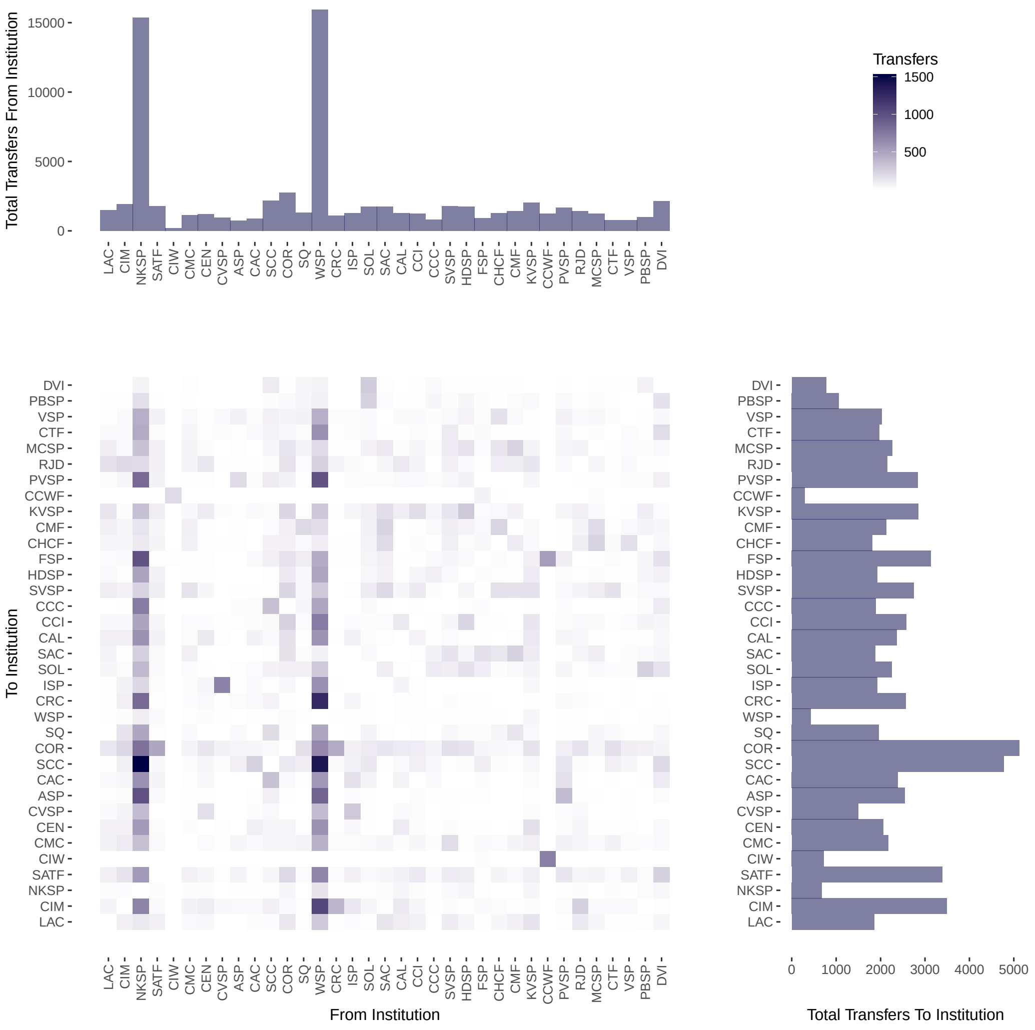


Figure G.2: **Transfers** of residents between CDCR institutions by source and destination institutions, with total number of transfers to and from each institution. Note women are housed at CCWF, CIW, and FSP only.

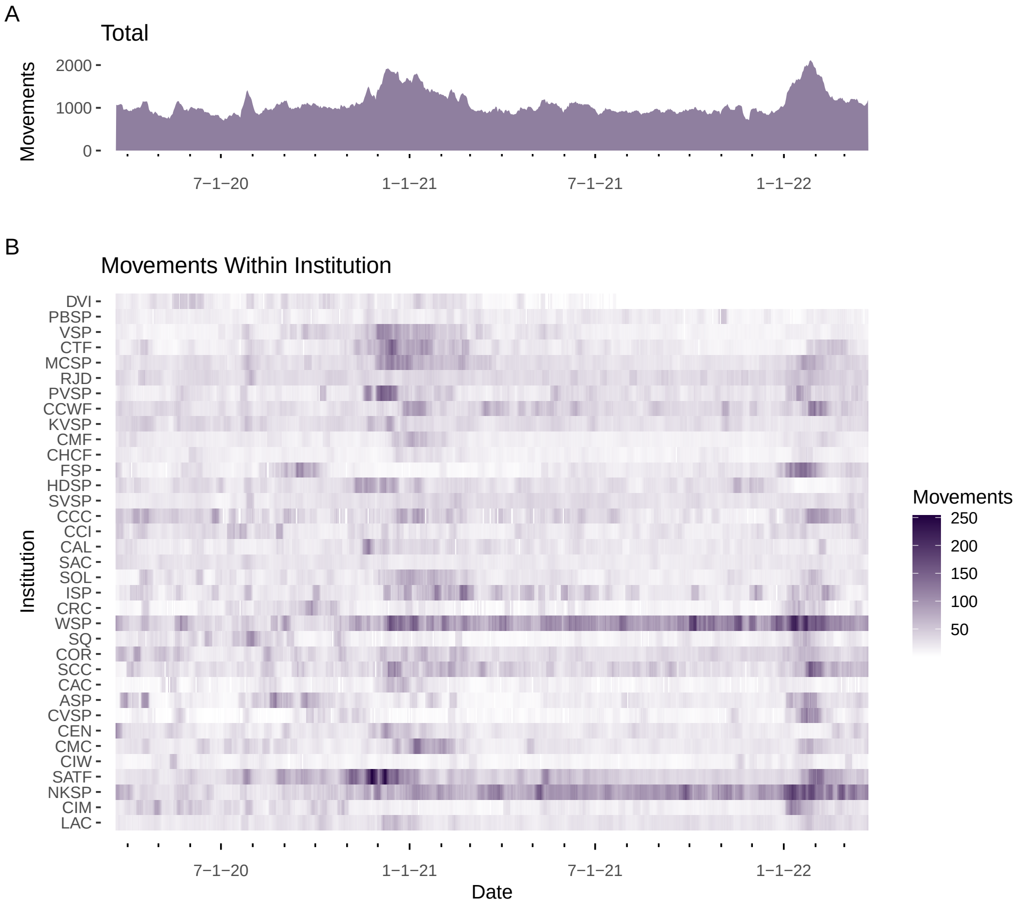


Figure G.3: **Daily movements** (7-day average) of residents between rooms within CDCR institutions: (A) over all institutions; (B) by institution.

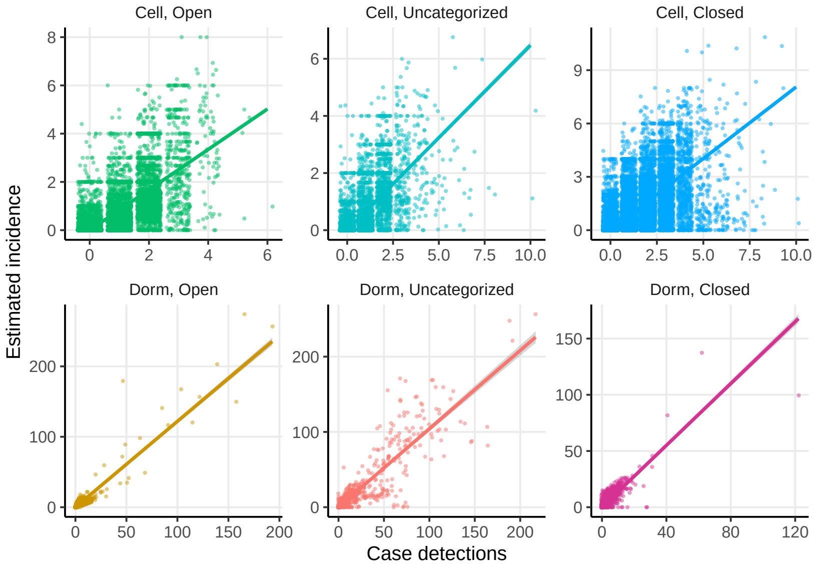


Figure G.4: Estimated number of cases infected (incidence) versus number of cases detected in each room, by room type.

### Supplement H. Long COVID estimates

The overall prevalences of long COVID and of disability due to long COVID over time, estimated by our long COVID model, are presented in Figures H.1 and H.2, respectively. Figure H.3 presents the estimated count of both (A) and rate of both per person-year (B) by race/ethnicity and gender.

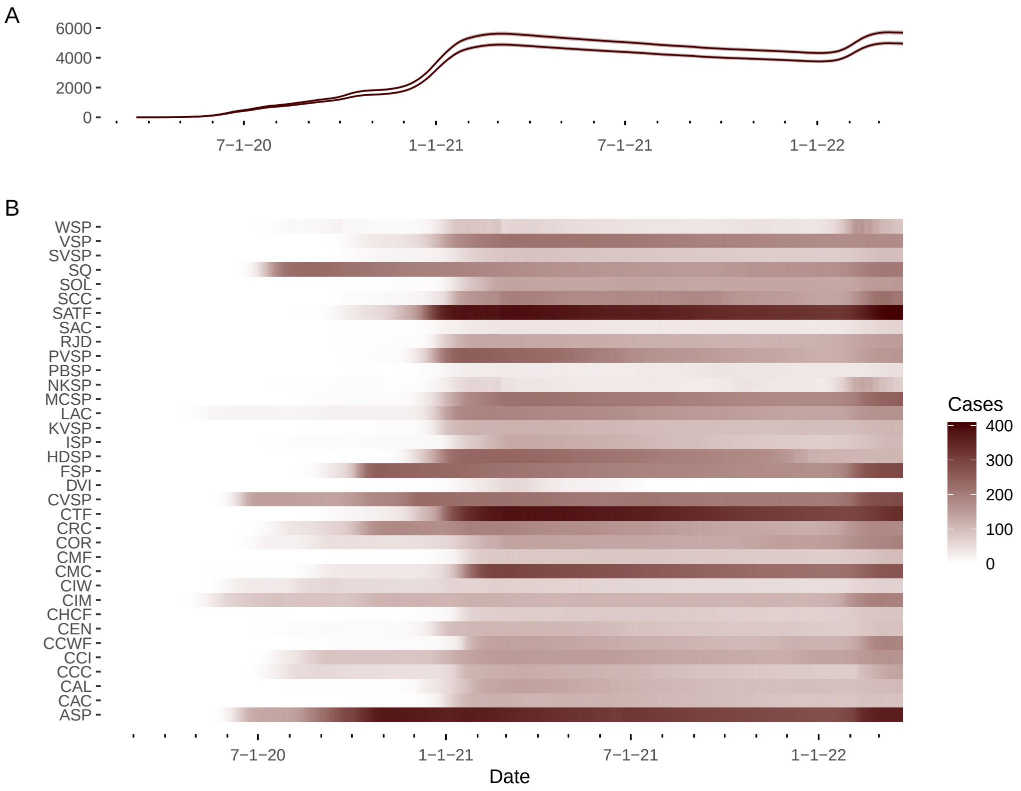


Figure H.1: **A. Estimated overall prevalence of long COVID across all CDCR residents**, from individuals' test results, ages, and vaccination records, accounting for movements, with 95% confidence intervals (upper curve: more stratified model, lower curve: less stratified), **B. Estimated distribution of long COVID prevalence across institutions**, taking into account movements. Prevalence may be underestimated because residents infected before entering the CDCR system are not estimated.

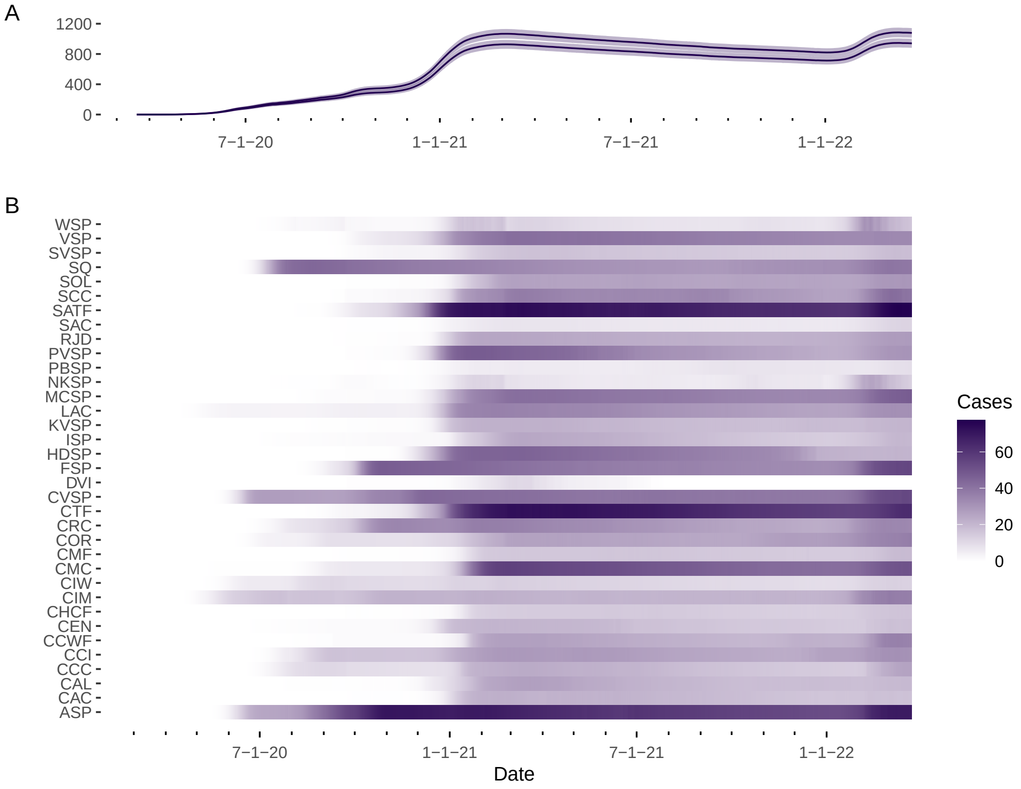


Figure H.2: **A. Estimated overall prevalence of disabling long COVID across all CDCR residents**, from individuals' test results, ages, and vaccination records, movements, with 95% confidence intervals (upper curve: more stratified model, lower curve: less stratified), **B. Estimated distribution of disabling long COVID prevalence across institutions**, taking into account movements and transfers.

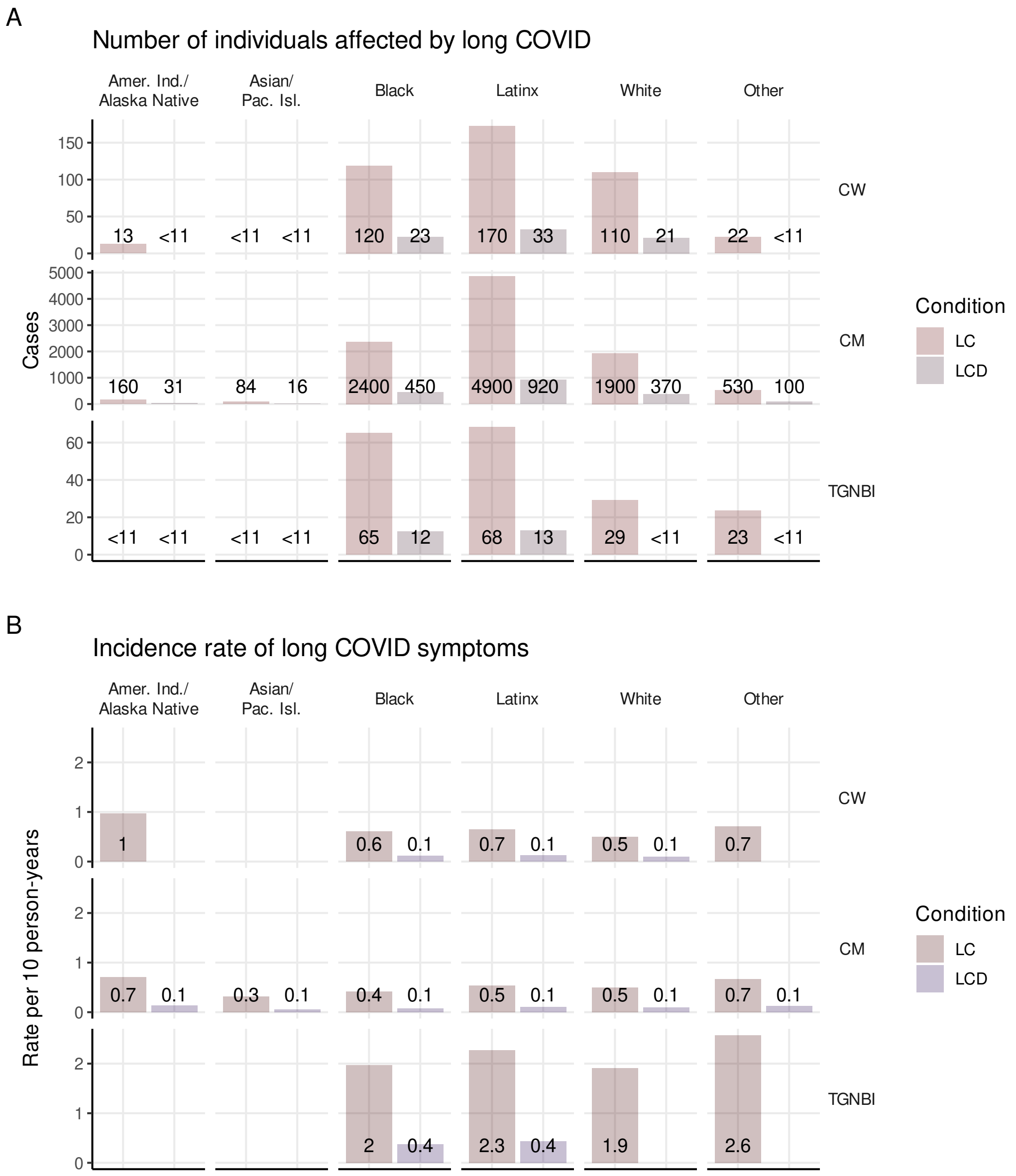


Figure H.3: **(A) Estimated count** and **(B) incidence rate** of long COVID (LC) and disability due to long COVID (LCD), by race/ethnicity and gender (C=cisgender, M=men, W=women, TGNBI=transgender, nonbinary, or intersex). Case counts of 10 or fewer are censored from figure (A), and corresponding values in (B) are omitted to avoid possible deductive disclosure.

### Supplement I. Reproduction number and incidence estimates by institution

Statistics and estimates of daily true incidence and effective reproduction number by building in the individual institutions are presented in Figures I.1–I.35, below.

Each figure includes six plots, summarizing the sequence of SARS-CoV-2 outbreaks at one CDCR institution. In plot (A) the overall number of cases detected each day was plotted. This provided a summary of the number, timing, size and duration of outbreaks, and of their rates of growth and decline during their progression. Vertical bars marked the start of each separate outbreak at the institution. Daily counts were shown as vertical bars, and the seven-day rolling average of daily counts was plotted as a darker curve.

In plot (B), the number of cases detected by positive tests each day was plotted for each building in the institution, together with the number of negative tests administered. All tests were plotted on their date of administration, not the date the results were received, and negative tests were shown stacked atop positive tests, so that total height indicated total tests administered. Buildings were arranged vertically, grouped by facility, with a black line next to the building ID number showing the grouping. Facilities were ordered from bottom to top by first positive test date, and buildings were ordered the same way within each facility. A scale bar was included in the upper right showing the number of tests in the tallest bar.

Plot (C) presented the reconstructed number of incident cases per day in each building. Transparency was used to display the number of cases, with higher numbers less transparent, and color was used to display the type of room occupied by individuals in each building. Days with expected incidence of 0.1 or more cases in a building were included. While some buildings contained multiple room types, overlap of colors in these plots was rare enough that we considered them useful. Buildings and facilities were ordered as in (B).

Plot (D) presented a histogram of reconstructed incident cases by room type in each outbreak. Numbers smaller than or equal to 10 cases were not labeled and were plotted as 10, for protection of individuals' privacy.

In plot (E), the estimated effective reproduction number ($R$) was plotted by day for each building. Because values of $R$ greater than one reflect ongoing spread of the disease, red color was used for values greater than one and blue for values less than one. Orange, yellow, and green were used for exceptionally high values. Days on which total infectiousness profile (see Supplement A) in a building was over 0.1 were shown. For comparison with figure 2, note that due to the reduced granularity of spatial divisions the threshold for plotting there is larger. Buildings and facilities were ordered as in (B).

In plot (F), the $R$ estimates included in plot (E) were summarized in a histogram for each room type. The vertical scale for each room type was rescaled, and heights do not afford a comparison of frequency between different room types in plot (F). Plot (D) provides that comparison. In some cases, room types appearing in plots (C) and/or (D) at very low counts did not meet the inclusion criterion for (E) and (F), and did not appear in (F).

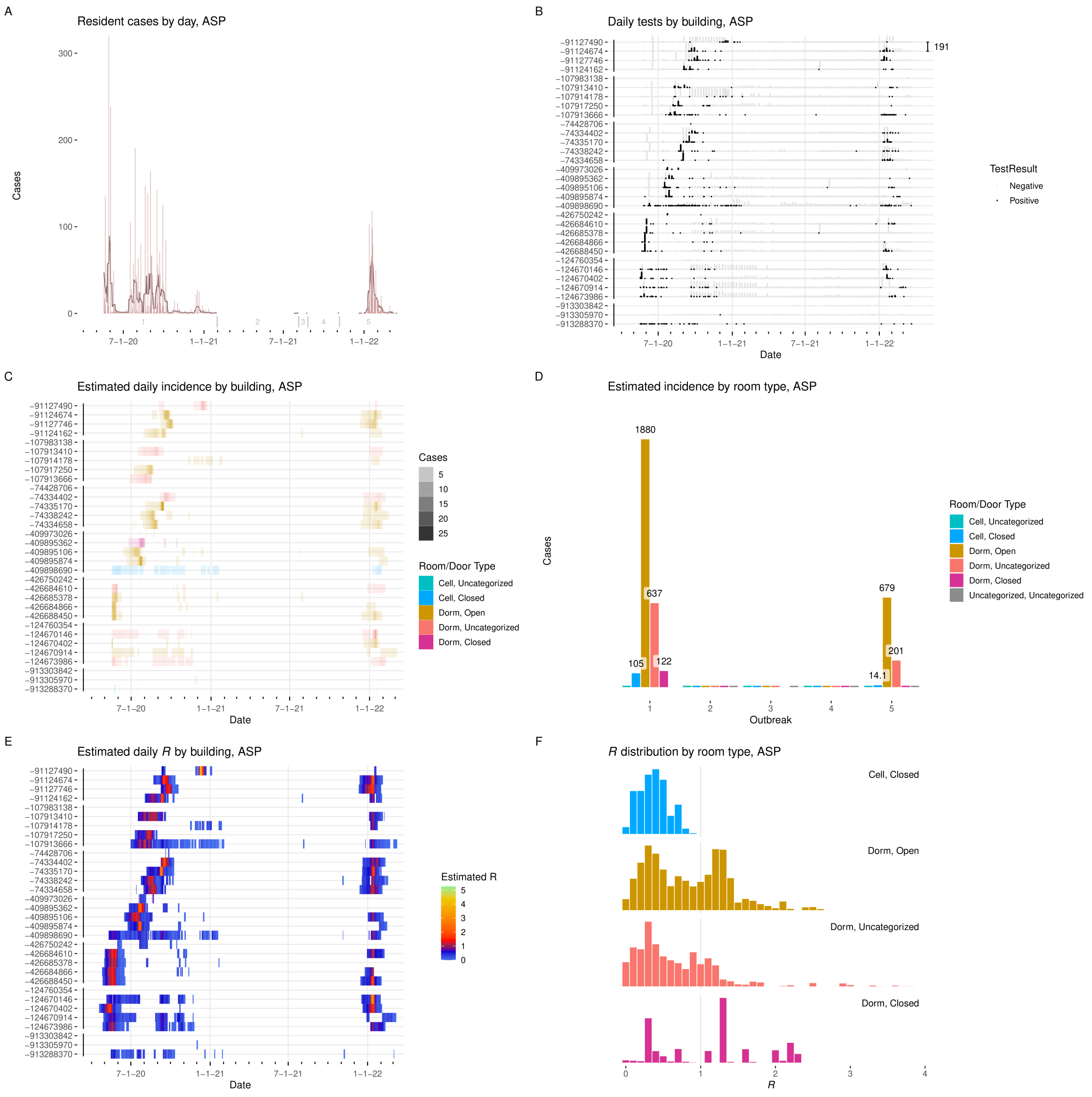


Figure I.1: **Results from ASP. A:** Cases detected per day, with seven-day average. Starting date of each outbreak is marked by a vertical gray line. **B:** Positive and negative tests per day by building. $Y$-axis labels are building ID numbers. Buildings are grouped by facility. Scale bar at upper right gives number of tests. **C:** Estimated incidence by day by building, colored by room type. **D:** Incidence by room type and outbreak. **E:** Estimated effective $R$ by day per building. **F:** Distribution of effective $R$ in each room type.

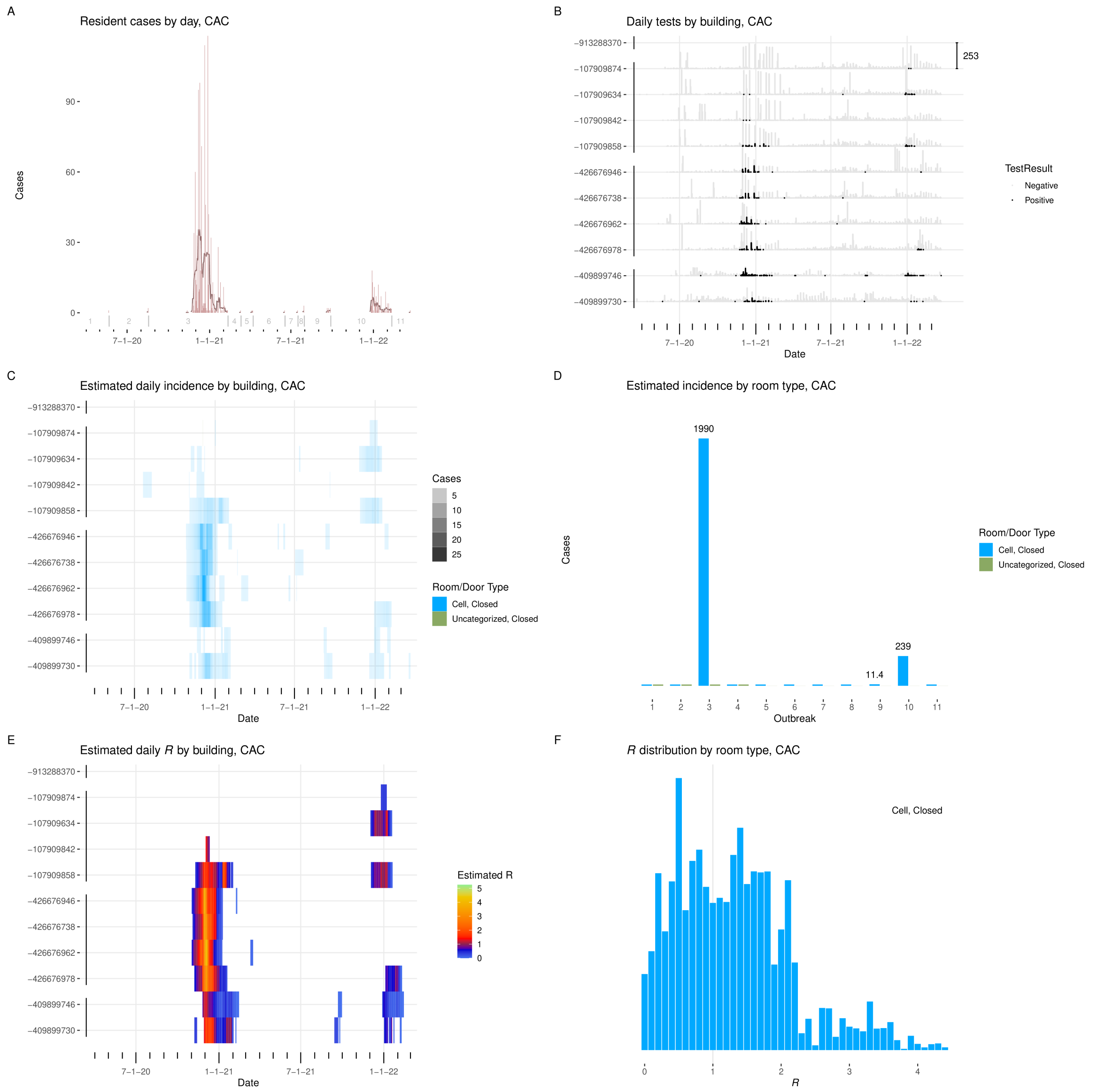


Figure I.2: **Results from CAC. A:** Cases detected per day, with seven-day average. Starting date of each outbreak is marked by a vertical gray line. **B:** Positive and negative tests per day by building. $Y$-axis labels are building ID numbers. Buildings are grouped by facility. Scale bar at upper right gives number of tests. **C:** Estimated incidence by day by building, colored by room type. **D:** Incidence by room type and outbreak. **E:** Estimated effective $R$ by day per building. **F:** Distribution of effective $R$ in each room type.

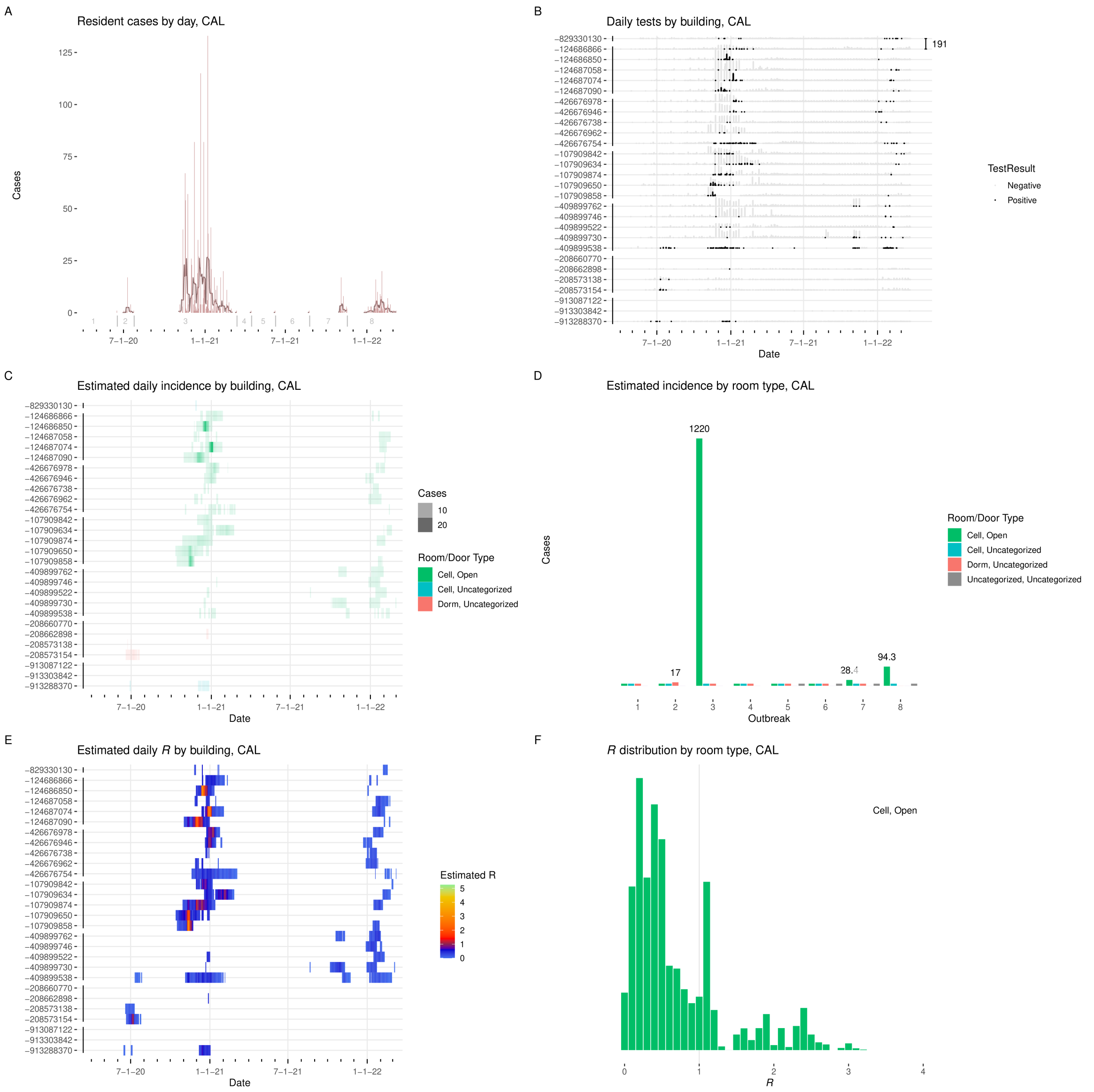


Figure I.3: **Results from CAL. A:** Cases detected per day, with seven-day average. Starting date of each outbreak is marked by a vertical gray line. **B:** Positive and negative tests per day by building. $Y$-axis labels are building ID numbers. Buildings are grouped by facility. Scale bar at upper right gives number of tests. **C:** Estimated incidence by day by building, colored by room type. **D:** Incidence by room type and outbreak. **E:** Estimated effective $R$ by day per building. **F:** Distribution of effective $R$ in each room type.

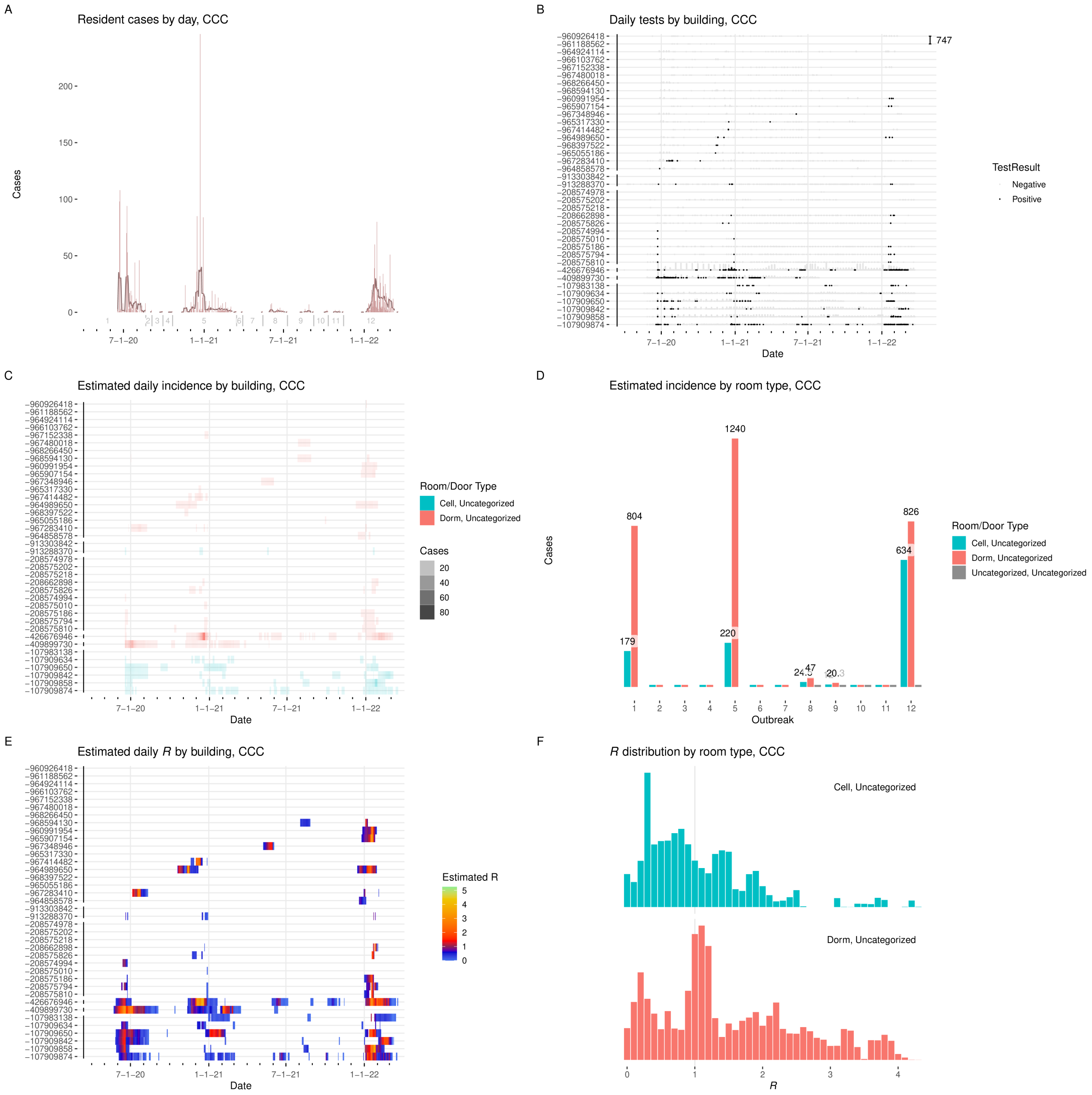

Figure I.4: **Results from CCC. A:** Cases detected per day, with seven-day average. Starting date of each outbreak is marked by a vertical gray line. **B:** Positive and negative tests per day by building. $Y$-axis labels are building ID numbers. Buildings are grouped by facility. Scale bar at upper right gives number of tests. **C:** Estimated incidence by day by building, colored by room type. **D:** Incidence by room type and outbreak. **E:** Estimated effective $R$ by day per building. **F:** Distribution of effective $R$ in each room type.

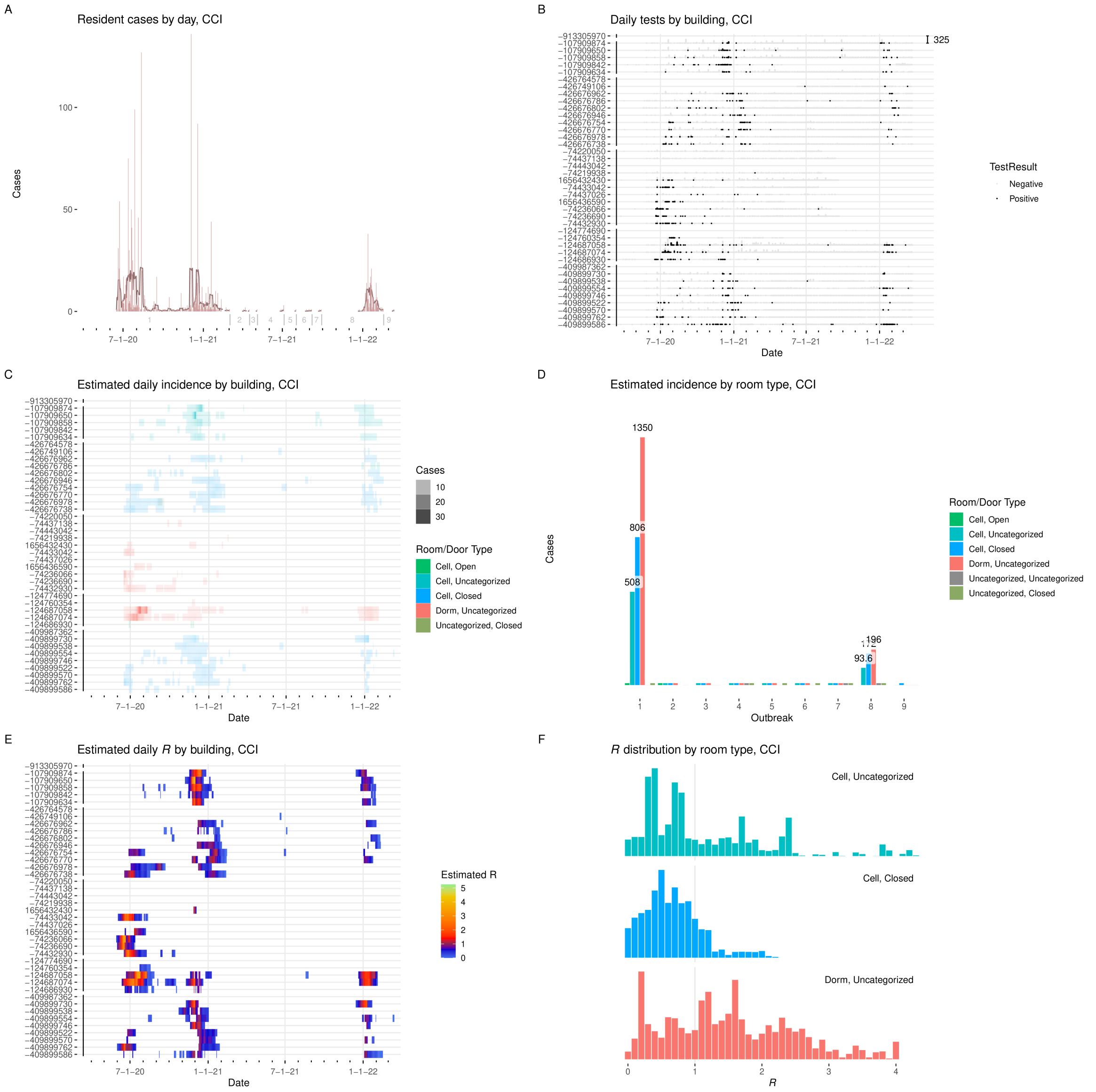


Figure I.5: **Results from CCI. A:** Cases detected per day, with seven-day average. Starting date of each outbreak is marked by a vertical gray line. **B:** Positive and negative tests per day by building. $Y$-axis labels are building ID numbers. Buildings are grouped by facility. Scale bar at upper right gives number of tests. **C:** Estimated incidence by day by building, colored by room type. **D:** Incidence by room type and outbreak. **E:** Estimated effective $R$ by day per building. **F:** Distribution of effective $R$ in each room type.

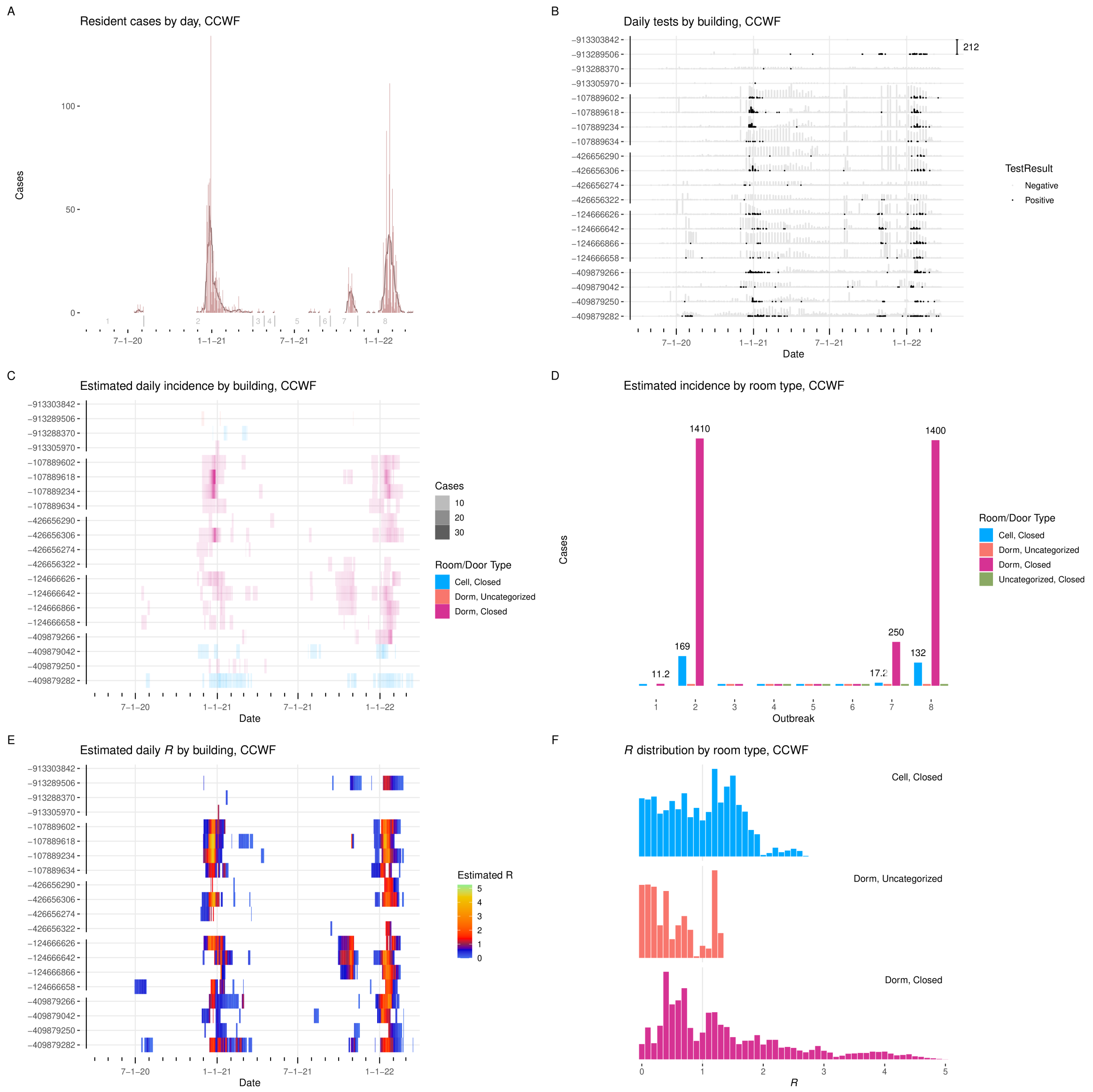


Figure I.6: **Results from CCWF. A:** Cases detected per day, with seven-day average. Starting date of each outbreak is marked by a vertical gray line. **B:** Positive and negative tests per day by building. $Y$-axis labels are building ID numbers. Buildings are grouped by facility. Scale bar at upper right gives number of tests. **C:** Estimated incidence by day by building, colored by room type. **D:** Incidence by room type and outbreak. **E:** Estimated effective $R$ by day per building. **F:** Distribution of effective $R$ in each room type.

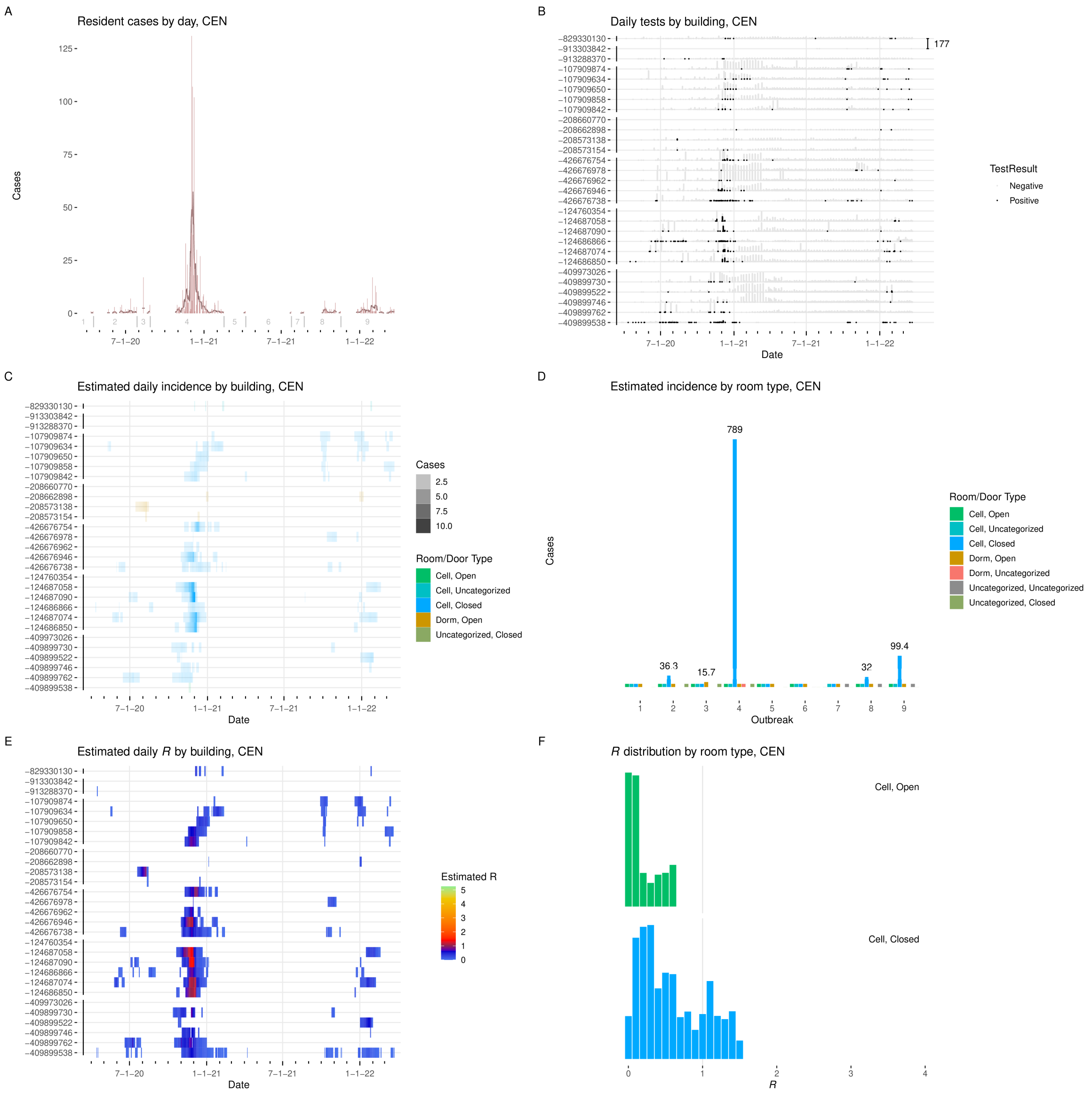


Figure I.7: **Results from CEN. A:** Cases detected per day, with seven-day average. Starting date of each outbreak is marked by a vertical gray line. **B:** Positive and negative tests per day by building. $Y$-axis labels are building ID numbers. Buildings are grouped by facility. Scale bar at upper right gives number of tests. **C:** Estimated incidence by day by building, colored by room type. **D:** Incidence by room type and outbreak. **E:** Estimated effective $R$ by day per building. **F:** Distribution of effective $R$ in each room type.

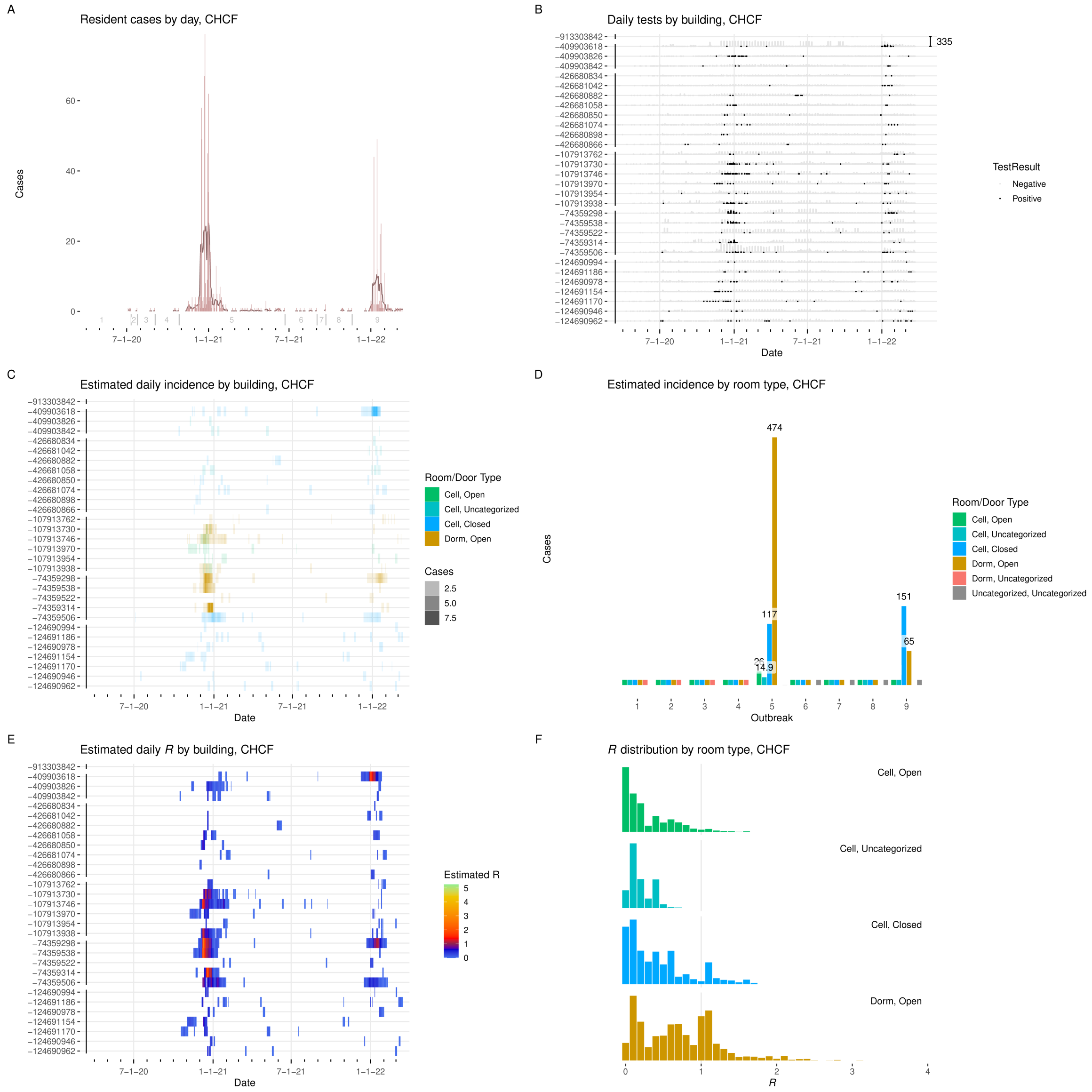


Figure I.8: **Results from CHCF. A:** Cases detected per day, with seven-day average. Starting date of each outbreak is marked by a vertical gray line. **B:** Positive and negative tests per day by building. $Y$-axis labels are building ID numbers. Buildings are grouped by facility. Scale bar at upper right gives number of tests. **C:** Estimated incidence by day by building, colored by room type. **D:** Incidence by room type and outbreak. **E:** Estimated effective $R$ by day per building. **F:** Distribution of effective $R$ in each room type.

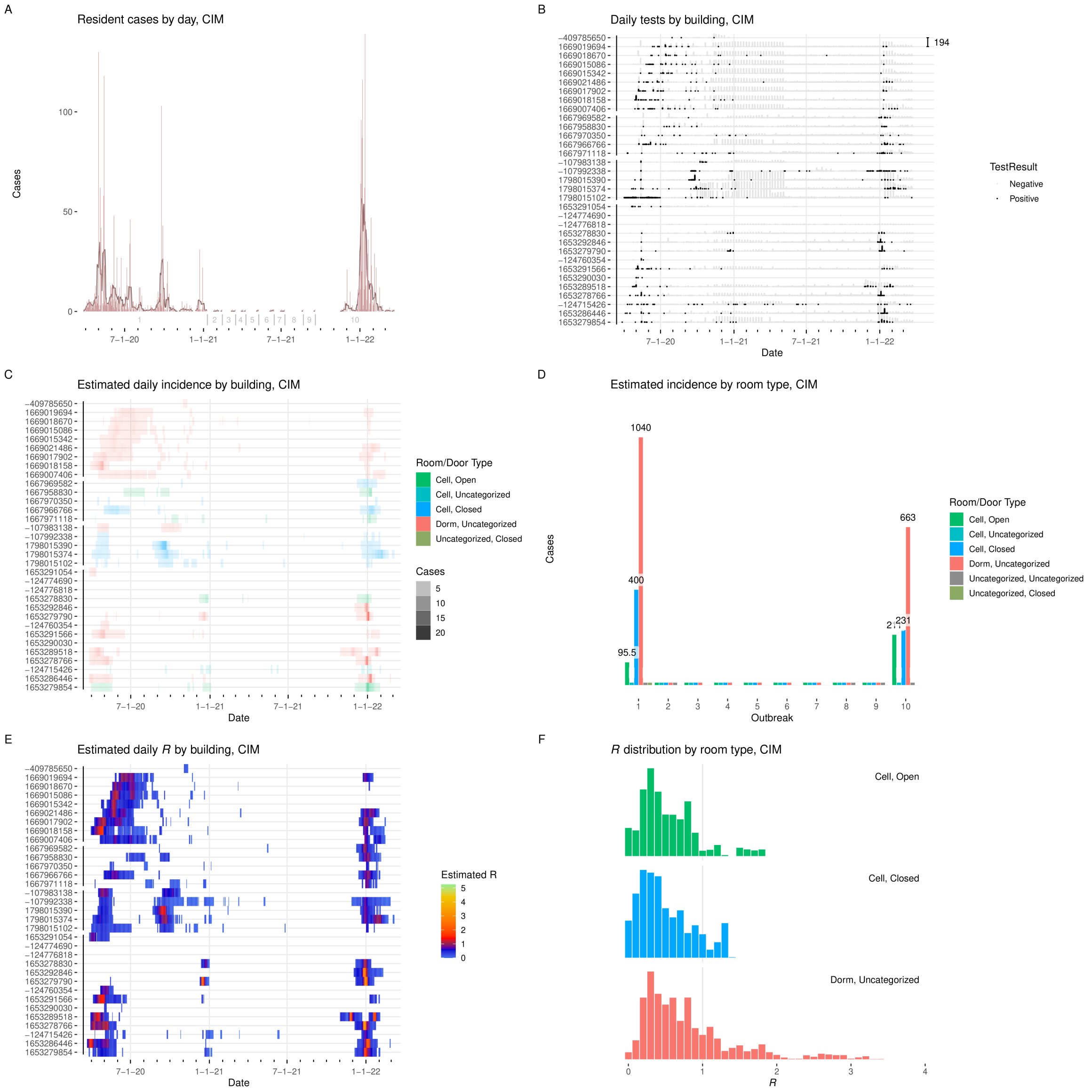


Figure I.9: **Results from CIM. A:** Cases detected per day, with seven-day average. Starting date of each outbreak is marked by a vertical gray line. **B:** Positive and negative tests per day by building. $Y$-axis labels are building ID numbers. Buildings are grouped by facility. Scale bar at upper right gives number of tests. **C:** Estimated incidence by day by building, colored by room type. **D:** Incidence by room type and outbreak. **E:** Estimated effective $R$ by day per building. **F:** Distribution of effective $R$ in each room type.

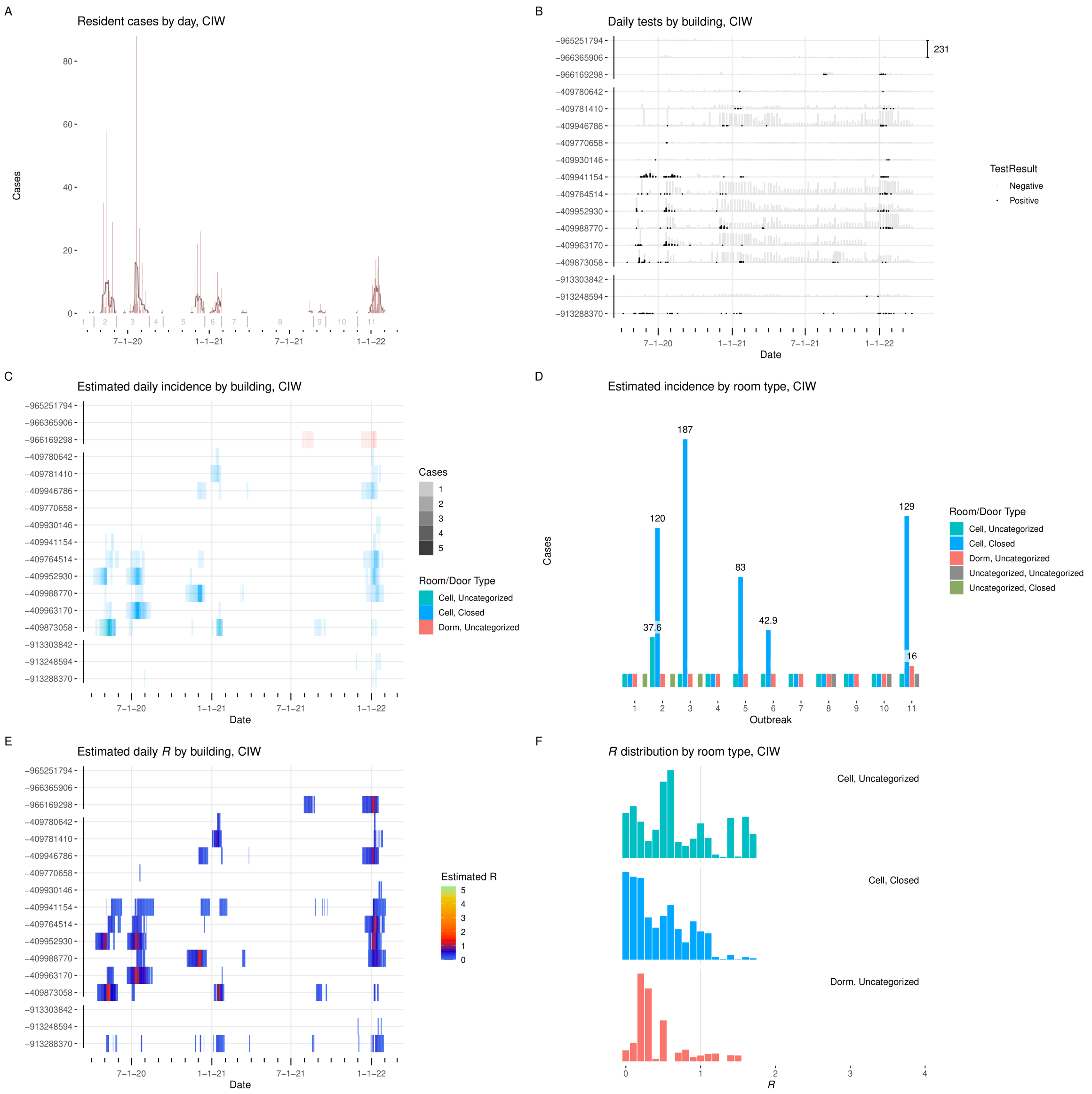


Figure I.10: **Results from CIW. A:** Cases detected per day, with seven-day average. Starting date of each outbreak is marked by a vertical gray line. **B:** Positive and negative tests per day by building. $Y$-axis labels are building ID numbers. Buildings are grouped by facility. Scale bar at upper right gives number of tests. **C:** Estimated incidence by day by building, colored by room type. **D:** Incidence by room type and outbreak. **E:** Estimated effective $R$ by day per building. **F:** Distribution of effective $R$ in each room type.

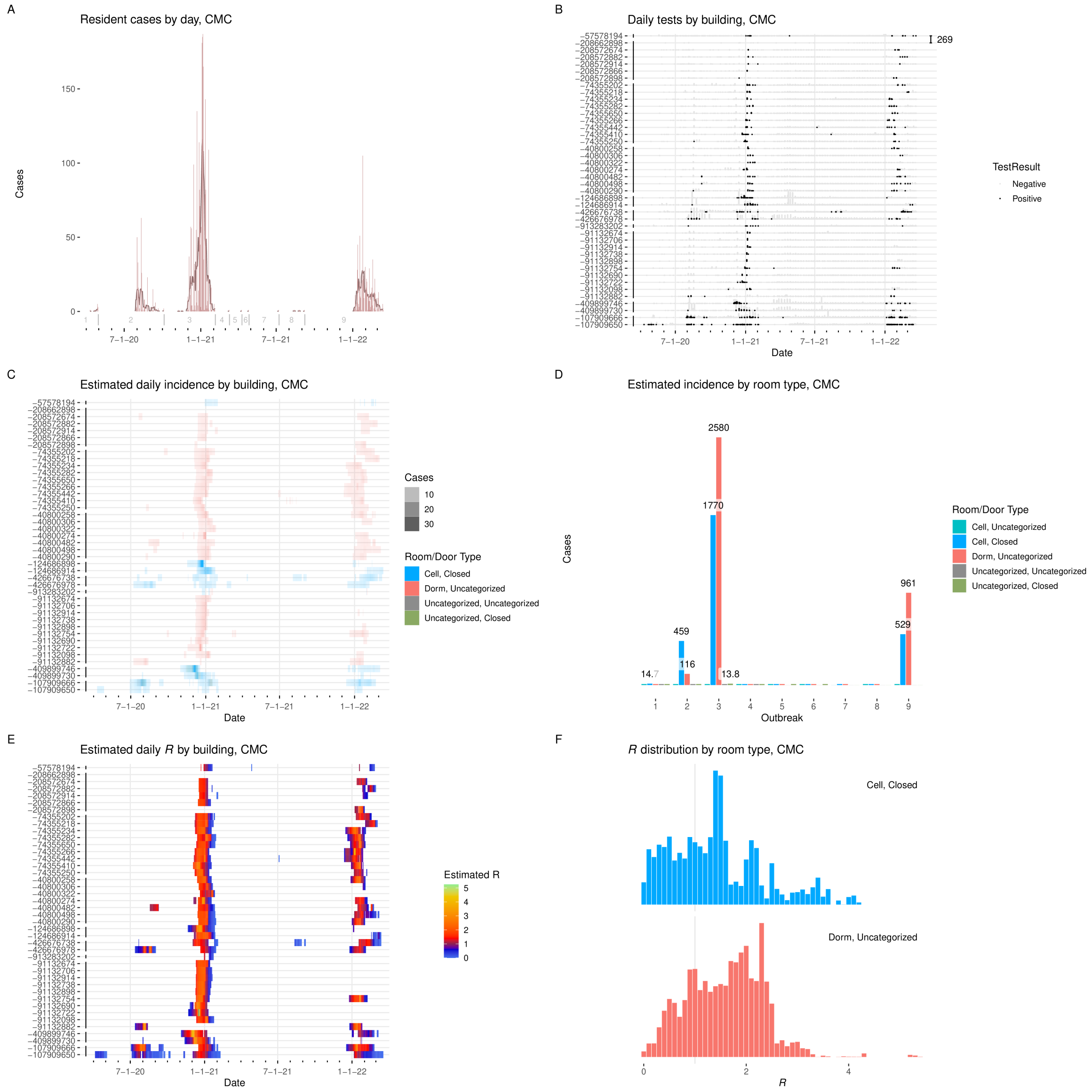


Figure I.11: **Results from CMC. A:** Cases detected per day, with seven-day average. Starting date of each outbreak is marked by a vertical gray line. **B:** Positive and negative tests per day by building. $Y$-axis labels are building ID numbers. Buildings are grouped by facility. Scale bar at upper right gives number of tests. **C:** Estimated incidence by day by building, colored by room type. **D:** Incidence by room type and outbreak. **E:** Estimated effective $R$ by day per building. **F:** Distribution of effective $R$ in each room type.

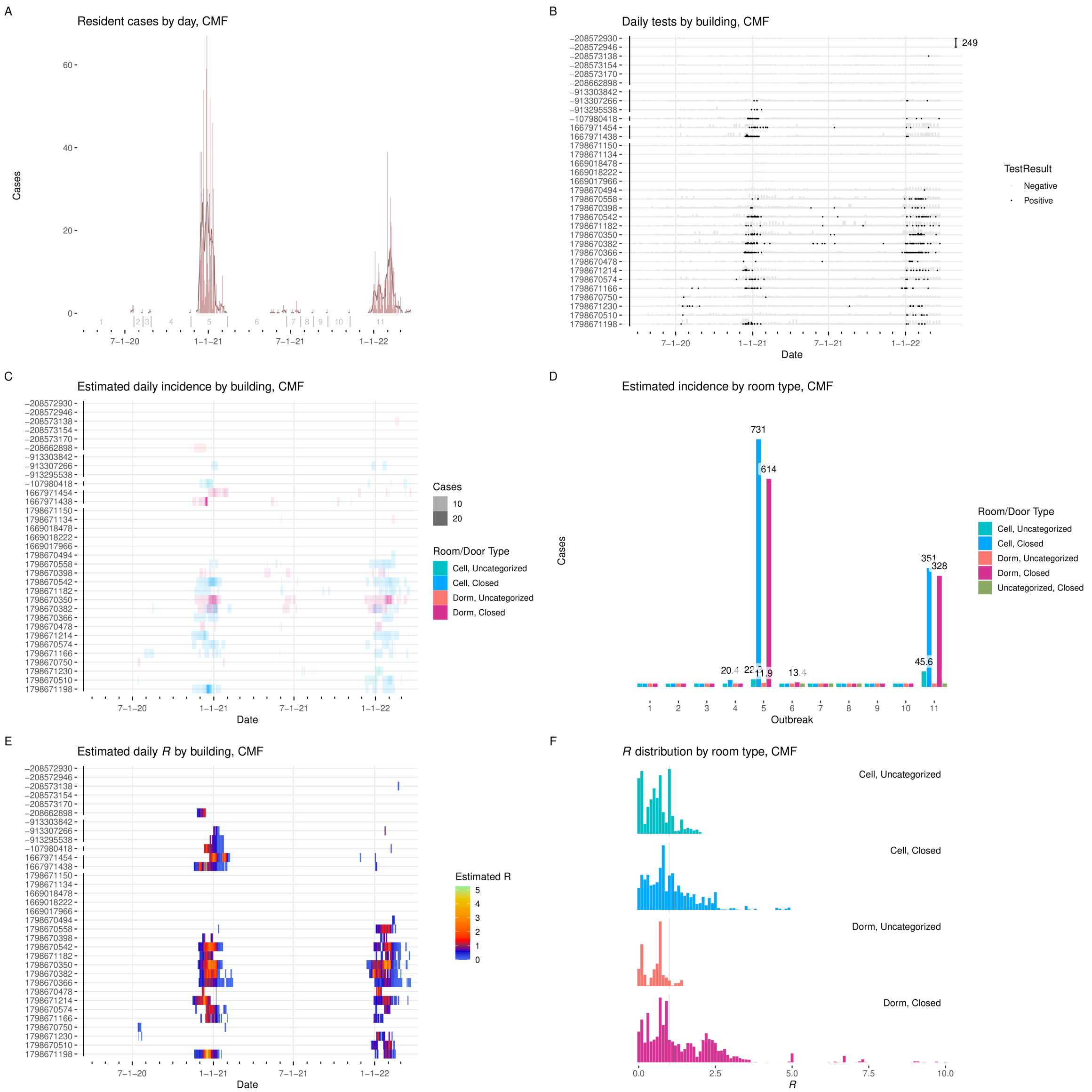

Figure I.12: **Results from CMF. A:** Cases detected per day, with seven-day average. Starting date of each outbreak is marked by a vertical gray line. **B:** Positive and negative tests per day by building. $Y$-axis labels are building ID numbers. Buildings are grouped by facility. Scale bar at upper right gives number of tests. **C:** Estimated incidence by day by building, colored by room type. **D:** Incidence by room type and outbreak. **E:** Estimated effective $R$ by day per building. **F:** Distribution of effective $R$ in each room type.

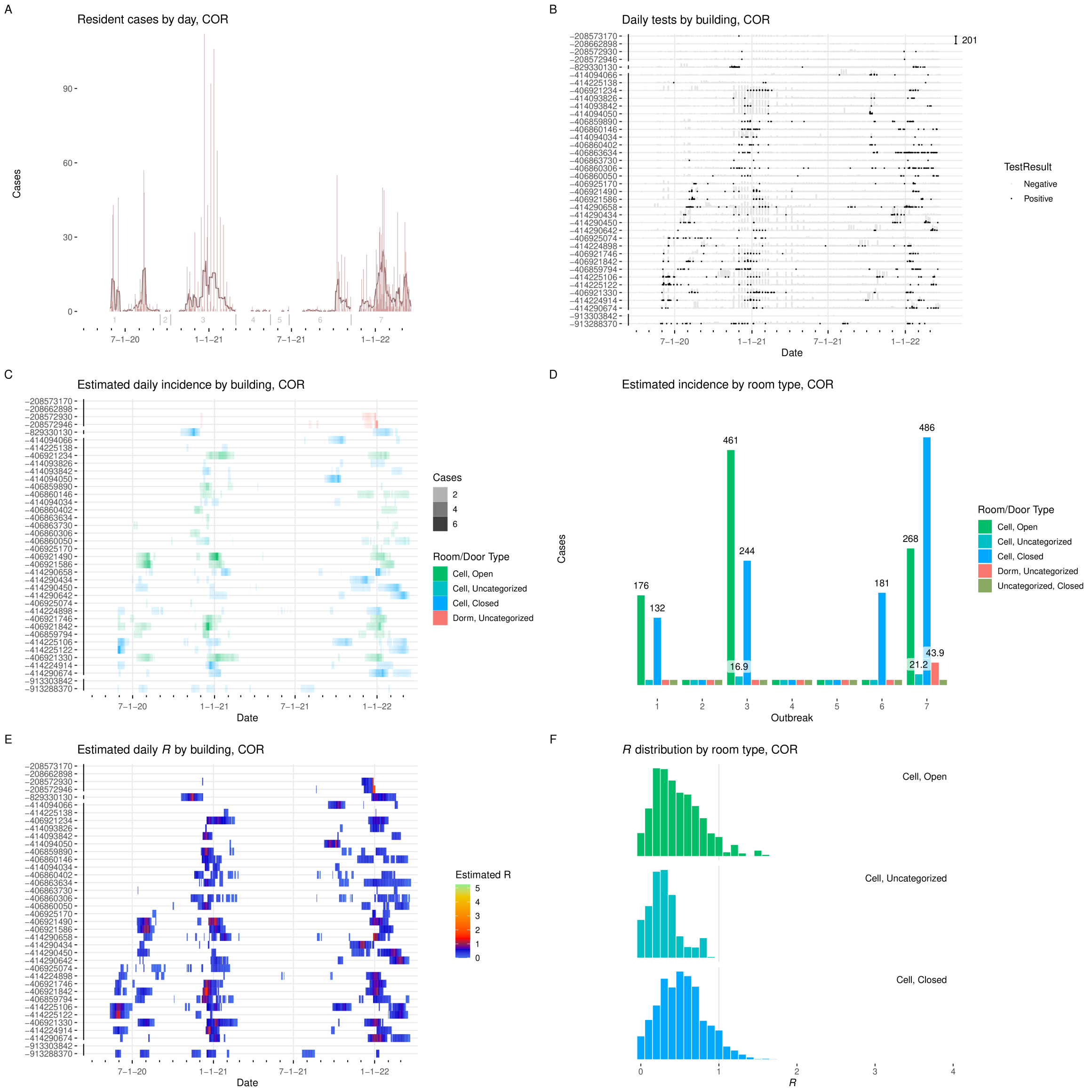


Figure I.13: **Results from COR. A:** Cases detected per day, with seven-day average. Starting date of each outbreak is marked by a vertical gray line. **B:** Positive and negative tests per day by building. $Y$-axis labels are building ID numbers. Buildings are grouped by facility. Scale bar at upper right gives number of tests. **C:** Estimated incidence by day by building, colored by room type. **D:** Incidence by room type and outbreak. **E:** Estimated effective $R$ by day per building. **F:** Distribution of effective $R$ in each room type.

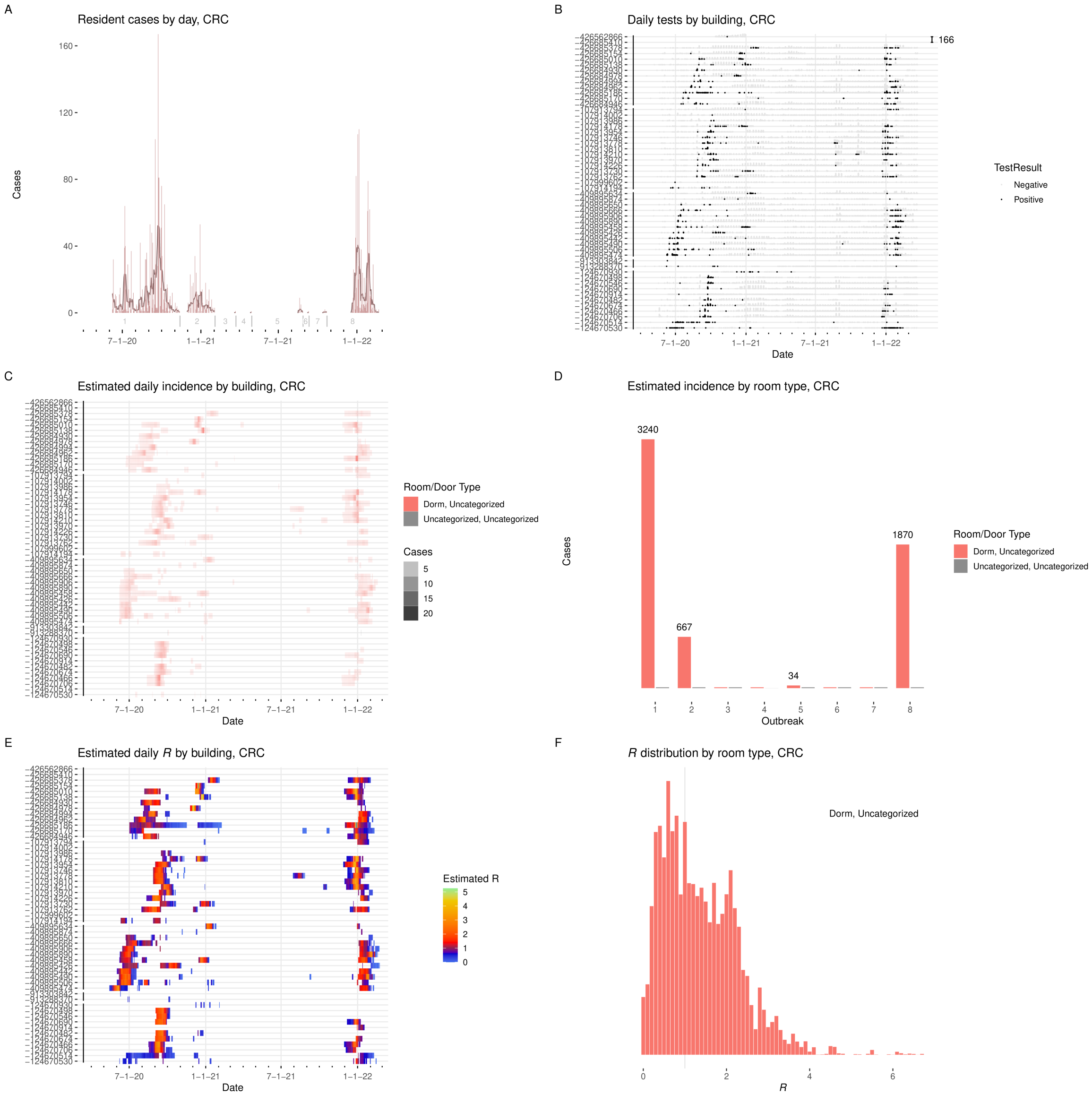


Figure I.14: **Results from CRC. A:** Cases detected per day, with seven-day average. Starting date of each outbreak is marked by a vertical gray line. **B:** Positive and negative tests per day by building. $Y$-axis labels are building ID numbers. Buildings are grouped by facility. Scale bar at upper right gives number of tests. **C:** Estimated incidence by day by building, colored by room type. **D:** Incidence by room type and outbreak. **E:** Estimated effective $R$ by day per building. **F:** Distribution of effective $R$ in each room type.

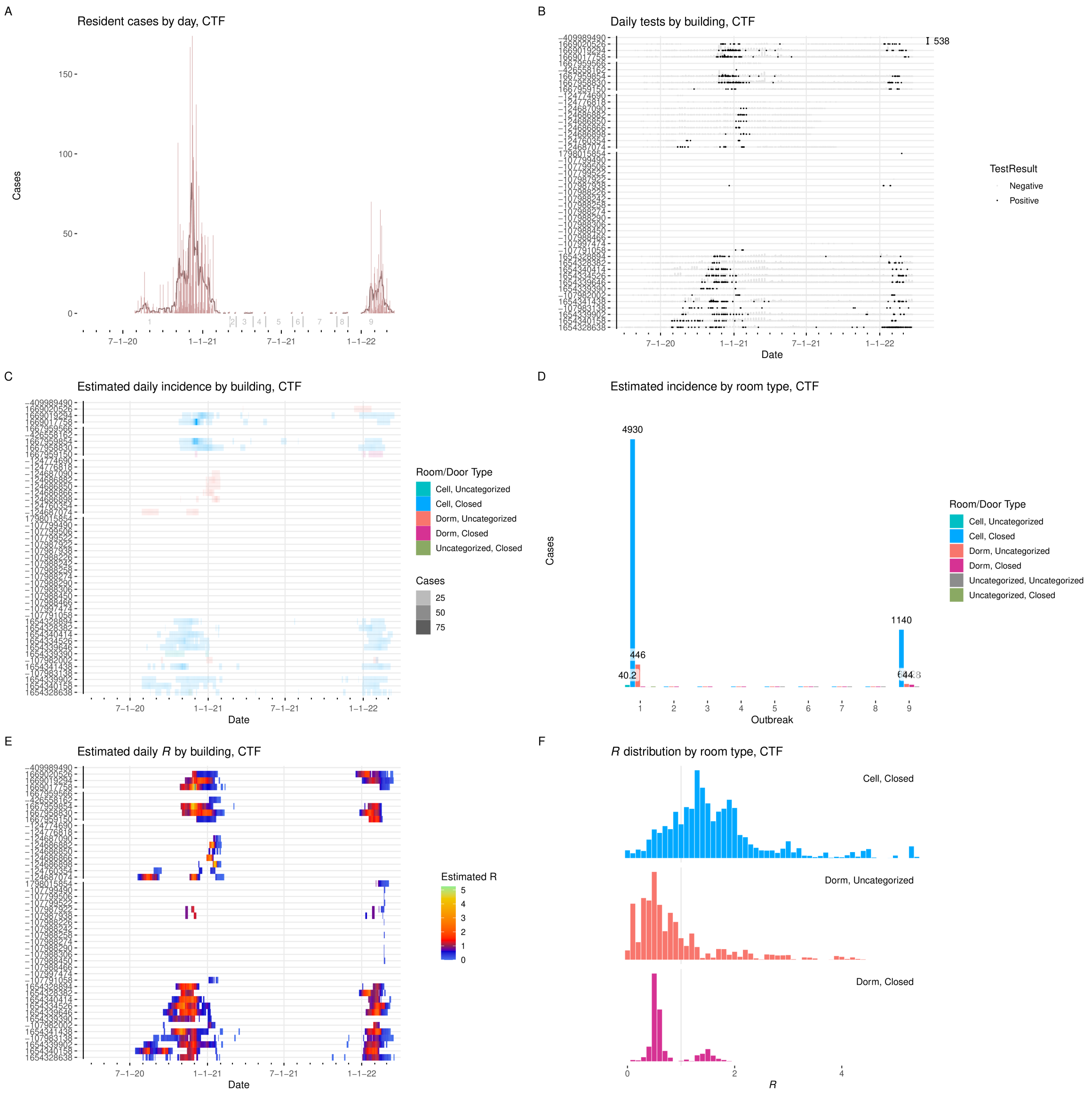


Figure I.15: **Results from CTF. A:** Cases detected per day, with seven-day average. Starting date of each outbreak is marked by a vertical gray line. **B:** Positive and negative tests per day by building. $Y$-axis labels are building ID numbers. Buildings are grouped by facility. Scale bar at upper right gives number of tests. **C:** Estimated incidence by day by building, colored by room type. **D:** Incidence by room type and outbreak. **E:** Estimated effective $R$ by day per building. **F:** Distribution of effective $R$ in each room type.

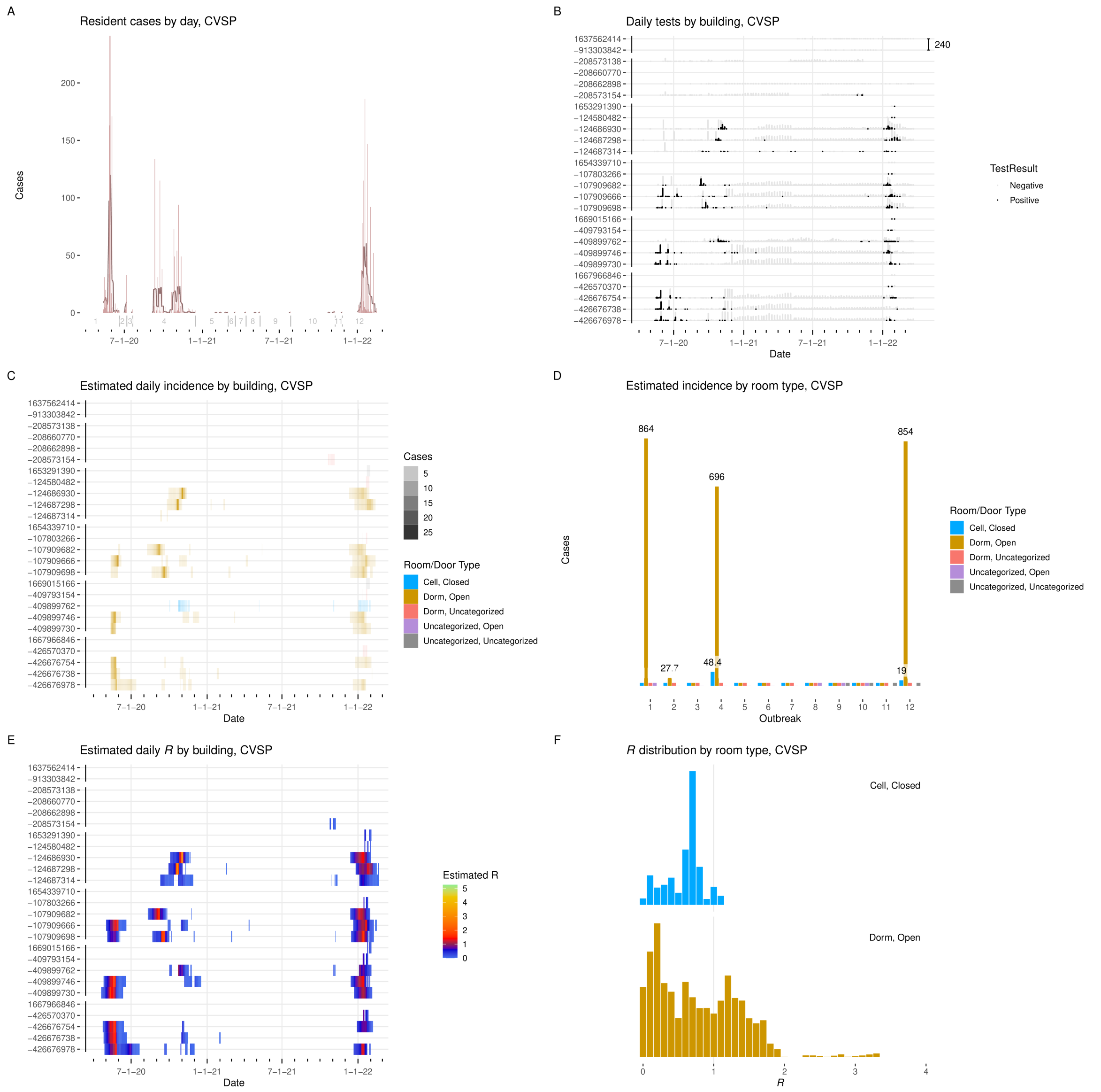


Figure I.16: **Results from CVSP. A:** Cases detected per day, with seven-day average. Starting date of each outbreak is marked by a vertical gray line. **B:** Positive and negative tests per day by building. $Y$-axis labels are building ID numbers. Buildings are grouped by facility. Scale bar at upper right gives number of tests. **C:** Estimated incidence by day by building, colored by room type. **D:** Incidence by room type and outbreak. **E:** Estimated effective $R$ by day per building. **F:** Distribution of effective $R$ in each room type.

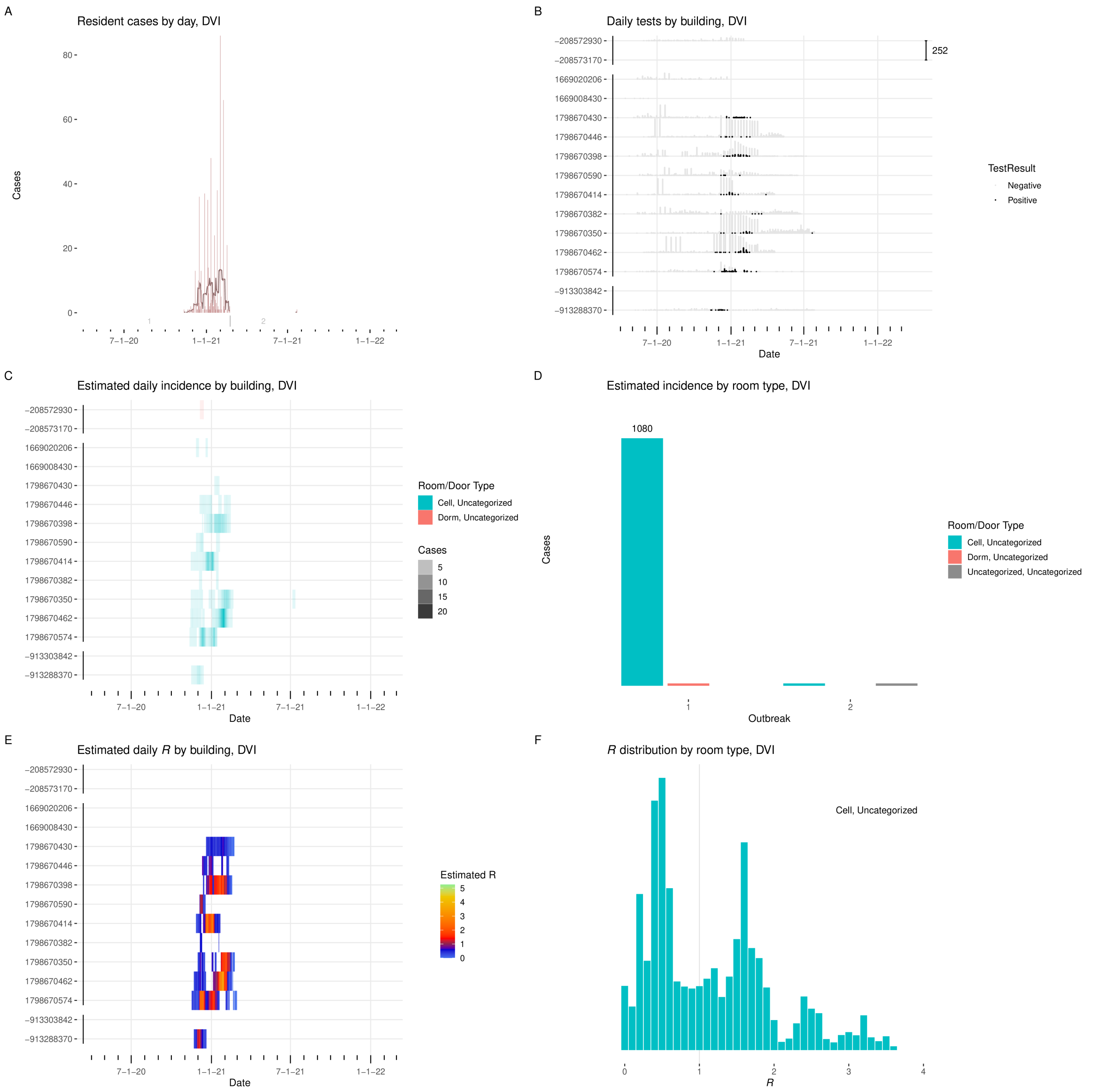


Figure I.17: **Results from DVI. A:** Cases detected per day, with seven-day average. Starting date of each outbreak is marked by a vertical gray line. **B:** Positive and negative tests per day by building. $Y$-axis labels are building ID numbers. Buildings are grouped by facility. Scale bar at upper right gives number of tests. **C:** Estimated incidence by day by building, colored by room type. **D:** Incidence by room type and outbreak. **E:** Estimated effective $R$ by day per building. **F:** Distribution of effective $R$ in each room type.

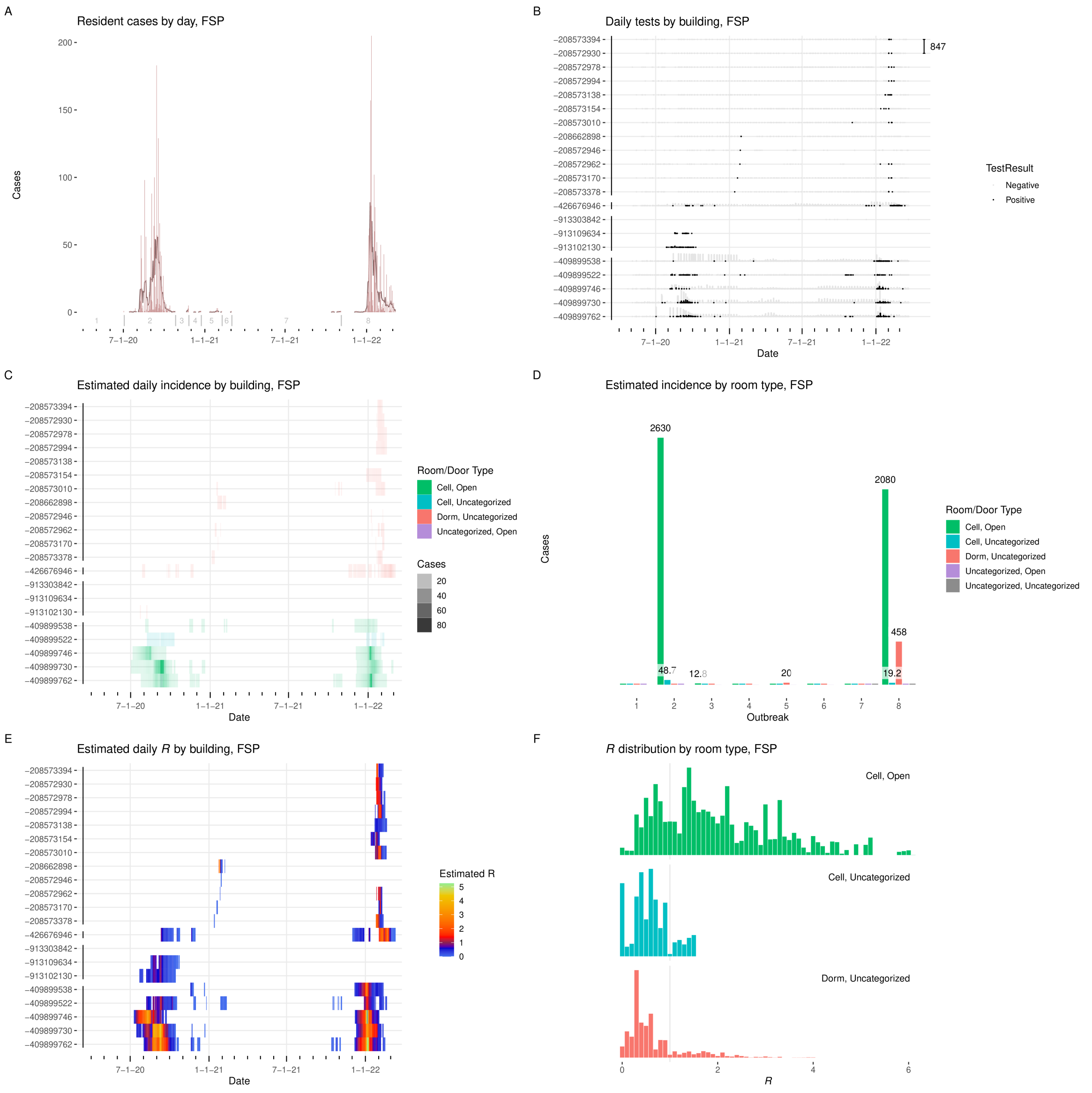


Figure I.18: **Results from FSP. A:** Cases detected per day, with seven-day average. Starting date of each outbreak is marked by a vertical gray line. **B:** Positive and negative tests per day by building. $Y$-axis labels are building ID numbers. Buildings are grouped by facility. Scale bar at upper right gives number of tests. **C:** Estimated incidence by day by building, colored by room type. **D:** Incidence by room type and outbreak. **E:** Estimated effective $R$ by day per building. **F:** Distribution of effective $R$ in each room type.

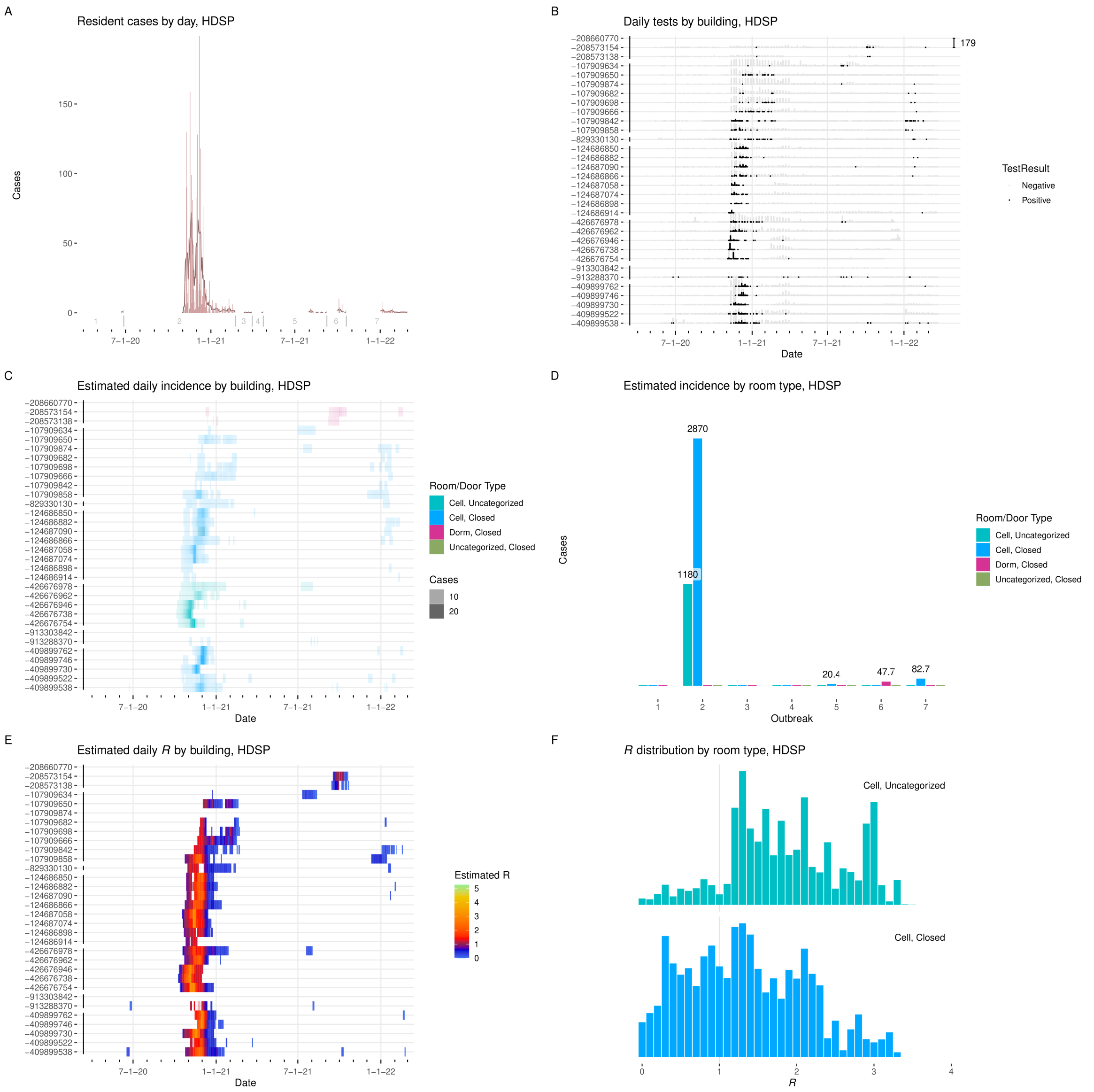


Figure I.19: **Results from HDSP. A:** Cases detected per day, with seven-day average. Starting date of each outbreak is marked by a vertical gray line. **B:** Positive and negative tests per day by building. $Y$-axis labels are building ID numbers. Buildings are grouped by facility. Scale bar at upper right gives number of tests. **C:** Estimated incidence by day by building, colored by room type. **D:** Incidence by room type and outbreak. **E:** Estimated effective $R$ by day per building. **F:** Distribution of effective $R$ in each room type.

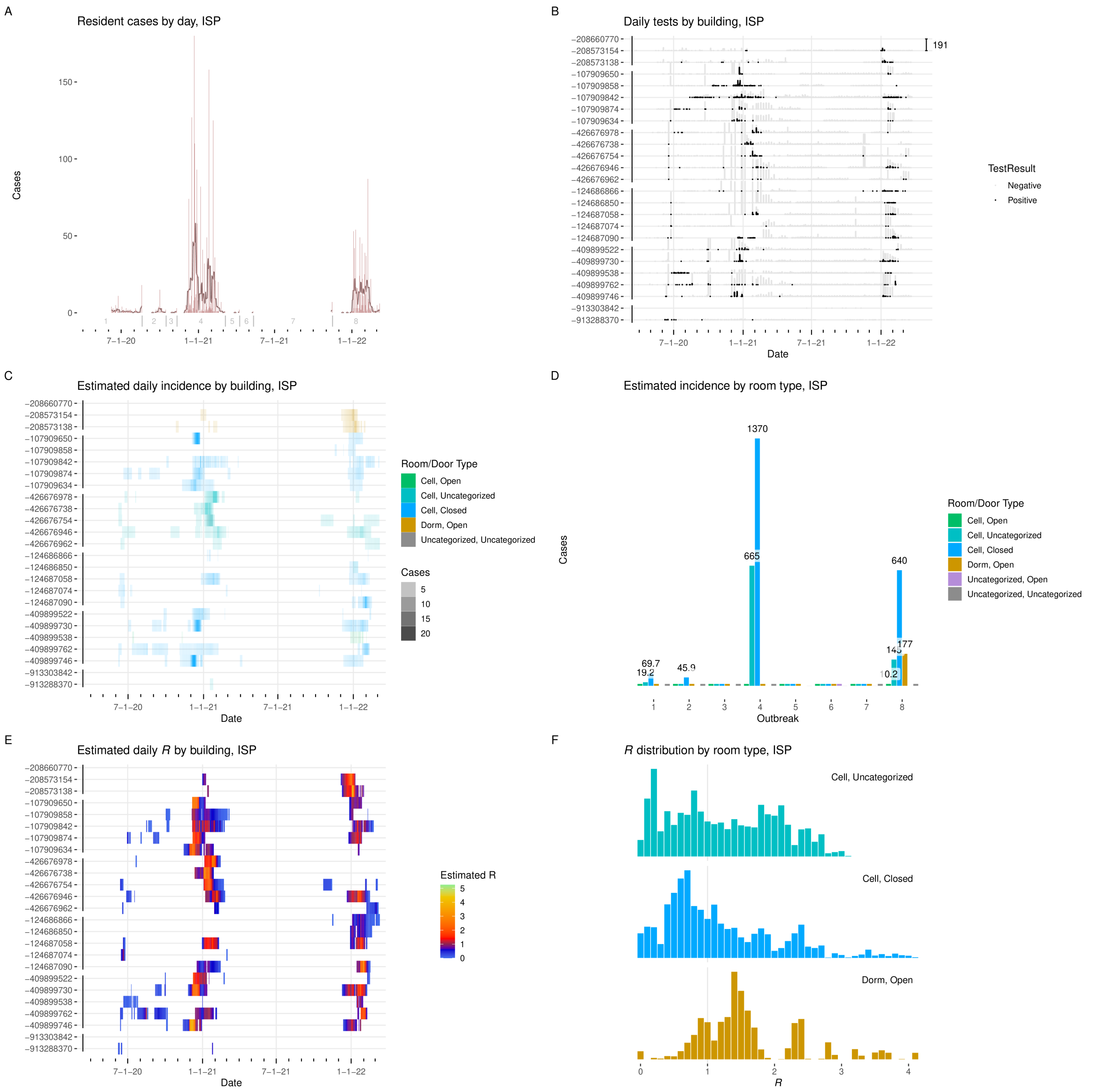


Figure I.20: **Results from ISP. A:** Cases detected per day, with seven-day average. Starting date of each outbreak is marked by a vertical gray line. **B:** Positive and negative tests per day by building. $Y$-axis labels are building ID numbers. Buildings are grouped by facility. Scale bar at upper right gives number of tests. **C:** Estimated incidence by day by building, colored by room type. **D:** Incidence by room type and outbreak. **E:** Estimated effective $R$ by day per building. **F:** Distribution of effective $R$ in each room type.

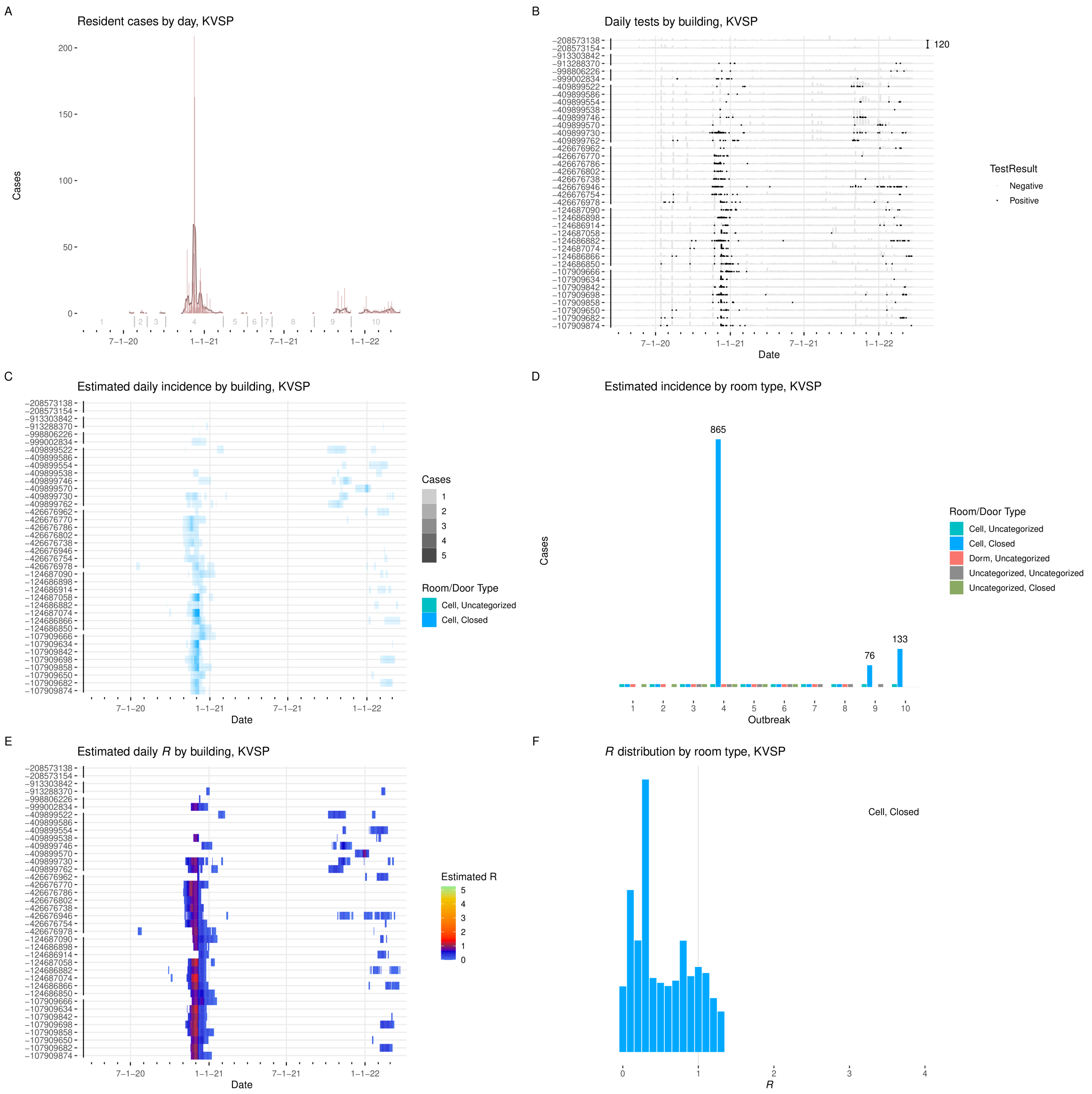


Figure I.21: **Results from KVSP. A:** Cases detected per day, with seven-day average. Starting date of each outbreak is marked by a vertical gray line. **B:** Positive and negative tests per day by building. $Y$-axis labels are building ID numbers. Buildings are grouped by facility. Scale bar at upper right gives number of tests. **C:** Estimated incidence by day by building, colored by room type. **D:** Incidence by room type and outbreak. **E:** Estimated effective $R$ by day per building. **F:** Distribution of effective $R$ in each room type.

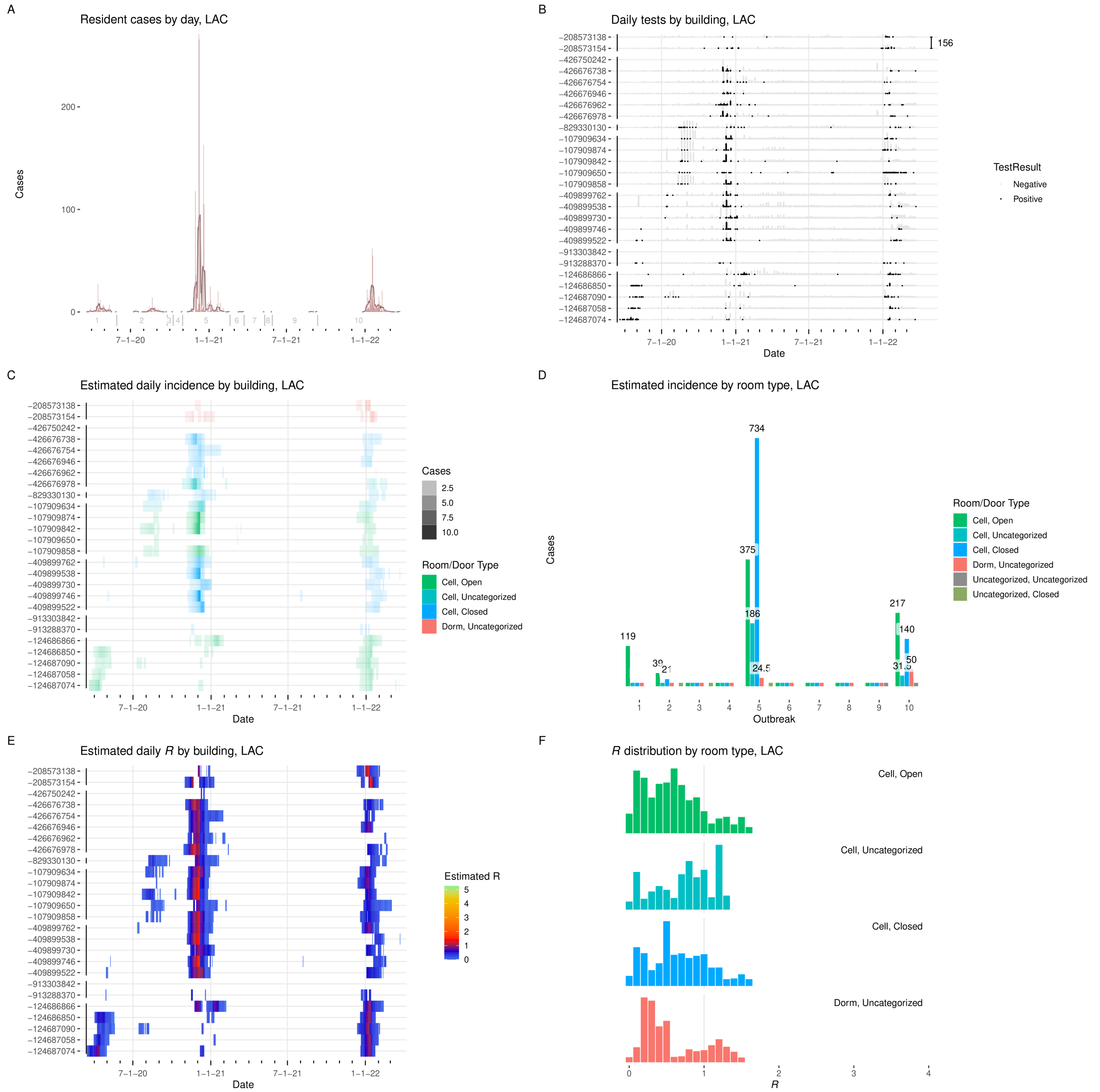


Figure I.22: **Results from LAC. A:** Cases detected per day, with seven-day average. Starting date of each outbreak is marked by a vertical gray line. **B:** Positive and negative tests per day by building. $Y$-axis labels are building ID numbers. Buildings are grouped by facility. Scale bar at upper right gives number of tests. **C:** Estimated incidence by day by building, colored by room type. **D:** Incidence by room type and outbreak. **E:** Estimated effective $R$ by day per building. **F:** Distribution of effective $R$ in each room type.

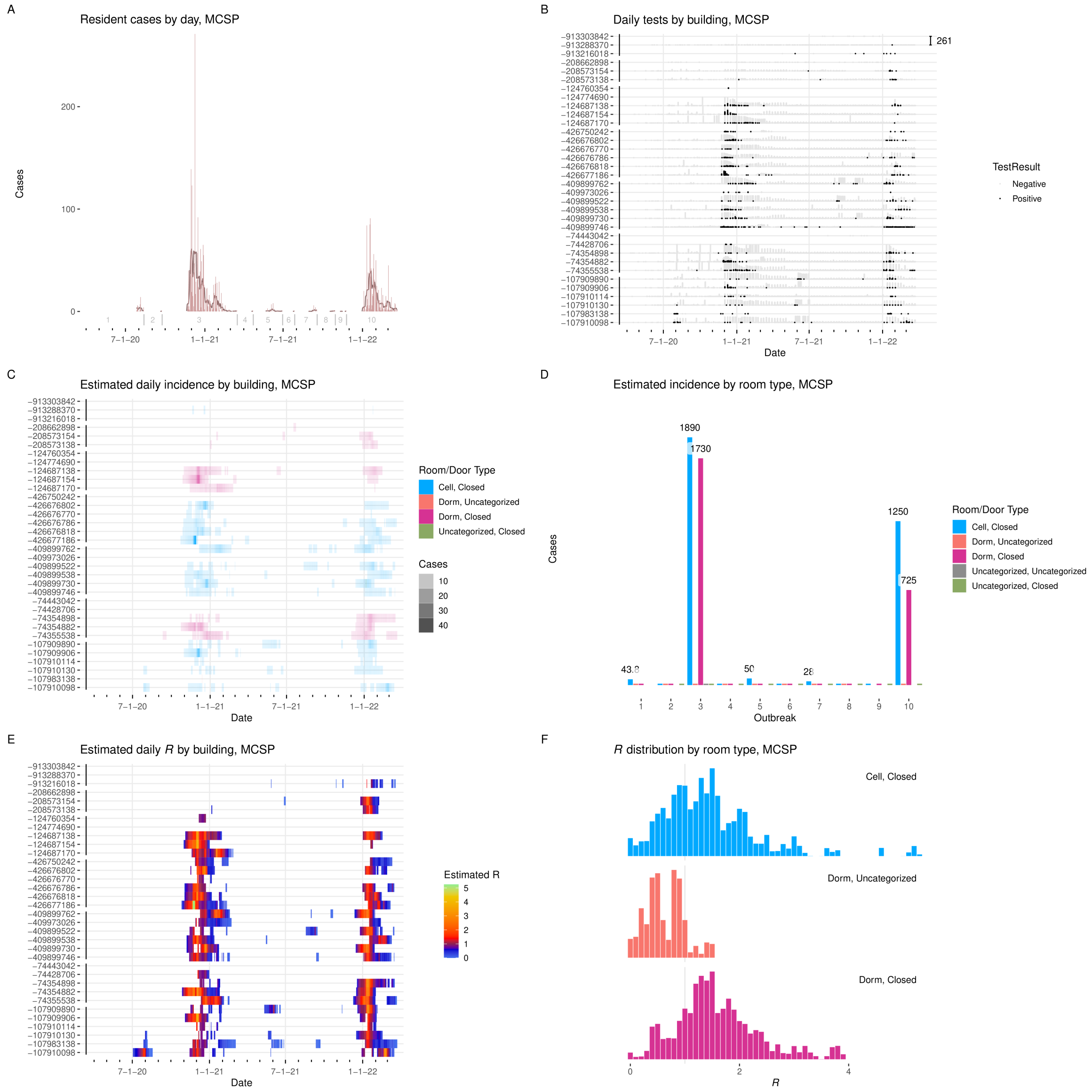


Figure I.23: **Results from MCSP. A:** Cases detected per day, with seven-day average. Starting date of each outbreak is marked by a vertical gray line. **B:** Positive and negative tests per day by building. $Y$-axis labels are building ID numbers. Buildings are grouped by facility. Scale bar at upper right gives number of tests. **C:** Estimated incidence by day by building, colored by room type. **D:** Incidence by room type and outbreak. **E:** Estimated effective $R$ by day per building. **F:** Distribution of effective $R$ in each room type.

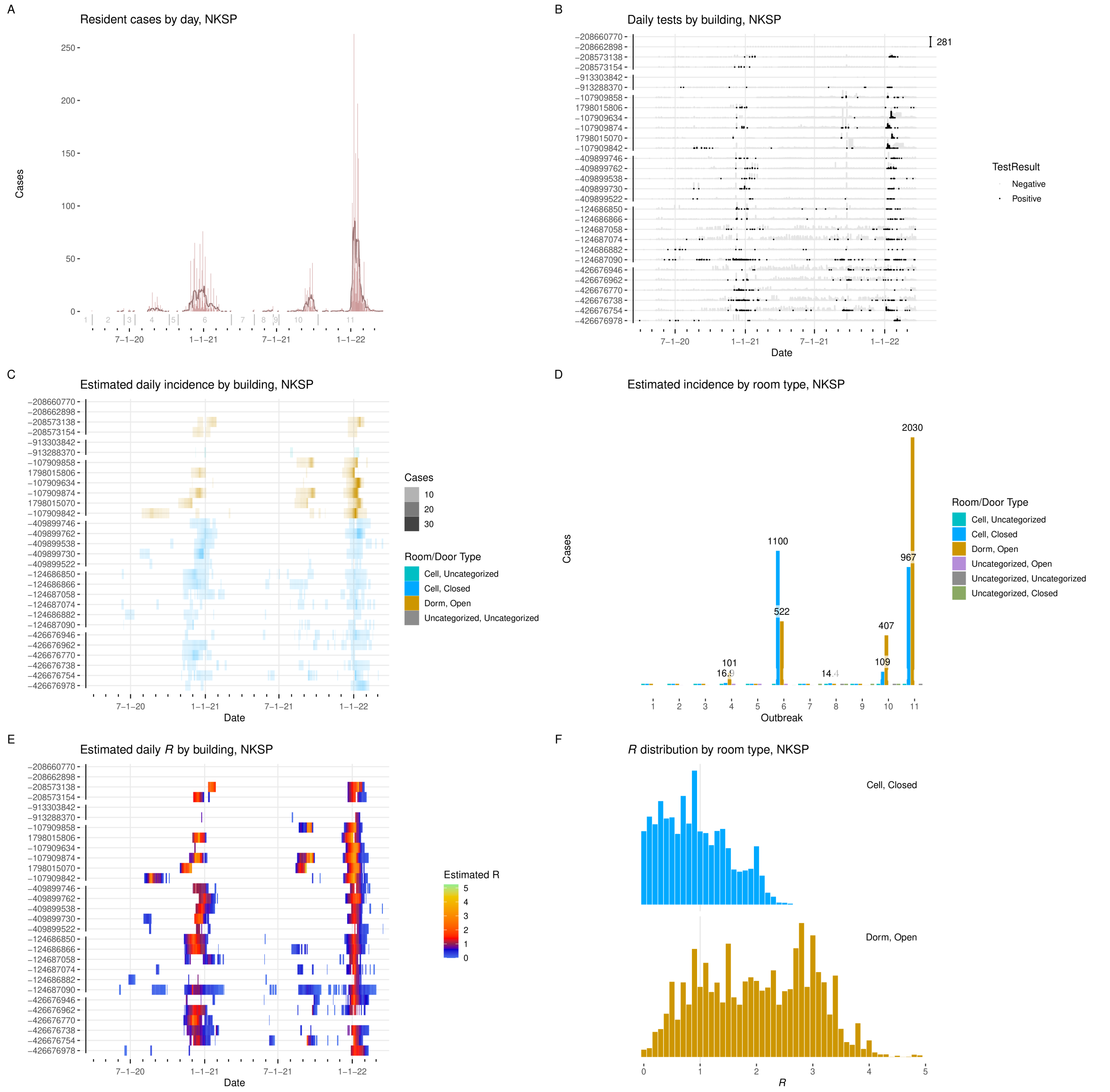


Figure I.24: **Results from NKSP. A:** Cases detected per day, with seven-day average. Starting date of each outbreak is marked by a vertical gray line. **B:** Positive and negative tests per day by building. $Y$-axis labels are building ID numbers. Buildings are grouped by facility. Scale bar at upper right gives number of tests. **C:** Estimated incidence by day by building, colored by room type. **D:** Incidence by room type and outbreak. **E:** Estimated effective $R$ by day per building. **F:** Distribution of effective $R$ in each room type.

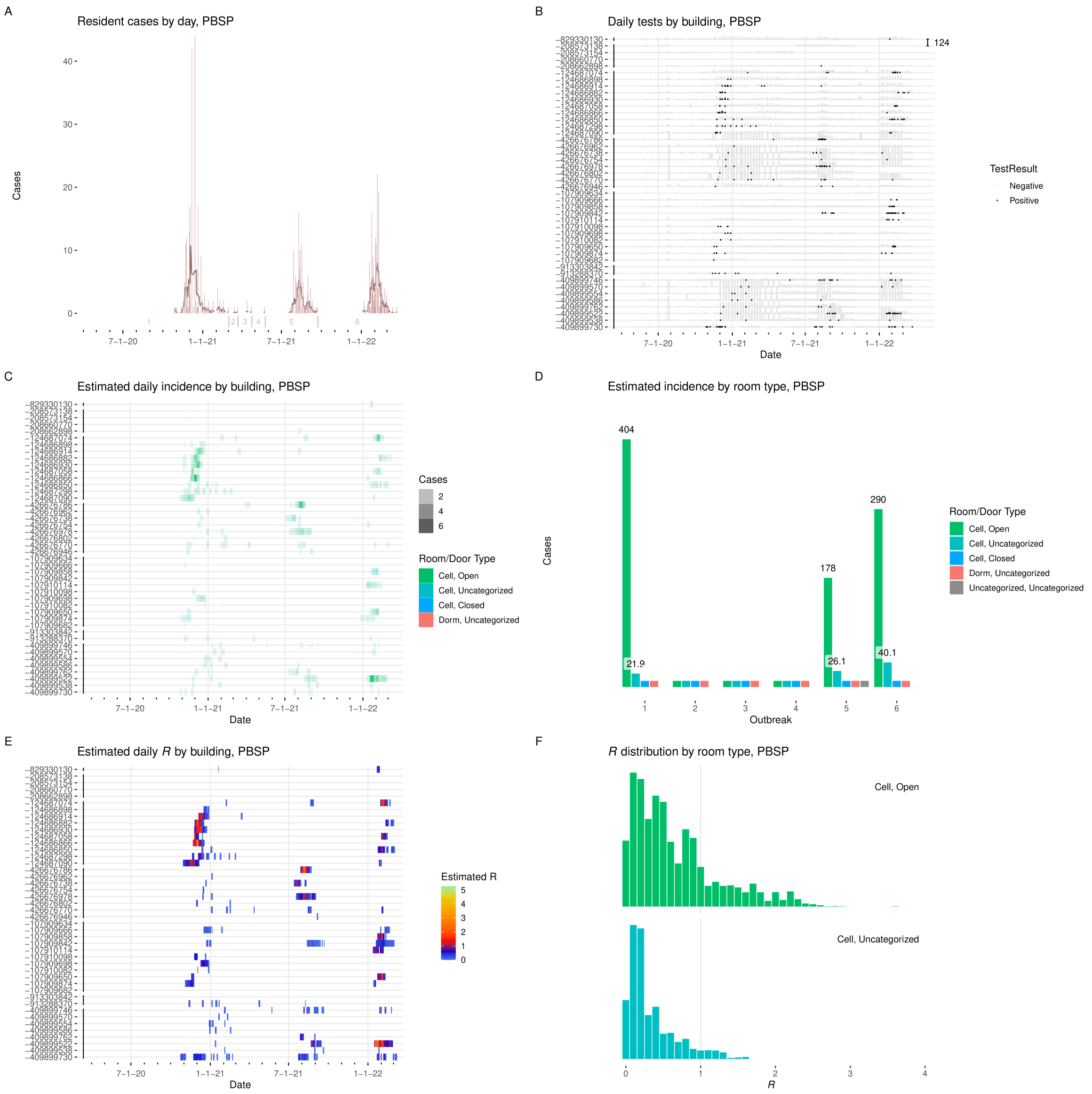


Figure I.25: **Results from PBSP. A:** Cases detected per day, with seven-day average. Starting date of each outbreak is marked by a vertical gray line. **B:** Positive and negative tests per day by building. $Y$-axis labels are building ID numbers. Buildings are grouped by facility. Scale bar at upper right gives number of tests. **C:** Estimated incidence by day by building, colored by room type. **D:** Incidence by room type and outbreak. **E:** Estimated effective $R$ by day per building. **F:** Distribution of effective $R$ in each room type.

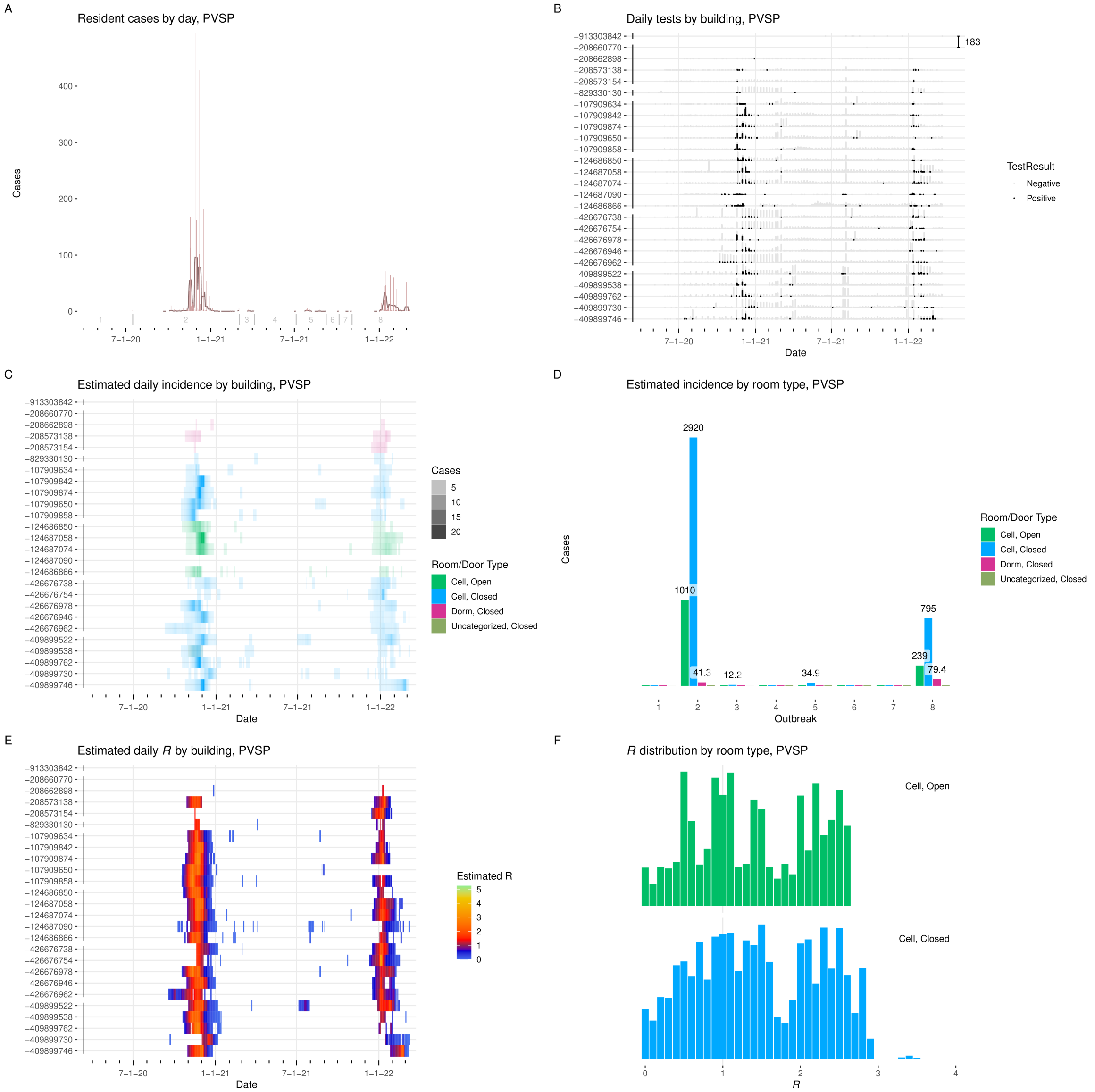


Figure I.26: **Results from PVSP. A:** Cases detected per day, with seven-day average. Starting date of each outbreak is marked by a vertical gray line. **B:** Positive and negative tests per day by building. $Y$-axis labels are building ID numbers. Buildings are grouped by facility. Scale bar at upper right gives number of tests. **C:** Estimated incidence by day by building, colored by room type. **D:** Incidence by room type and outbreak. **E:** Estimated effective $R$ by day per building. **F:** Distribution of effective $R$ in each room type.

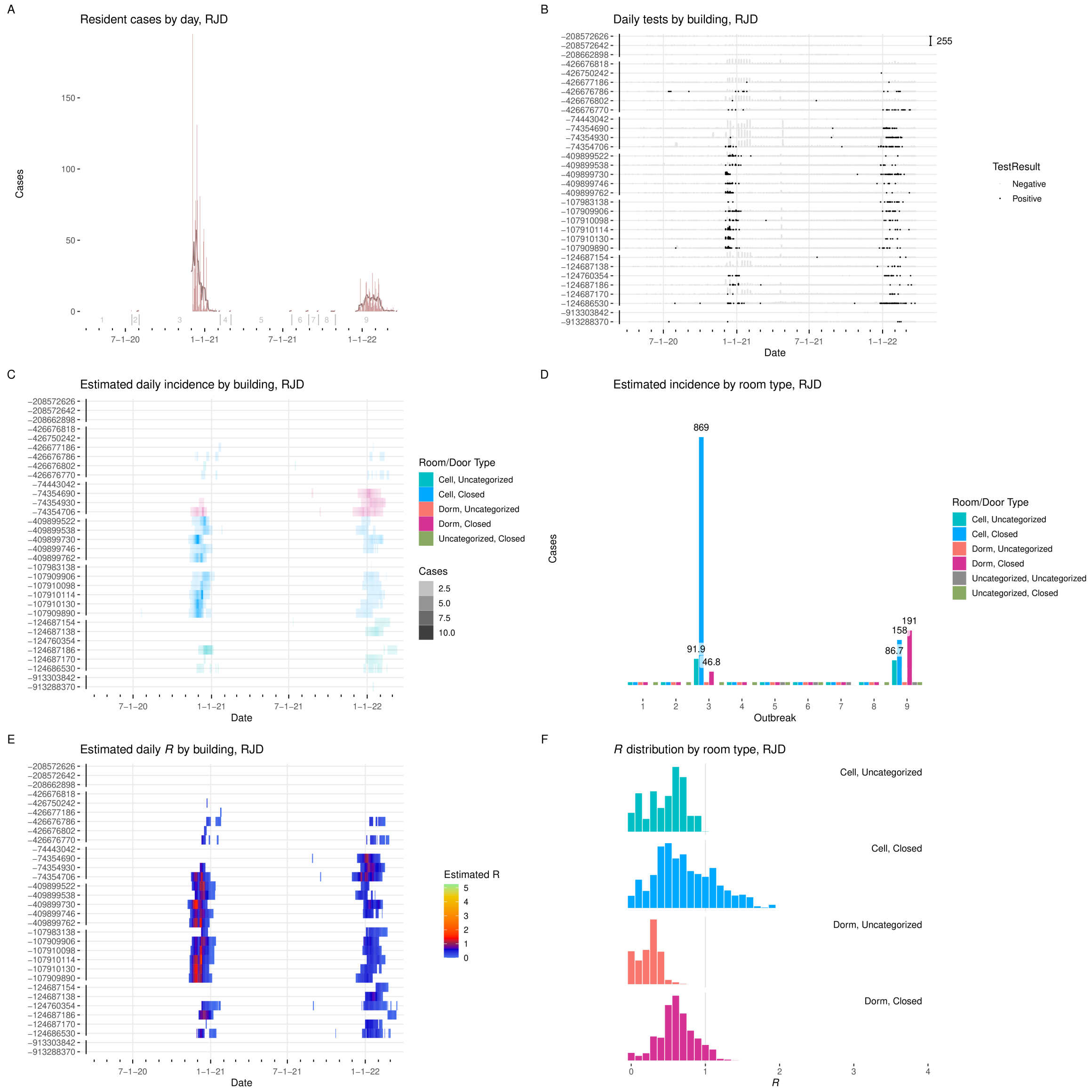


Figure I.27: **Results from RJD. A:** Cases detected per day, with seven-day average. Starting date of each outbreak is marked by a vertical gray line. **B:** Positive and negative tests per day by building. $Y$-axis labels are building ID numbers. Buildings are grouped by facility. Scale bar at upper right gives number of tests. **C:** Estimated incidence by day by building, colored by room type. **D:** Incidence by room type and outbreak. **E:** Estimated effective $R$ by day per building. **F:** Distribution of effective $R$ in each room type.

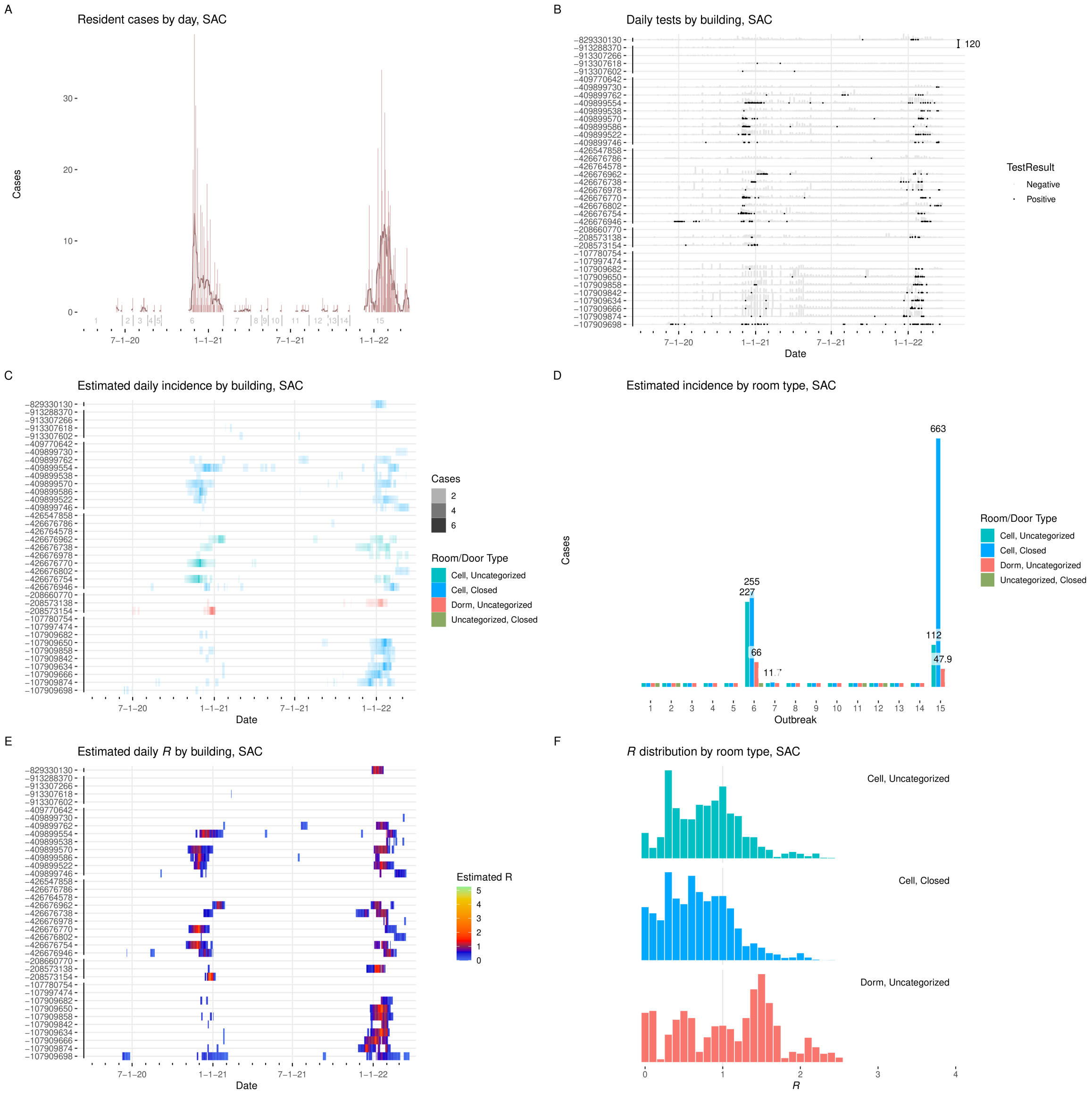


Figure I.28: **Results from SAC. A:** Cases detected per day, with seven-day average. Starting date of each outbreak is marked by a vertical gray line. **B:** Positive and negative tests per day by building. $Y$-axis labels are building ID numbers. Buildings are grouped by facility. Scale bar at upper right gives number of tests. **C:** Estimated incidence by day by building, colored by room type. **D:** Incidence by room type and outbreak. **E:** Estimated effective $R$ by day per building. **F:** Distribution of effective $R$ in each room type.

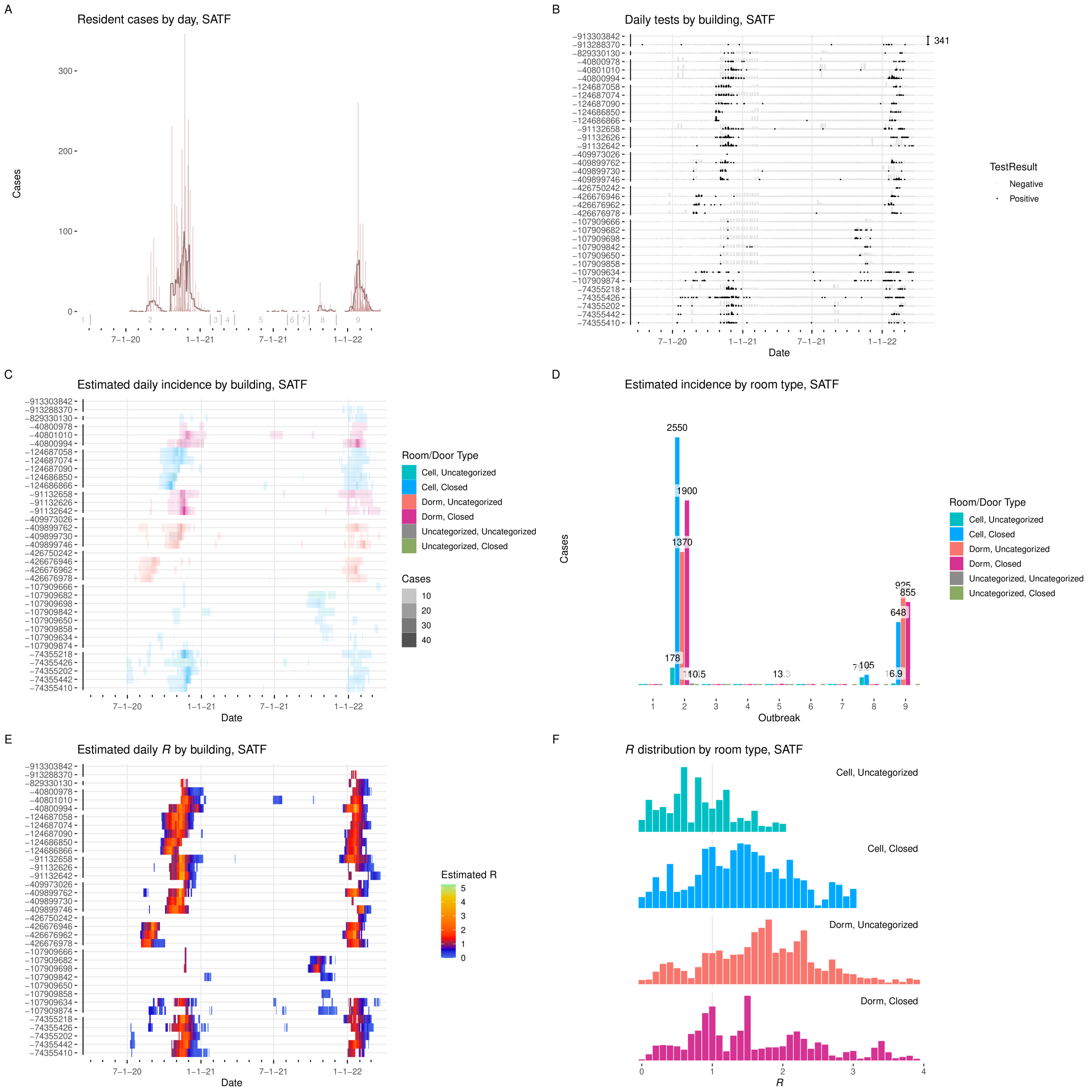

Figure I.29: **Results from SATF. A:** Cases detected per day, with seven-day average. Starting date of each outbreak is marked by a vertical gray line. **B:** Positive and negative tests per day by building. $Y$-axis labels are building ID numbers. Buildings are grouped by facility. Scale bar at upper right gives number of tests. **C:** Estimated incidence by day by building, colored by room type. **D:** Incidence by room type and outbreak. **E:** Estimated effective $R$ by day per building. **F:** Distribution of effective $R$ in each room type.

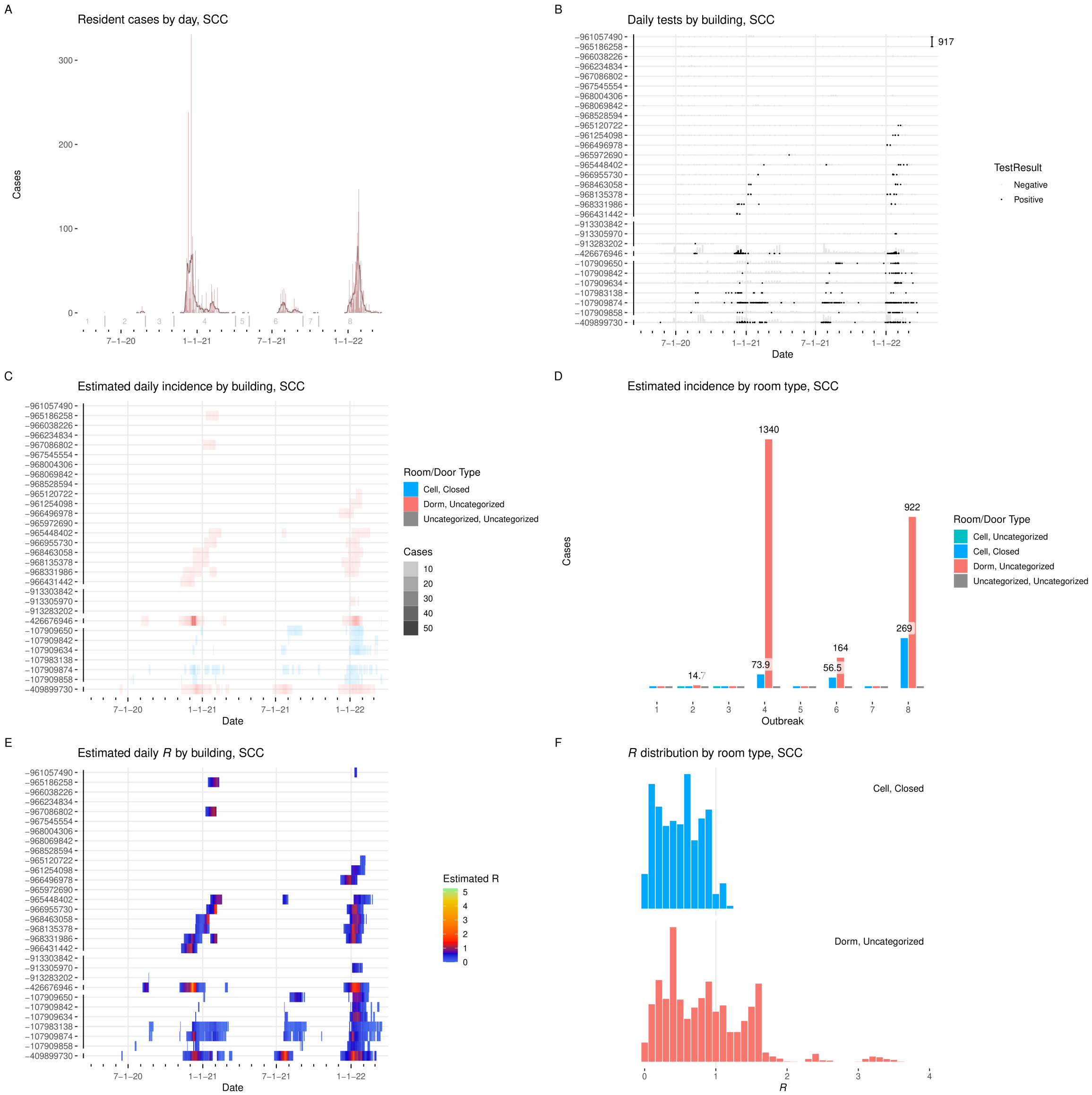


Figure I.30: **Results from SCC. A:** Cases detected per day, with seven-day average. Starting date of each outbreak is marked by a vertical gray line. **B:** Positive and negative tests per day by building. $Y$-axis labels are building ID numbers. Buildings are grouped by facility. Scale bar at upper right gives number of tests. **C:** Estimated incidence by day by building, colored by room type. **D:** Incidence by room type and outbreak. **E:** Estimated effective $R$ by day per building. **F:** Distribution of effective $R$ in each room type.

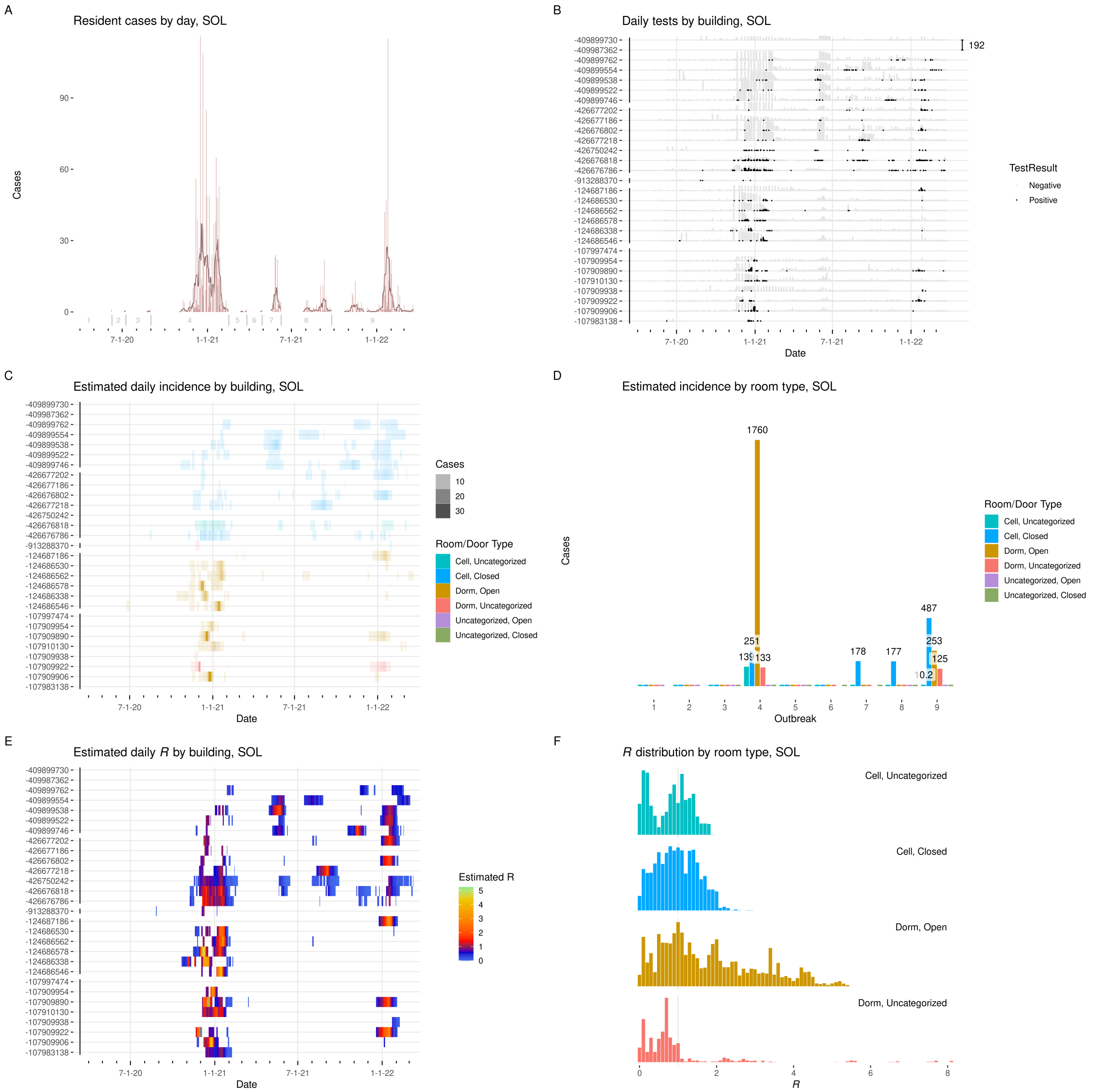


Figure I.31: **Results from SOL. A:** Cases detected per day, with seven-day average. Starting date of each outbreak is marked by a vertical gray line. **B:** Positive and negative tests per day by building. $Y$-axis labels are building ID numbers. Buildings are grouped by facility. Scale bar at upper right gives number of tests. **C:** Estimated incidence by day by building, colored by room type. **D:** Incidence by room type and outbreak. **E:** Estimated effective $R$ by day per building. **F:** Distribution of effective $R$ in each room type.

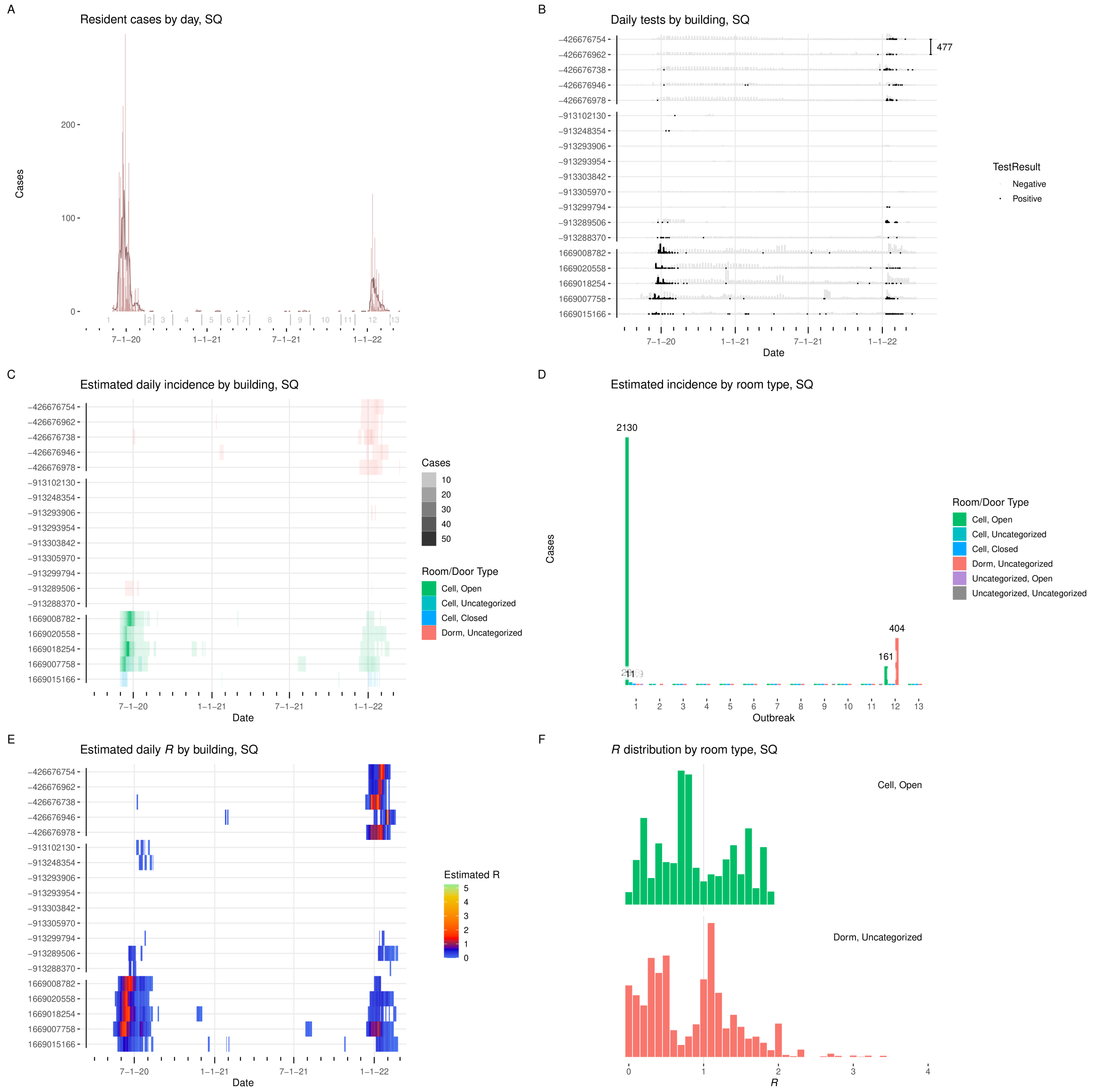


Figure I.32: **Results from SQ. A:** Cases detected per day, with seven-day average. Starting date of each outbreak is marked by a vertical gray line. **B:** Positive and negative tests per day by building. $Y$-axis labels are building ID numbers. Buildings are grouped by facility. Scale bar at upper right gives number of tests. **C:** Estimated incidence by day by building, colored by room type. **D:** Incidence by room type and outbreak. **E:** Estimated effective $R$ by day per building. **F:** Distribution of effective $R$ in each room type.

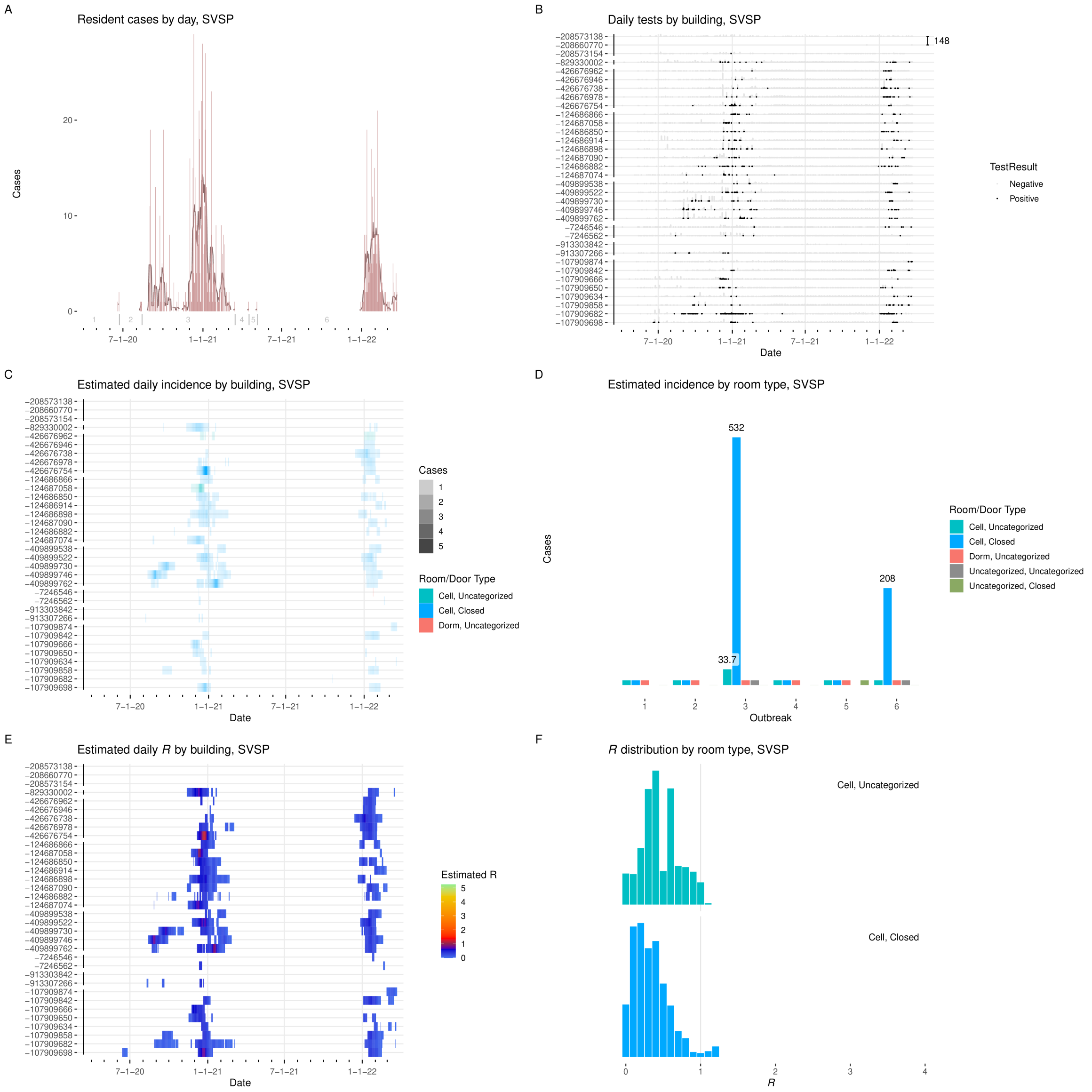


Figure I.33: **Results from SVSP. A:** Cases detected per day, with seven-day average. Starting date of each outbreak is marked by a vertical gray line. **B:** Positive and negative tests per day by building. $Y$-axis labels are building ID numbers. Buildings are grouped by facility. Scale bar at upper right gives number of tests. **C:** Estimated incidence by day by building, colored by room type. **D:** Incidence by room type and outbreak. **E:** Estimated effective $R$ by day per building. **F:** Distribution of effective $R$ in each room type.

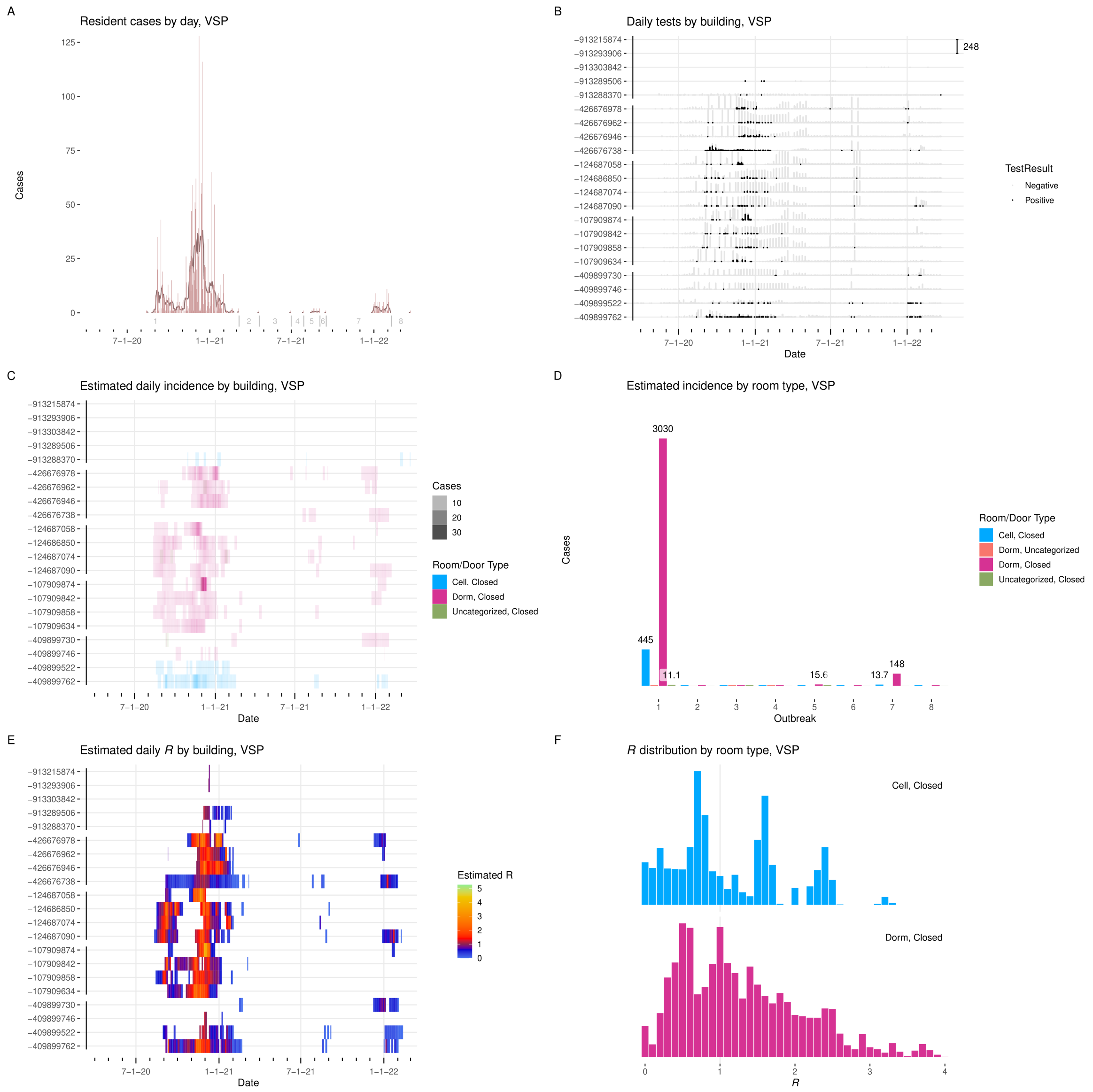


Figure I.34: **Results from VSP. A:** Cases detected per day, with seven-day average. Starting date of each outbreak is marked by a vertical gray line. **B:** Positive and negative tests per day by building. $Y$-axis labels are building ID numbers. Buildings are grouped by facility. Scale bar at upper right gives number of tests. **C:** Estimated incidence by day by building, colored by room type. **D:** Incidence by room type and outbreak. **E:** Estimated effective $R$ by day per building. **F:** Distribution of effective $R$ in each room type.

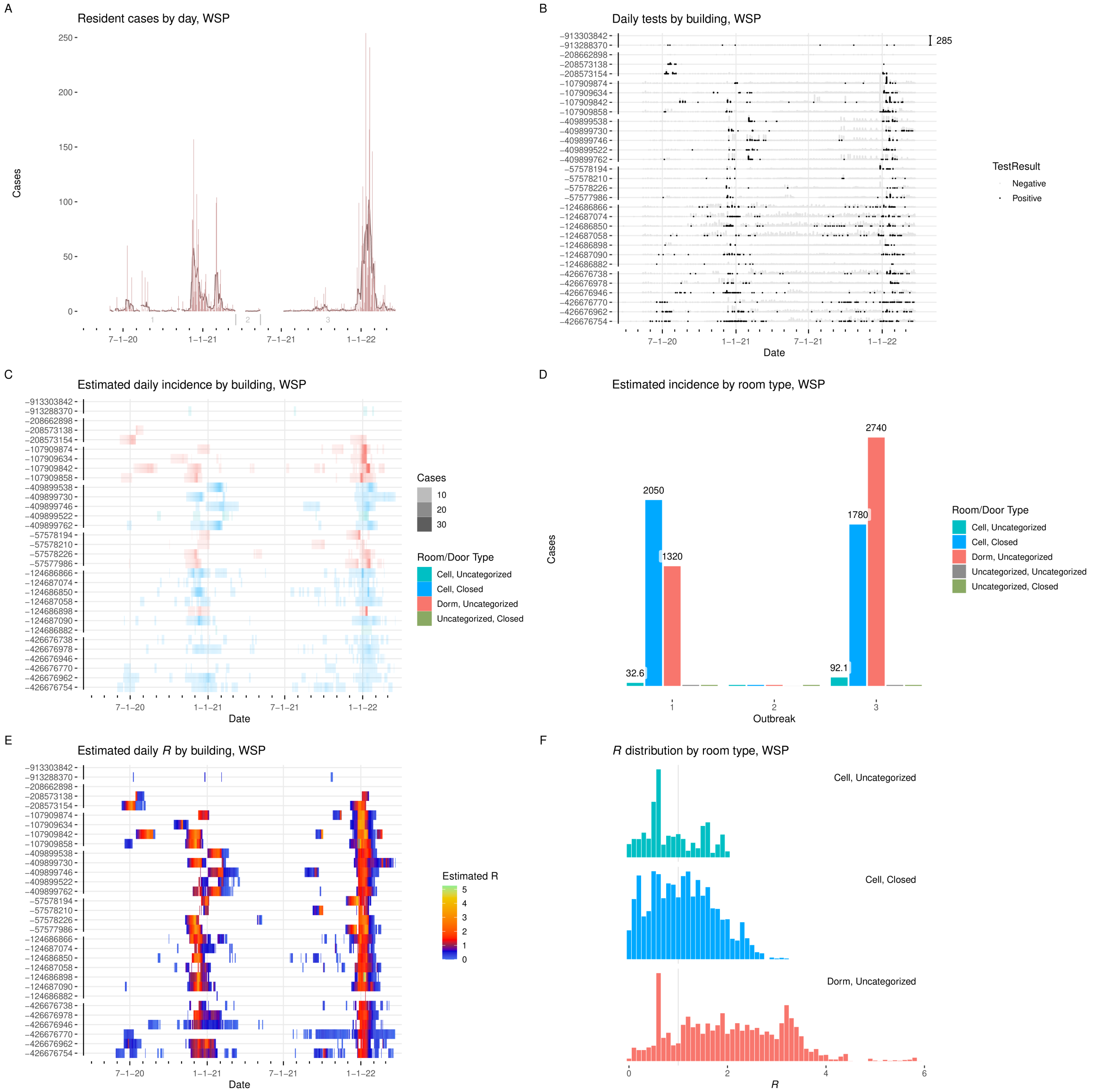


Figure I.35: **Results from WSP. A:** Cases detected per day, with seven-day average. Starting date of each outbreak is marked by a vertical gray line. **B:** Positive and negative tests per day by building. $Y$-axis labels are building ID numbers. Buildings are grouped by facility. Scale bar at upper right gives number of tests. **C:** Estimated incidence by day by building, colored by room type. **D:** Incidence by room type and outbreak. **E:** Estimated effective $R$ by day per building. **F:** Distribution of effective $R$ in each room type.

### Supplement J. Reproduction number and incidence estimates by room type

In Figures J.1–J.9, we have presented a panel of plots of the reconstructed incidence and reproductive numbers occurring in each room type separately.

Plot (A) of each panel showed reconstructed incidence by day at each institution, restricted to the rooms of the given type. Days on which that expected incidence was 1 or more cases at an institution were included.

Plot (B) of each panel showed the estimated reproduction number restricted to all cases housed in rooms of the given type at each institution each day. Days on which the expected total infectiousness profile (see Supplement A) over those cases was 1 or more at an institution were included.

In plot (C) the reconstructed incidence in the given room type was aggregated by season at each institution. Seasons in which the aggregated incidence was 1 or more cases were included, and values below 10.1 were summarized as <10.1.

### Supplement K. Supplemental information on limitations

*Supplement K.1. Smoothness*

The exact date on which specific residents became infected was not known, and could not be known in most cases. We constructed probabilistic estimates of these dates based on their history of test results and symptom reporting, reflecting what was known about their disease course, and equally reflecting the intrinsic uncertainty imposed by the limitations of available data. This uncertainty was reflected in our estimates, which took the form of a smooth curve of probability of, for instance, dates of infection of a case, rising from zero over several days to a peak at the most likely day, and then gradually declining again.

This uncertainty in dates of infection and infectiousness, appearing as smoothing of estimates over several days, was reflected by a corresponding smoothing of the estimated $R$ and aggregate incidence over time.

It may be that true reproduction numbers changed abruptly on specific dates in certain places, for example because of movement of many residents at once, or because of changes in HVAC system operation. While we would expect our estimates to reflect that a change in $R$ occurred, the intrinsic smoothing caused by granularity of the testing and symptom data was likely to cause the estimates to change gradually over several days from an earlier to a later value, even though the true change occurred all at once.

Similarly, if there were to be a sudden, brief moment of dangerous conditions, characterized by a quick spike in reproduction number, because of uncertainty in inferring timing from test data, the estimated $R$ would likely appear as a longer, smoother rise and fall, with the peak not as high as the true spike.

*Supplement K.2. Missingness of timing data at the beginning of outbreaks*

Several outbreaks in the CDCR system displayed a pattern in which the first, or nearly first, cases were detected by a large number of positive test results all on a single day. This pattern was seen, for example in the large Fall/Winter 2020–2021 outbreak at RJD (Figure K.1). A vertical bar marks the beginning of the outbreak (labeled outbreak 3). Nearly 200 cases were detected on a single day, after many days of no case detection, and then large numbers of cases were detected on subsequent days.

Our method of estimation of reproduction numbers required estimation of when transmission events occurred, in order to infer which individuals were source cases for others. A set of cases appearing all at once like this likely included multiple generations of transmission, but the data could provide very little indication how many generations from the data, since the timing of transmission was obscured by the lack of earlier test results. For this reason, estimates of reproduction numbers at the beginning of these outbreaks may have been less accurate than estimates for other time spans.

### Supplement L. Abbreviations for CDCR institutions

Table L.1 lists the names and abbreviations used for CDCR's institutions.

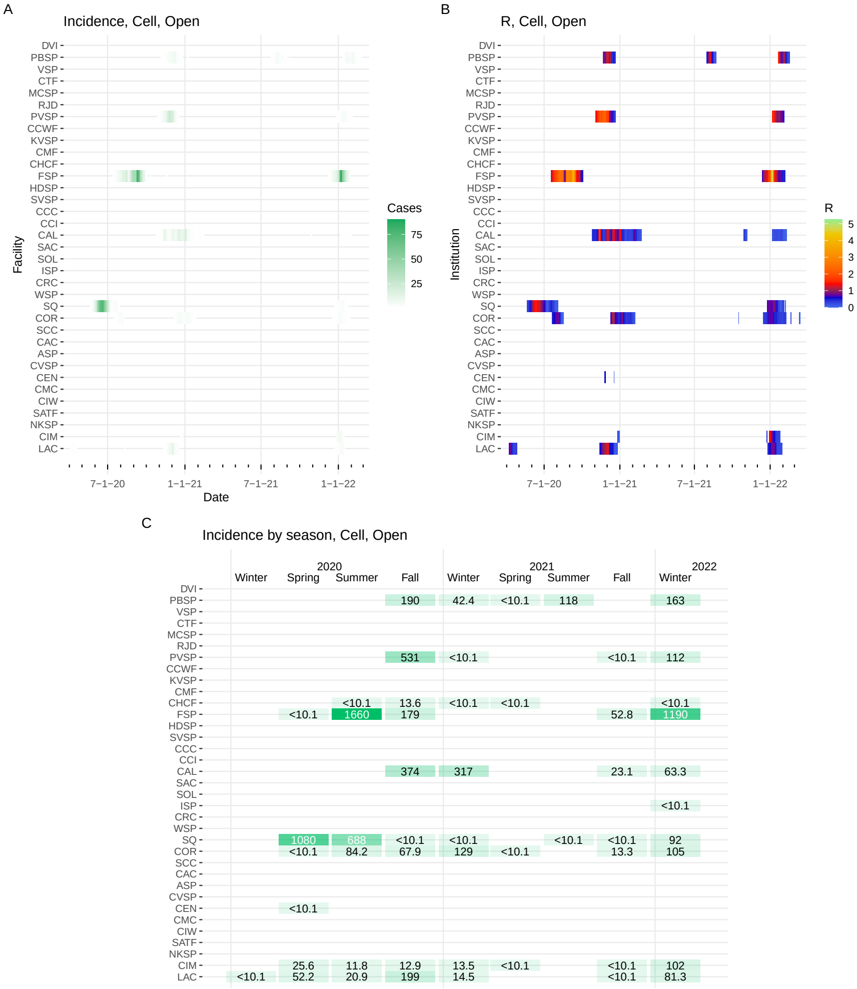


Figure J.1: **Statistics for room type Cell, Open. A:** Daily incidence at each institution, **B:** daily effective reproduction numbers, **C:** incidence by season.

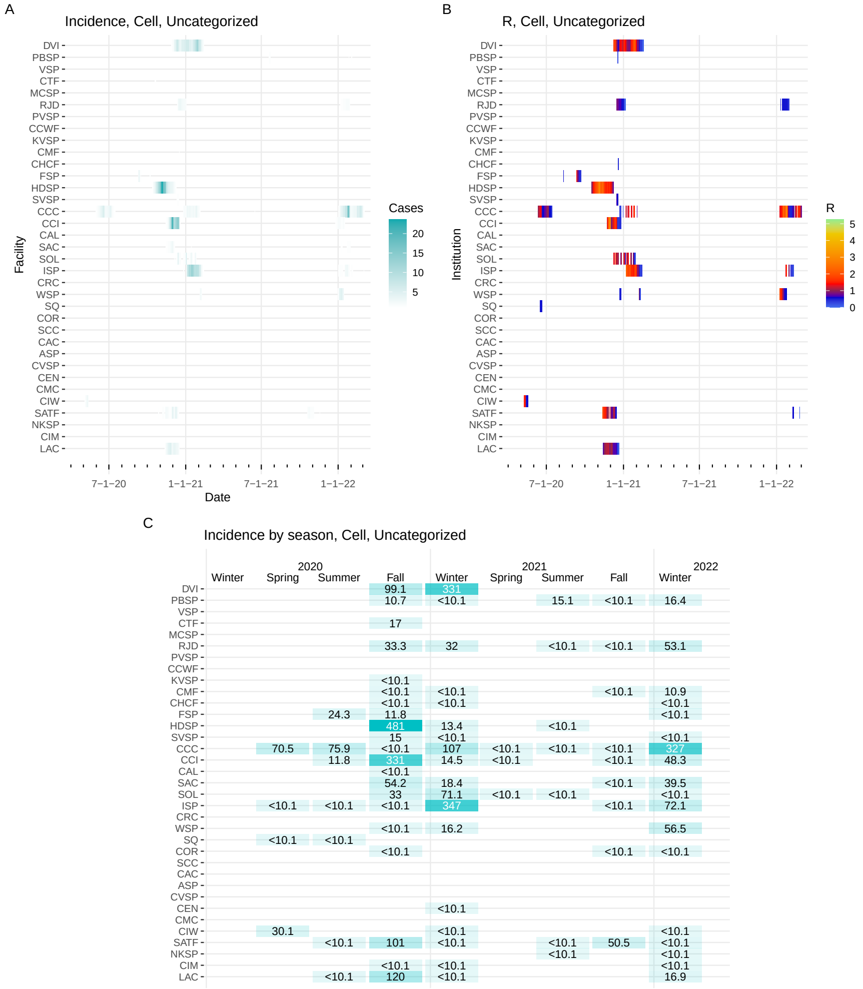


Figure J.2: **Statistics for room type Cell, Uncategorized.** **A:** Daily incidence at each institution, **B:** daily effective reproduction numbers, **C:** incidence by season.

A

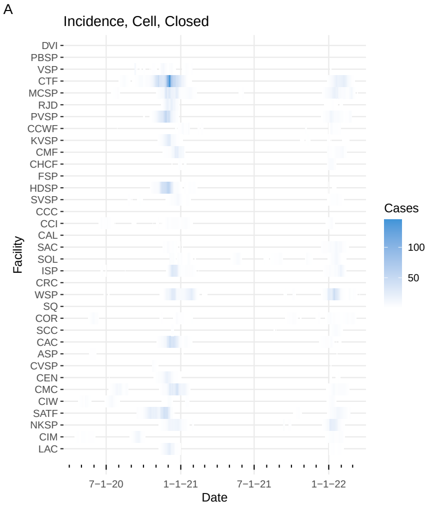


B

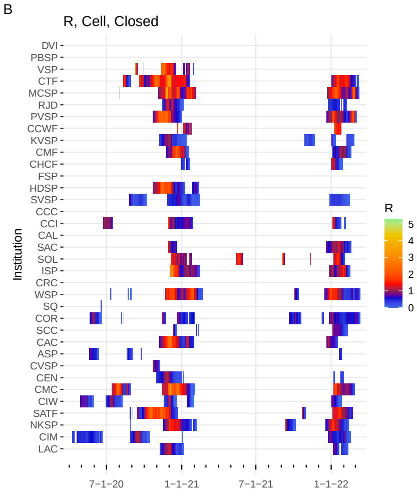


C

Incidence by season, Cell, Closed

| | 2020 | | | | 2021 | | | | 2022 |
|---|---|---|---|---|---|---|---|---|---|
| | Winter | Spring | Summer | Fall | Winter | Spring | Summer | Fall | Winter |
| DVI | | | | | | | | | |
| PBSP | | | | <10.1 | <10.1 | | <10.1 | | |
| VSP | | | 45.1 | 229 | 60.9 | | <10.1 | | 10.2 |
| CTF | | | 129 | 3240 | 595 | <10.1 | <10.1 | <10.1 | 800 |
| MCSP | | | 55.5 | 760 | 336 | 33.1 | 13.3 | 21.7 | 653 |
| RJD | | | | 446 | 41.9 | | | <10.1 | 63.4 |
| PVSP | | | <10.1 | 1380 | 38.2 | <10.1 | 16.3 | 18.5 | 367 |
| CCWF | | | <10.1 | 14.5 | 133 | | <10.1 | 17.3 | 104 |
| KVSP | | | <10.1 | 503 | 39.2 | <10.1 | | 52.9 | 70.3 |
| CMF | | | <10.1 | 225 | 286 | <10.1 | <10.1 | <10.1 | 191 |
| CHCF | | | <10.1 | 25.5 | 46.1 | <10.1 | <10.1 | 11 | 117 |
| FSP | | | | | | | | | |
| HDSP | | <10.1 | | 1490 | 125 | <10.1 | 15.1 | <10.1 | 39.3 |
| SVSP | | <10.1 | 40 | 110 | 141 | | | <10.1 | 111 |
| CCC | | | | | | | | | |
| CCI | | 23.4 | 117 | 126 | 137 | <10.1 | <10.1 | <10.1 | 72.6 |
| CAL | | | | | | | | | |
| SAC | | <10.1 | <10.1 | 86.1 | 30.2 | <10.1 | <10.1 | 33.7 | 304 |
| SOL | | | | 64.5 | 104 | 107 | 104 | 66.7 | 203 |
| ISP | | 13.4 | 41.1 | 474 | 271 | | | 13 | 325 |
| CRC | | | | | | | | | |
| WSP | | <10.1 | 30.8 | 531 | 662 | <10.1 | 31.3 | 70.5 | 1030 |
| SQ | | <10.1 | | | | | | | <10.1 |
| COR | | 103 | 16.3 | 48.1 | 87.2 | <10.1 | 15.1 | 120 | 204 |
| SCC | | | | 22.3 | 10.2 | | 29.1 | <10.1 | 132 |
| CAC | | | <10.1 | 934 | 243 | <10.1 | <10.1 | 30.6 | 94.4 |
| ASP | | 45.3 | 23.1 | <10.1 | <10.1 | | | | <10.1 |
| CVSP | | | | 38.7 | | | | | 10.8 |
| CEN | | 11.6 | 12.9 | 414 | 42.1 | | <10.1 | 25.8 | 57 |
| CMC | | 12.8 | 261 | 559 | 523 | | <10.1 | <10.1 | 233 |
| CIW | | 91.1 | 137 | 75.5 | 33.5 | | <10.1 | <10.1 | 83.7 |
| SATF | | | 25.9 | 1490 | <10.1 | | | 55.5 | 360 |
| NKSP | | <10.1 | 13.4 | 355 | 256 | <10.1 | 38.3 | 40.3 | 573 |
| CIM | | 112 | 146 | 87.4 | <10.1 | <10.1 | | <10.1 | 166 |
| LAC | | <10.1 | <10.1 | 442 | 31.2 | | <10.1 | <10.1 | 76.4 |

Figure J.3: **Statistics for room type Cell, Closed. A:** Daily incidence at each institution, **B:** daily effective reproduction numbers, **C:** incidence by season.

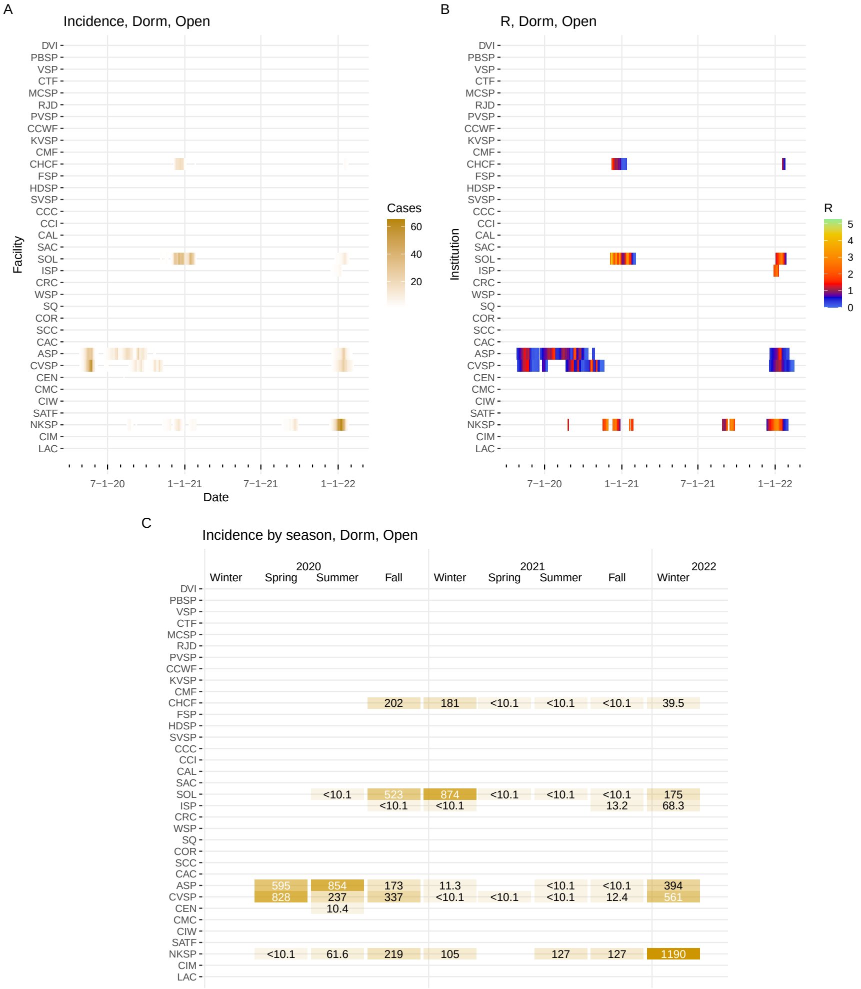


Figure J.4: **Statistics for room type Dorm, Open. A:** Daily incidence at each institution, **B:** daily effective reproduction numbers, **C:** incidence by season.

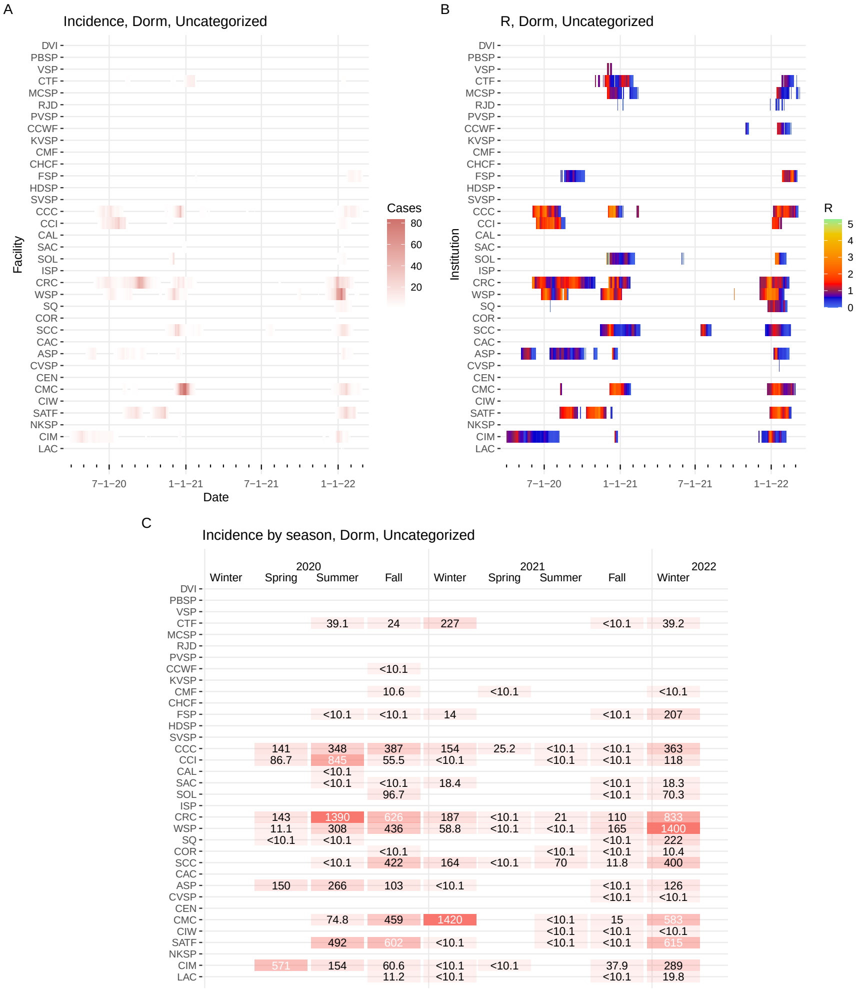


Figure J.5: **Statistics for room type Dorm, Uncategorized. A:** Daily incidence at each institution, **B:** daily effective reproduction numbers, **C:** incidence by season.

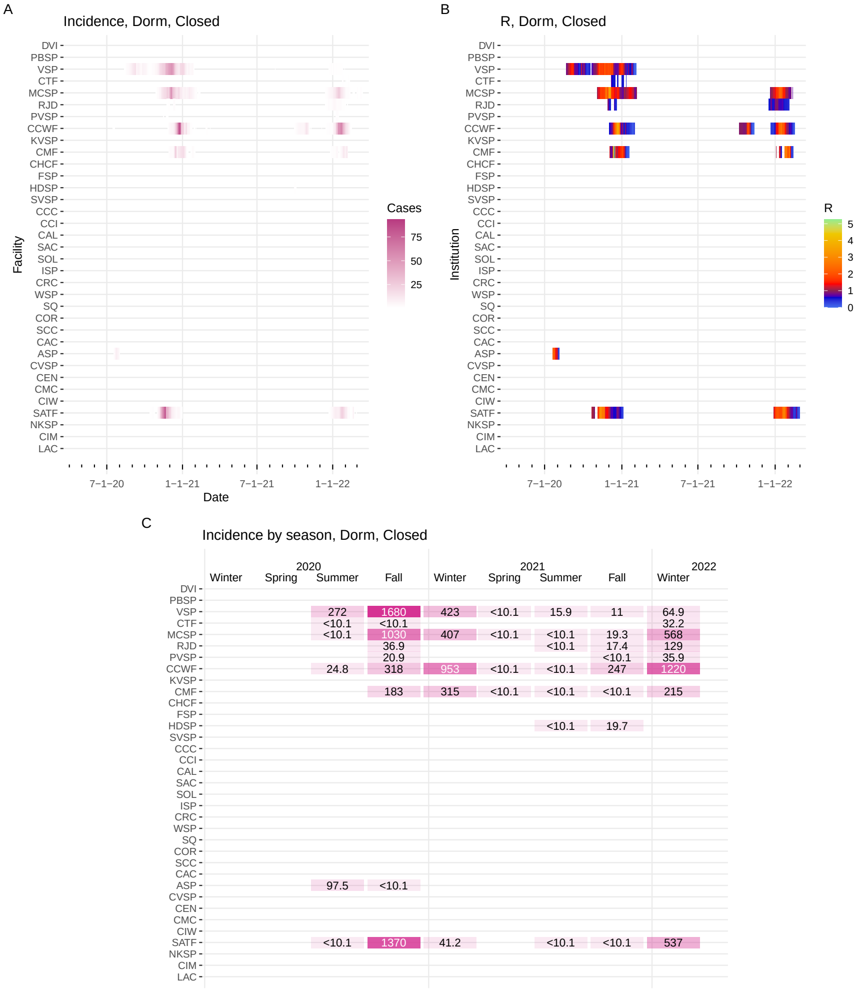


Figure J.6: **Statistics for room type Dorm, Closed. A:** Daily incidence at each institution, **B:** daily effective reproduction numbers, **C:** incidence by season.

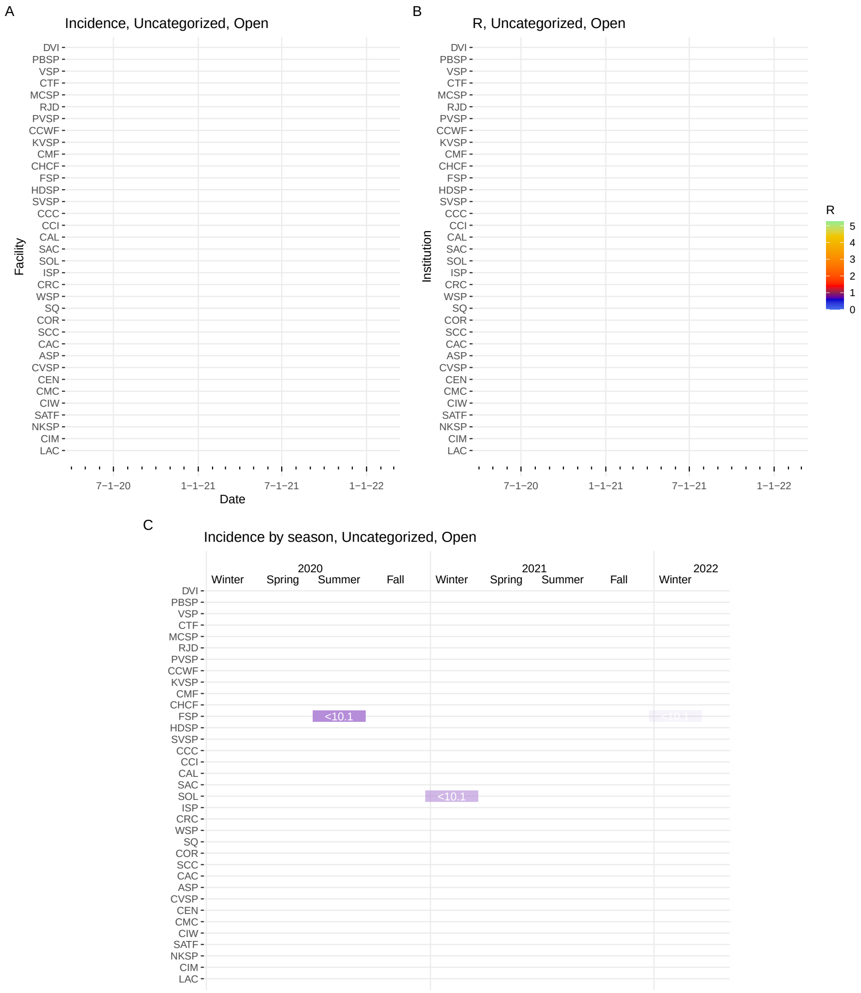


Figure J.7: **Statistics for room type Uncategorized, Open. A:** Daily incidence at each institution, **B:** daily effective reproduction numbers, **C:** incidence by season.

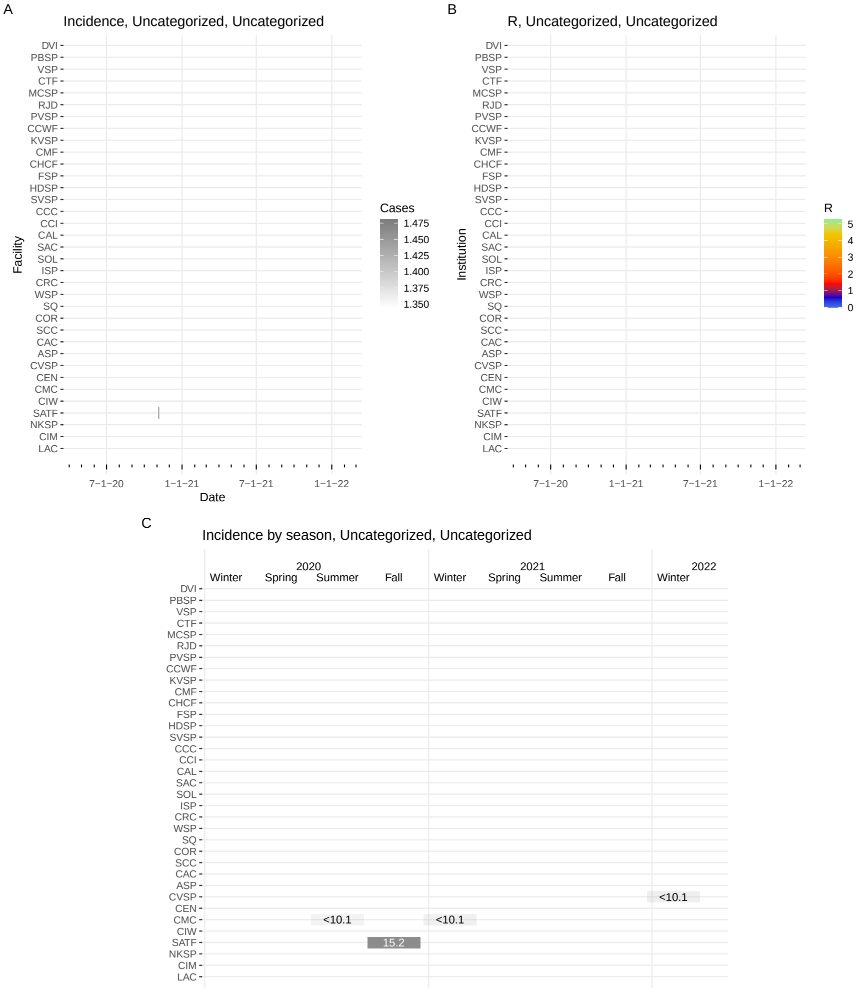


Figure J.8: **Statistics for room type Uncategorized, Uncategorized. A:** Daily incidence at each institution, **B:** daily effective reproduction numbers, **C:** incidence by season.

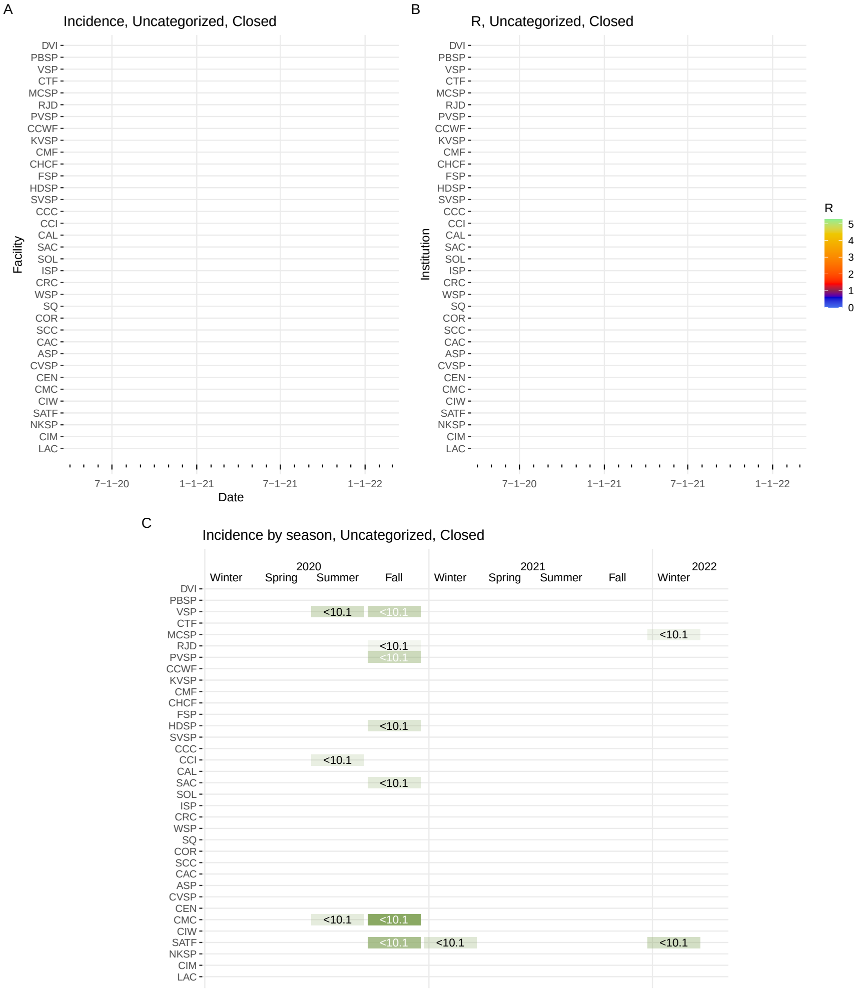


Figure J.9: **Statistics for room type Uncategorized, Closed. A:** Daily incidence at each institution, **B:** daily effective reproduction numbers, **C:** incidence by season.

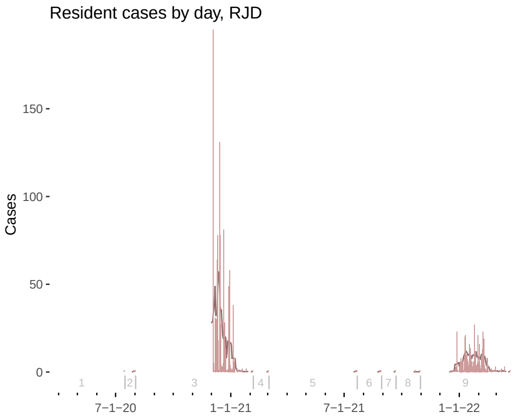


Figure K.1: Cases detected by day at RJD (light vertical bars), with seven-day average (dark curve). Vertical gray lines show the division of days into outbreaks.
